# Sample sizes to achieve multiple surveillance objectives in primary care sentinel systems monitoring respiratory pathogens: a simulation approach

**DOI:** 10.64898/2026.08.20.26360887

**Authors:** Anne M. Presanis, Tommy Nyberg, Melissa A. Rolfes, Catherine Quinot, Rosalind Goudie, Heather Whitaker, William H. Elson, Rachel Byford, Tima Mikdashi, Jessica Y. Wong, Nick Andrews, Sofia S Villar, Benjamin J. Cowling, Andre Charlett, Gavin Dabrera, Richard Pebody, Jamie Lopez-Bernal, Simon de Lusignan, Daniela De Angelis

**Affiliations:** MRC Biostatistics Unit, University of Cambridge, Cambridge, United Kingdom; National Center for Immunizations and Respiratory Diseases, US Centers for Disease Control and Prevention, Atlanta, United States; Immunisation and Vaccine Preventable Diseases, UK Health Security Agency, London, United Kingdom; Nuffield Department of Primary Care Health Sciences, University of Oxford, Oxford, United Kingdom; School of Public Health, Yale University, New Haven, United States; School of Public Health, The University of Hong Kong, Hong Kong; Analytics and Intelligence Assessment, UK Health Security Agency, London, United Kingdom; Epidemic and Emerging Infections, UK Health Security Agency, London, United Kingdom; Research and Surveillance Centre, Royal College of General Practitioners, Oxford, United Kingdom

**Keywords:** sentinel surveillance, design, sample sizes, respiratory infections

## Abstract

Influenza surveillance has typically been carried out using influenza-like illness (ILI) rates and proportions of laboratory tests positive for influenza as metrics to monitor, with sample sizes for the number of tests to carry out based on the precision of the resulting estimate of proportions positive. The transition out of the Severe Acute Respiratory Syndrome Coronavirus 2 (SARS-CoV-2) pandemic period has encouraged the establishment of integrated surveillance of respiratory pathogens, in the context of multiple surveillance objectives, as set out by WHO in its revised integrated surveillance guidance and Mosaic Respiratory Surveillance Framework. These objectives include outbreak detection, situational awareness and intensity evaluation, among others. We illustrate how to design respiratory surveillance in primary care, by considering multiple surveillance objectives for different metrics of different types of respiratory pathogen circulation seasons in England, the USA and Hong Kong. We focus on a proxy of influenza activity as a metric to compare between these countries/regions. Taking advantage of England’s integrated sentinel primary care surveillance system, we propose further metrics to monitor: a proxy of respiratory activity, novelly defined as the product of an acute respiratory infection (ARI) consultation rate and the proportion of tests positive for *at least one pathogen*; pathogen-specific ARI-based activity proxies for more detailed monitoring of influenza and SARS-CoV-2; and integrated monitoring of proportions positive for all pathogens tested. We use a simulation approach to determine sample sizes by optimising either the probability of, or time to, detection of different events in monitored metrics, according to the different surveillance objectives. We find that sample sizes to maximise detection probabilities or minimise detection times vary by metric, objective, event and country/region. At a national level, the current sample sizes used are sufficient to detect most events in most weeks for both the USA and Hong Kong, but for England the numbers of swabs taken for ILI consultations may not be sufficient in all weeks, particularly at the start of the season when outbreak detection is important. However, broadening the criteria for swabbing to acute respiratory symptoms does allow for sufficient sample sizes.

## 1 Introduction

The transition over the last few years out of the Severe Acute Respiratory Syndrome Coronavirus 2 (SARS-CoV-2) pandemic period of 2020-22 has been an opportunity to integrate surveillance of multiple respiratory viruses in a cohesive system, rather than considering each virus (e.g. influenza, SARS-CoV-2, respiratory syncytial virus [RSV]) in isolation. Several examples of integration efforts include: the UK Health Security Agency (UKHSA) consolidating its integrated surveillance of respiratory viruses (UK Health Security Agency 2023a, 2024a,b); the Oxford-Royal College of General Practitioners Research and Surveillance Centre (RSC) and UKHSA reconsidering definitions and recording of a hierarchical acute respiratory infection (ARI) phenotype (Elson et al. 2024; Gu et al. 2024); the UK Joint Committee on Vaccination and Immunisation (JCVI) recommending enhancing surveillance to sufficiently power real-world evaluation of vaccine effectiveness (Joint Committee on Vaccination and Immunisation 2024); and the World Health Organization (WHO) publishing guidance on integrated surveillance within the Global Influenza Surveillance and Response System (GISRS, World Health Organization 2022, 2024a), launching the Mosaic Respiratory Surveillance Framework (Mott et al. 2023; World Health Organization 2023) and revising its guidelines for Pandemic Influenza Severity Assessment (PISA, Bracher and Littek 2024; Eales et al. 2024; World Health Organization 2024b. The revised GISRS guidance (World Health Organization 2024a) sets out the objectives of integrated surveillance, via sentinel systems, in different broad groups: monitoring clinical, epidemiological and virological characteristics of circulating ARIs; providing evidence to guide interventions; and supporting early warning and event-based surveillance. The Mosaic framework is described as “resilient surveillance for respiratory viruses of epidemic and pandemic potential” (World Health Organization 2023), and similarly groups the objectives of respiratory pathogen surveillance into three domains: detection and assessment; monitoring epidemiological characteristics; and informing use of interventions. Fearon et al. 2026 set out the need for testing designed to achieve different objectives in these various groups and domains. Sentinel surveillance of respiratory infections in primary care is a key component contributing to all these objectives (Gu et al. 2024; World Health Organization 2023, 2024a).

Despite these initiatives, challenges remain in improving the design of sentinel surveillance systems for monitoring respiratory pathogens. Two of these challenges, in the context of primary care sentinel surveillance, include: monitoring suitable measures of disease circulation (“metrics”) to achieve multiple surveillance objectives; and ensuring sufficient sample sizes from such sentinel systems (“rightsizing”) to achieve them.

Primary care sentinel systems typically: record the number of individuals consulting a general practitioner (GP) or outpatient clinic with symptoms consistent with the Influenza-Like-Illness (ILI) case definition (Gu et al. 2024; World Health Organization 2024a); select a sample of these individuals from whom to take nasopharyngeal swabs to send for laboratory virological testing; and record the results of such testing. Two metrics are often used widely for situational awareness of influenza epidemics and seasons: the ILI consultation rate, where the denominator in most countries is the total number of outpatient or GP consultations, but in England is the population covered by the GP sentinel system; and the proportion of patients with ILI whose swab samples test positive for influenza, among those tested. However, the magnitude of these metrics depends on healthcare-seeking behaviour, resulting testing patterns, test sensitivity, and cocirculation of non-influenza respiratory pathogens, all of which may vary over time (Eales et al. 2023, 2024). The interpretation of the ILI rate and proportion positive metrics as measures of influenza circulation is therefore problematic, motivating the use, instead, of a metric (sometimes denoted ILI+) defined as the product of the two, as a proxy for influenza activity, i.e. a measure of current circulation (Goldstein et al. 2012). During periods with co-circulating respiratory viruses, Eales et al. 2024 have shown mathematically that ILI+ is a more appropriate indicator of pathogen-specific incidence than the ILI rate or proportion positive. ILI+ has been used in various studies to estimate influenza-attributable mortality (Goldstein et al. 2012; Wong et al. 2013; Wu et al. 2017) and influenza incidence (Birrell et al. 2011); and has been assessed as a metric to monitor by various countries (e.g. AbdElGawad et al. 2020; Tay et al. 2013), particularly following its recommendation by WHO (World Health Organization 2017). However, it has not yet been widely adopted as a metric to monitor in real time. Furthermore, although ILI symptoms are appropriate for ensuring the ILI+ metric is a good proxy for influenza incidence (Eales et al. 2024; Wong et al. 2013), they may not be appropriate for other pathogens. For example, when the omicron variant of SARS-CoV-2 emerged, it was noted that its symptom profile changed, affecting the lower respiratory tract less than previous variants (Vihta et al. 2023). The broad range of symptoms associated with SARS-CoV-2 meant that a wider set of criteria for swabbing was in use in England during the pandemic, and this was formalised post-pandemic with the RSC’s ARI phenotyping algorithm. This algorithm is used to identify cases of ARI from a patient’s electronic health record, covering the range of clinical syndromes that fall under the umbrella of ARIs and identifying patients for whom a swab is indicated. These clinical syndromes include ILI, exacerbations of chronic lung disease (ECLDs), lower respiratory tract infections (LRTIs), upper respiratory tract infections (URTIs), and other ARIs. Cases of these clinical syndromes are identified from a patient’s record using clinical code lists that include diagnostic and symptom codes associated with each syndrome (Elson et al. 2024; Gu et al. 2024). GPs are advised to code ARI episodes in the following order of priority: ILI, ECLD, LRTI, URTI, or other not-specified ARI. This increased richness of coding gives opportunities to investigate the use of ARI and/or its components in defining activity proxies to monitor that may be more suited to integrated surveillance than ILI alone or ILI-based proxies.

In terms of rightsizing for virological and serological testing, historically, a single surveillance objective at a time has been pursued, for example to ensure sufficient precision of estimates: of the proportion testing positive for influenza to detect when it crosses a threshold (epidemic detection, including detecting new strains or variants, Association of Public Health Laboratories 2022; World Health Organization 2017); or of vaccine effectiveness to determine vaccine coverage needed (Chung et al. 2021; Whitaker et al. 2024). In these cases, precision of a specific estimate is a “target” quantity to optimise for a single surveillance objective. However, surveillance systems often aim to meet multiple objectives for multiple respiratory viruses, such as detecting epidemics, providing situational awareness, monitoring transmission and severity of activity, and intervention evaluation, among others (World Health Organization 2023, 2024a). Precision is not the only target quantity that should be optimised to optimally design surveillance to achieve these objectives. Sample sizes can be determined also to maximise our *power to detect* an event; and to minimise the *time to detection* of an event.

To address these challenges in the context of these multiple objectives and target quantities, we propose a set of metrics to monitor, along with a set of events these metrics experience that should be detected, and an approach to determine sample sizes for event detection using power to detect and time to detection as target quantities to optimise.

### Metrics

The metrics we consider are all in the form of proportions or rates. We assume that any proportions monitored are *estimated*, i.e. point estimates are given by the maximum likelihood estimate which is the observed numerator divided by the observed denominator; and binomial confidence intervals are obtained to monitor the uncertainty in the metrics. Throughout, we consider the ILI or ARI rates to be known quantities, fixed to their observed values, since the numbers of ILI or ARI consultations are typically large relative to the number of swabs tested. Therefore we assume any uncertainty in the ILI+ activity proxy and other activity proxies defined below is driven by the uncertainty in the proportions positive for specific viruses.

We first consider the ILI+ metric for single-pathogen (influenza) objectives, using surveillance data from the 2022/23 season or 2023 year for three countries/regions with different types of influenza circulation: the temperate winter season in England in 2022/23 when all three of influenza, SARS-CoV-2 and RSV were circulating; the temperate winter influenza season in the USA in 2022/23; and the non-temperate influenza year in Hong Kong in 2023. For data from England, we additionally consider other metrics in the interest of integrated surveillance. First, we propose a new proxy of any respiratory infection activity, which we denote ARI+, defined as the product of the ARI GP consultation rate and the proportion of swabs that test positive for *at least one* monitored pathogen. Second, we contrast this broad indication of activity with two ARI-based pathogen-specific proxies: ARI-flu defined as the product of the ARI GP consultation rate and the proportion of swabs that test positive for influenza, originally recommended by WHO (World Health Organization 2017, 2024b); and ARI-SC2 defined as the product of the ARI GP consultation rate and the proportion of swabs that test positive for SARS-CoV-2. We compare the ARI-flu proxy with the ILI+ proxy. Finally, we consider the proportion positive metric for achieving multi-pathogen objectives, where we consider jointly all respiratory viruses tested in England.

### Events

We demonstrate sample size determination through defining a set of events to detect that address different aspects of surveillance objectives such as those in the GISRS integrated surveillance guidelines (World Health Organization 2024a) and the Mosaic Domains (World Health Organization 2023) concerning detection, assessment, monitoring of epidemiologic characteristics and situational awareness. These events include: threshold-based events, where estimated values of either a metric or its growth rate cross a threshold, either in absolute terms or with statistical confidence if their confidence intervals also cross the threshold; and consecutive weekly increases or decreases in the estimated values of a metric.

### Sample size determination

Optimising the proposed targets analytically is challenging, as the power and time to detect events of interest are difficult to express analytically as a function of sample size. Therefore, we use a simulation approach, based on seasonal respiratory virus surveillance data from 2022/23, to illustrate choosing sample sizes to optimise detection of different types of event for one particular season/year.

We set out our simulation approach in Section 2, illustrate which samples are required to reach target power and time to detect events in Section 3, and end with a discussion in Section 4.

## 2 Methods

Our simulation approach consisted of defining, for each country/region, a “ground truth” 2022/23 season dataset based on smoothing the observed proportions positive for influenza and ILI+ rates; and for England, additionally, ground truth season datasets based on the smoothed proportions positive for each tested pathogen and the ARI-based activity proxies. These ground truth metrics were taken to be the expected values from which we simulate multiple surveillance datasets, assuming different weekly sample sizes. We assessed the sample sizes and power to detect an event in terms of the probability of detection and the time to detection. The probabilities of detection were obtained as the proportion of the simulated datasets where the event of interest occurs within the period of time defined by each season considered. The times to detection were defined conditional on detection occurring within the season, i.e. based on the simulated datasets where the event of interest occurs, and were defined as the time between the true event occurring in the ground truth dataset from which the simulated datasets are drawn and the detected event in each simulation. Note that if the event did not occur in the ground truth dataset within the season or if it did not occur in *any* of the simulated datasets, then the detection time is undefined.

We describe the data sources considered in Section 2.1 and current surveillance systems and the metrics they record in Section 2.2. We define the events we aim to detect to achieve different surveillance objectives in Section 2.3 and give details of the simulations in Section 2.4.

### 2.1 Data sources

For England, individually-linked, pseudo-anonymised data on consultations for ILI and ARI and swab test results for the 2022/23 season were available through the RSC (Gu et al. 2024).

Corresponding aggregate summary data are published regularly (e.g. https://www.gov.uk/government/statistics/national-flu-and-covid-19-surveillance-reports-2022-to-2023-season; https://www.rcgp.org.uk/representing-you/research-at-rcgp/research-surveillance-centre/public-health-data). The RSC pseudo-anonymised individual-level surveillance data were collected by NHS England and the UK Health Security Agency with permissions granted under Regulation 3 of The Health Service (Control of Patient Information) Regulations 2002, and without explicit patient permission under Section 2.1 of the NHS Act 2006.

Aggregate surveillance data for the USA were publicly available through Centers for Disease Control and Prevention Fluview (https://gis.cdc.gov/grasp/fluview/fluportaldashboard.html), with weekly outpatient visits (with ILI and total) sourced from ILINet and virological positivity data reported to CDC from U.S. Influenza/NREVSS collaborating clinical laboratories. Aggregate data from Hong Kong were publicly available from Hong Kong Centre for Health Protection Flu Express (https://www.chp.gov.hk/en/resources/29/100148.html).

### 2.2 Primary care sentinel surveillance: metrics recorded

In England, sentinel surveillance for respiratory pathogens in primary care is coordinated by the RSC, with around 2, 000 practices taking part now (Gu et al. 2024), of whom around 500 practices contributed to the 2022/23 season collection. The currently involved practices cover 31.9% of the population of England and Wales and are considered geographically and demographically representative (Leston et al. 2022). Denote by *N_P,t_*the total population size of England in week *t* and by *g_P,t,s_* the number of consultations at GPs in the total population for symptoms consistent with either influenza-like illness (*s* = ILI) or acute respiratory infection (*s* = ARI) in week *t*. Note that these consultations may be defined retrospectively based on events following the consultation, such as test results or secondary care, but are counted based on the consultation date, where symptoms have been recorded within 10 days of the consultation. Then the weekly consultation rate per 100, 000 population is 100, 000×*g_P,t,s_/N_P,t_*. This quantity is not observed, since the sentinel surveillance only covers a portion of the total population. Let *N_S,t_* be the size of the sentinel population covered by the participating practices in the winter season of 2022/23 and *g_t,s_* be the number of consultations in week *t* for symptoms consistent with *s* = ILI or *s* = ARI, where both these quantities are observed. Then the weekly consultation rate per 100, 000 population 100, 000 × *g_t,s_/N_S,t_* can be considered an estimator for the unobserved ILI or ARI consultation rate in the general population, 100, 000 × *g_P,t,s_/N_P,t_*, provided the sentinel population is representative of the general population. Figure A.1 in Appendix A shows the large difference in magnitude between the ILI and ARI consultation rates in England during the 2022/23 season.

The new ARI phenotype (Elson et al. 2024; Gu et al. 2024) was being introduced during the 2022/23 season, such that in this analysis, we consider three subsets of swabs taken for testing, either from patients with: (a) ILI symptoms only; (b) ARI symptoms (consistent with the new phenotype), including ILI, URTI, LRTI, suspected COVID-19 and bronchiolitis; or (c) a broader ARI definition, including also fever without other ILI symptoms and loss of sense of smell or taste. We denote symptom profile (c) by *s* = ALL, with the corresponding number of consultations denoted *g_t,ALL_*. We consider the number of swabs taken from those with ILI symptoms *n_t,ILI_* as a denominator for the number of ILI cases testing positive for influenza *y_t,Flu,ILI_* for the single pathogen analyses using the ILI+ metric, i.e. the rate

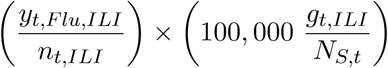

(Figure 1, third row). For analyses using ARI-based activity proxies, we used the total number of swabs, *n_t,ALL_*, as the denominator for influenza and SARS-CoV-2 proportions positive and for the proportion of swabs that test positive for at least one pathogen. The activity proxies are therefore defined as

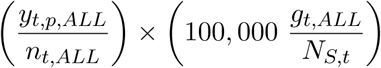

for *p* indexing the pathogen (influenza [*F lu*], SARS-CoV-2 [*SC*2] or at least one pathogen among all tested, Figure A.2 in Appendix A). For integrated analyses of positivity for multiple pathogens, we again used the total number of swabs *n_t,ALL_*as a denominator for the test-positive cases of each pathogen: (*y_t,Flu,ALL_*; *y_t,SC_*_2_*_,ALL_*; *y_t,RSV,ALL_*; *y_t,Pos,ALL_*) (Figure A.3 in Appendix A), where *Pos* denotes positive for any other respiratory virus tested (other coronaviruses, human metapneumovirus, adenovirus, enterovirus, human rhinovirus) and *Neg* denotes negative for all the pathogens tested. Figure A.4 in Appendix A compares the influenza activity proxies using either ILI or ALL consultation rates as the pool from which swabs are collected.

**Figure 1:**
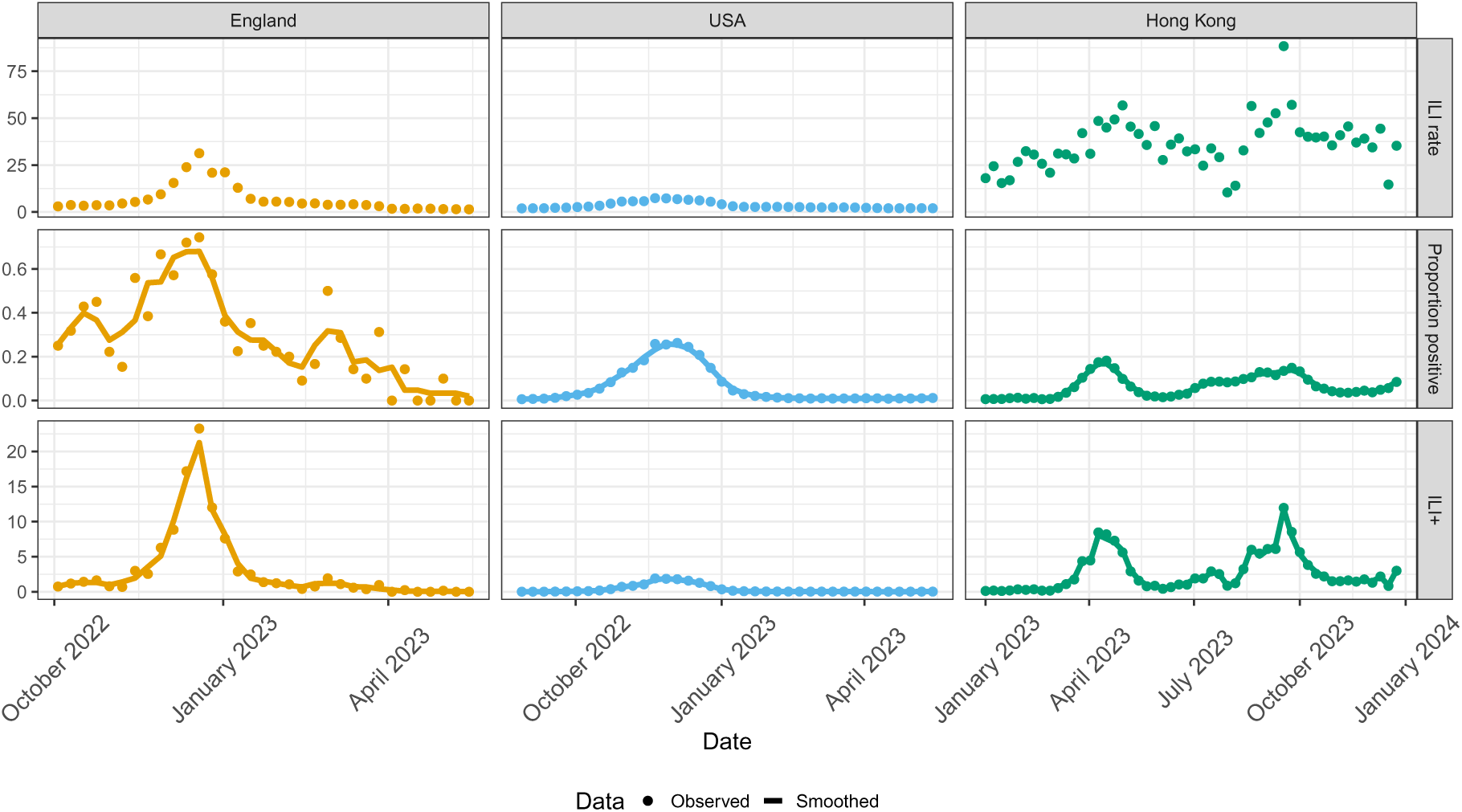
Observed metrics from 2022/23 by country/region (columns: England, USA, Hong Kong). Top row: ILI consultation rates (per 100,000 population in England; per 100 outpatient consultations in the USA; per 1,000 outpatient consultations in Hong Kong. Middle row: proportion of swabs testing positive for influenza, with smoothed version using 3-week moving averages). Bottom row: influenza activity proxy ILI+, with smoothed version using 3-week moving averages.

For the USA and Hong Kong, the sentinel systems included in the analysis cover outpatient clinics (rather than GPs as in England) and the population covered by an outpatient clinic is not available, or even well-defined. These sentinel systems therefore record the proportion *of all outpatient consultations* that are due to ILI symptoms, expressed as a rate per 100 or 1,000 outpatient consultations per week respectively. For Hong Kong and USA, denote by *g_t,ILI_* the number of ILI outpatient consultations in week *t*; and by *N_S,t_* the total number of outpatient consultations, so that the observed proportion consulting for ILI symptoms is *g_t,ILI_/N_S,t_*.

Figure 1 shows the observed ILI rates or proportions, influenza positivity and influenza activity proxy in the 2022/23 season for England and the USA, and in the whole 2023 year for Hong Kong. Note the differing scales across countries/regions for each metric, reflecting the different definitions of the ILI rate or proportion.

### 2.3 Events to detect: definitions

For each surveillance objective, we defined a range of events to detect that were elicited from discussions with surveillance experts in collaborating public health agencies and a technical working group on rightsizing for WHO’s revised GISRS integrated surveillance guidance (World Health Organization 2024a). Table 1 shows these events for any monitored metric, organised by the objectives that detection of the events achieves.

**Table 1:** Selection of events to detect for different surveillance objectives.

| Objective type | Specific aim/question | Event number | Event to detect |
| --- | --- | --- | --- |
| Detection & Assessment | Epidemic detection, intensity evaluation | 1 | metric ≥ a specified threshold |
| | | 2 | metric ≥ a specified threshold for $\geq 2$ consecutive weeks |
| | Confidence in epidemic detection, intensity evaluation | 3 | lower bound of 95% CI of metric ≥ a specified threshold |
| | | 4 | lower bound of 95% CI of metric ≥ a specified threshold for $\geq 2$ consecutive weeks |
| | Epidemic detection, peak assessment | 5 | $\geq 3$ consecutive weekly increases in a metric |
| | | 6 | $\geq 3$ consecutive weekly decreases in a metric |
| | Confidence in epidemic detection, peak assessment | 7 | lower bound of 95% CI for growth rate in a metric ≥ a specified threshold, based on data since the start of the season |
| | | 8 | upper bound of 95% CI for decline rate in a metric ≥ a specified threshold, based on data since the start of the season |
| Monitoring & Situational Awareness | Is the current trend increasing? | 9 | growth rate in a metric ≥ a specified threshold, data over the previous 4 weeks |
| | Confidence in a current increase | 10 | lower bound of 95% CI for growth rate in a metric ≥ a specified threshold, data over the previous 4 weeks |
| | Is the current trend decreasing? | 11 | decline rate in a metric ≤ a specified threshold, data over the previous 4 weeks |
| | Confidence in a current decrease | 12 | upper bound of 95% CI for decline rate in a metric ≤ a specified threshold, data over the previous 4 weeks |

#### Epidemic detection

At the Detection phase of a new epidemic or new respiratory infection season, Events 1-5 and 7 reflect different aspects of epidemic detection. Events 1 and 2 concern the estimated value of a metric crossing a threshold, possibly for a sustained period of time. Events 3 and 4 correspond to Events 1 and 2, but concern confidence, in terms of statistical significance, in having detected an epidemic, since they consist of the lower bound of a 95% confidence interval (CI) being greater than a threshold, possibly for a sustained period of time. Event 5 is defined as a sustained increase (at least 3 consecutive weekly increases) in the estimated value of a metric, based on the cumulative data over the course of a season. Detecting Event 5 is an alternative approach to being certain about an epidemic taking off. Detection of Event 7 frames confidence in epidemic detection in terms of comparing the lower bound of a 95% CI for the growth rate of a metric crossing a threshold. The growth rate is estimated by fitting a log-linear regression of the metric on calendar week, based on all data since the start of the season/observation period. In particular, for either the proportion positive or the activity proxy metric, a Poisson regression with log link is fitted to the number of positive tests, with an offset defined to be the appropriate denominator for each metric.

#### Intensity evaluation

In the context of seasonal epidemics, Events 1-4 can also be used in evaluating the “intensity” of the season, through comparison to a threshold based on historical data (Bracher and Littek 2024; World Health Organization 2024b). Various public health agencies define “intensity” in terms of peak levels observed in either ILI or ARI consultation rates or activity proxy metrics, where historical data on the peak levels of these metrics are used to define different levels of activity, using either the Moving Epidemic Method (MEM, defining levels of intensity, e.g. low, medium, high, based on quantiles of a normal distribution fitted to the peak values of previous seasons on an appropriate scale (Vega et al. 2013)) or some variation on it (Bracher and Littek 2024), standard deviation-based thresholds (Sinnathamby et al. 2024; World Health Organization 2024b), or expert knowledge.

#### Peak assessment

Detecting Events 6 and 8 are two alternatives for monitoring the peak of an epidemic. Event 6 is the analogue of Event 5, but defined in terms of sustained weekly decreases in the estimated value of a metric. Event 8 is the analogue of Event 7, concerning confidence, in terms of statistical significance, in the decline rate of a metric crossing a specified threshold.

#### Monitoring & situational awareness

Detecting Events 9-12 allows current situational awareness, by determining, based on the most recent four weeks of data, whether the current trend in the estimated values of a metric is increasing or decreasing in each week (9 and 11), with confidence (10 and 12). The same log-linear regression of a metric on calendar week is used to estimate growth/decline rates as for Events 7-8, with the only difference being the amount of data used to estimate them, four weeks rather than all data from the start of the season.

#### Integrated surveillance

For the integrated surveillance analyses, Events 5 and 6 are re-defined to being ≥ 3 consecutive weeks of increase/decrease in *at least one of the four groups of monitored pathogens*, influenza, SARS-CoV-2, RSV and any other respiratory pathogen tested. Similarly, Events 7-12 become growth/decline rates greater/smaller than a specified threshold in at least one of the monitored pathogens. Given potential correlation between circulating pathogens (Eales et al. 2023, 2024), for estimating the growth/decline rates, we consider a multinomial log-linear regression, implemented as a series of conditional Poisson log-linear models.

#### Thresholds

For Events 1-4 which involve thresholds for the estimated values of a metric, we used two thresholds, one lower and one higher (Table B.1 in Appendix B), for each country/region and metric. Each threshold was set either based on historical baselines or in consultation with experts in the collaborating public health agencies and the above-mentioned WHO technical working group (denoted “expert knowledge” hereafter).

For ILI rates in England, we used thresholds defined as baseline and low based on applying MEM to 10 years historical data prior to the 2022/23 season, excluding the 2020-2021 acute SARS-CoV-2 pandemic era when ILI rates were disrupted, as reported by UKHSA (UK Health Security Agency 2022, 2023b). For the proportion positive for influenza out of ILI cases tested in England, we chose two thresholds based on expert knowledge of previous seasons. Combining these thresholds resulted in four thresholds to use for the ILI+ metric in England (Table B.1). For ARI consultation rates, since the 2022/23 season was the first to systematically code for ARI, we could not use historical data to inform thresholds. We therefore chose two thresholds each for the consultation rates and proportions positive, based on expert knowledge of the GP surveillance system.

For the USA ILI consultation rate per 100 patient visits, we chose the national baseline for the 2022/23 season reported by CDC based on an average of non-influenza weeks in two previous seasons plus two standard deviations (US Centers for Disease Control 2022, 2023). For the proportion positive for influenza out of those tested in the USA, we used 0.1 and 0.15 as a lower and upper threshold, resulting in two thresholds for the ILI+ metric.

For Hong Kong, we used a baseline threshold for the proportion positive for influenza based on an average of 5 years pre-COVID-19 pandemic historical data (2014-2019) for non-influenza weeks plus two standard deviations (Hong Kong Centre for Health Protection 2023). We additionally used a higher threshold of 0.15. We also set a single threshold for the ILI consultation rate per 1000 patient visits in Hong Kong to be 30, resulting in two thresholds for the ILI+ metric.

For Events 7-12, involving thresholds for estimated growth or decline rates in a metric, we used 1.0, 1.05 and 1.1 as thresholds for the growth rate and 1.0, 0.95 and 0.9 as thresholds for the decline rate in each metric in each country/region. These values represent any increase, at least a 5% increase and at least a 10% increase in the metric respectively for the growth rates; and any decrease, at least a 5% decrease and at least a 10% decrease in the metric respectively for the decline rates.

### 2.4 Simulation

For each country/region *c* (index dropped for brevity in what follows, see Table 2 for a glossary of notation), we simulated a hypothetical surveillance system. We simulated *I* = 2, 000 datasets for each country/region, metric and for each of a range of sample sizes. We then calculate the power and time to detect targets for each sample size by summarising detection across the *I* datasets.

To define ground truth metrics from which to simulate the datasets, we assume the ILI or ARI consultation rates/proportions are known and fixed to their observed values *α* × *g_t,s_/N_S,t_*where *α* is the country/region-specific scale for the consultation rates (per 100, 000; per 100; and per 1, 000 for England, USA and Hong Kong respectively). We focus, therefore, on choosing sample sizes only for the subset of individuals consulting for ILI or ARI who are selected for swabbing and virological testing, of size *n_t,s_, s* ∈ {*ILI, ALL*}.

We start with simulating the influenza proportion positive, obtaining the ILI+ metric as the product of the simulated proportion positive and the fixed ground truth ILI rate. Let *d* = 1*, . . . ,* 6 index the sample sizes (i.e. the number of patients tested for influenza each week) *n_ILI,d_* ∈ {25, 50, 100, 250, 500, 1000} considered, assuming these denominators for the proportion positive are constant over all weeks *t* = 1*, . . . , T* in a simulated season. For each *d* and country/region, we simulate *I* datasets (*i* = 1*, . . . , I* = 2000), of length *T* weeks each, where *T* is the country/region-specific number of weeks observed in the ground truth data. These simulations use the estimated 2022/23 proportions positive for influenza, *q_t,Flu,ILI_* = *y_t,Flu_/n_t,ILI_* or *q_t,Flu,ALL_* = *y_t,Flu_/n_t,ALL_*, suitably smoothed, as the assumed expected value (ground truth) *π_t,Flu,s_* from which to simulate. We use a moving average of three weeks to define the assumed ground truth values for the simulations, i.e. 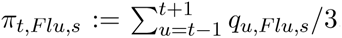, or a corresponding moving average of five weeks for weeks where the three-week period has zero observed positive tests.

Denote by *y_t,Flu,d,i_*the number of swabs positive for influenza (numerator) simulated for week *t*, sample size *d* in dataset *i*, drawn from a Binomial distribution with size *n_ILI,d_* and ground truth probability *π_t,Flu,ILI_* :

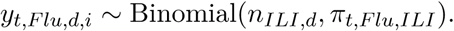

The *i*th simulated ILI+ metric is then defined as

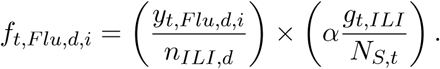

For the integrated multiple pathogen surveillance and ARI-based activity proxies in England, we instead jointly simulate the number testing positive for influenza, SARS-CoV-2, RSV and any other respiratory pathogen from a multinomial distribution:

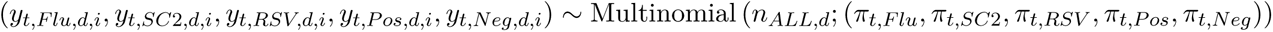

where *Neg* denotes negative for all pathogens tested and the denominator, *n_ALL,d_*, represents all swabs tested.

The *i*th simulated ARI+ metric *a_t,_*_+*,d,i*_ is then defined as the product of the ARI GP consultation rate and the proportion testing positive for at least one pathogen:

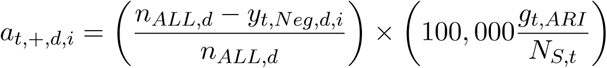

and the ARI-flu and ARI-SC2 metrics are defined analogously:

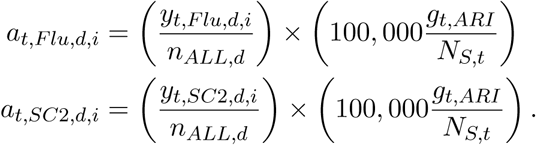

For both the ILI+ and ARI-based activity proxies, the 95% confidence interval for use in detecting Events 3 and 4 is obtained as the exact Clopper–Pearson binomial confidence interval (Clopper and Pearson 1934) for the product of the proportion positive and the respective consultation rate, since the consultation rates are considered known with certainty. For detecting events 7, 8, 10 and 12, the confidence intervals for the growth/decline rates in any metric considered are obtained using a standard profile likelihood method for generalised linear models (Venables and Ripley 2013). To approximate a realistic surveillance system the status of all events are re-evaluated based on the accumulated data every simulated epidemiological week. We assess the sample sizes and power to detect each event in Table 1 in terms of the two target quantities, the probability of detecting the event and time to detect the event. We summarise the distribution of the detection times over the *I* simulated datasets in terms of the mean, median, 2.5 and 97.5 percentiles. Full details of the mathematical expressions for detecting each event are given in Appendix C.

#### Software implementation

All analyses were implemented in R version 4.4.1 (2024-06-14), and the code is available on the git repository https://gitlab.developers.cam.ac.uk/amp62/gp-resp-surv-simulations.git.

## 3 Results

The simulated ILI+ data are shown in Figure D.1 in Appendix D.1. Throughout this section, we illustrate sample size determination using selected events. Results for all other events are given in Appendices D.2.1 to D.2.4. Note that detection times can be negative when an event in a simulated dataset occurs earlier than it did in the ground truth dataset used to simulate the data, given the stochasticity in the simulation, an indication of false-positive detections.

### 3.1 ILI+ activity

Detecting ILI+ crossing a threshold (Events 1 and 2, Figures 2) is straightforward, even at the smallest swabbing sample size of 25 tests per week, with 100% detection rates and mean detection times less than a week, although there is greater uncertainty in the detection times at smaller sample sizes than at larger ones. As the event to detect becomes more complex (being confident that the ILI+ activity rate has crossed a threshold, Event 3, and sustaining that confidence, Event 4, Figure D.2), we start to see smaller detection probabilities and longer detection times at smaller sample sizes, for Hong Kong. For example, for Event 4, Hong Kong requires a swabbing sample size of 250 a week to detect, with greater than 75% probability, significantly crossing a higher threshold for at least 2 consecutive weeks. As expected, crossing a lower threshold is easier to detect than the higher thresholds considered. Detection of Events 3 and 4 given the chosen thresholds remains straightforward for England and the USA.

**Figure 2:**
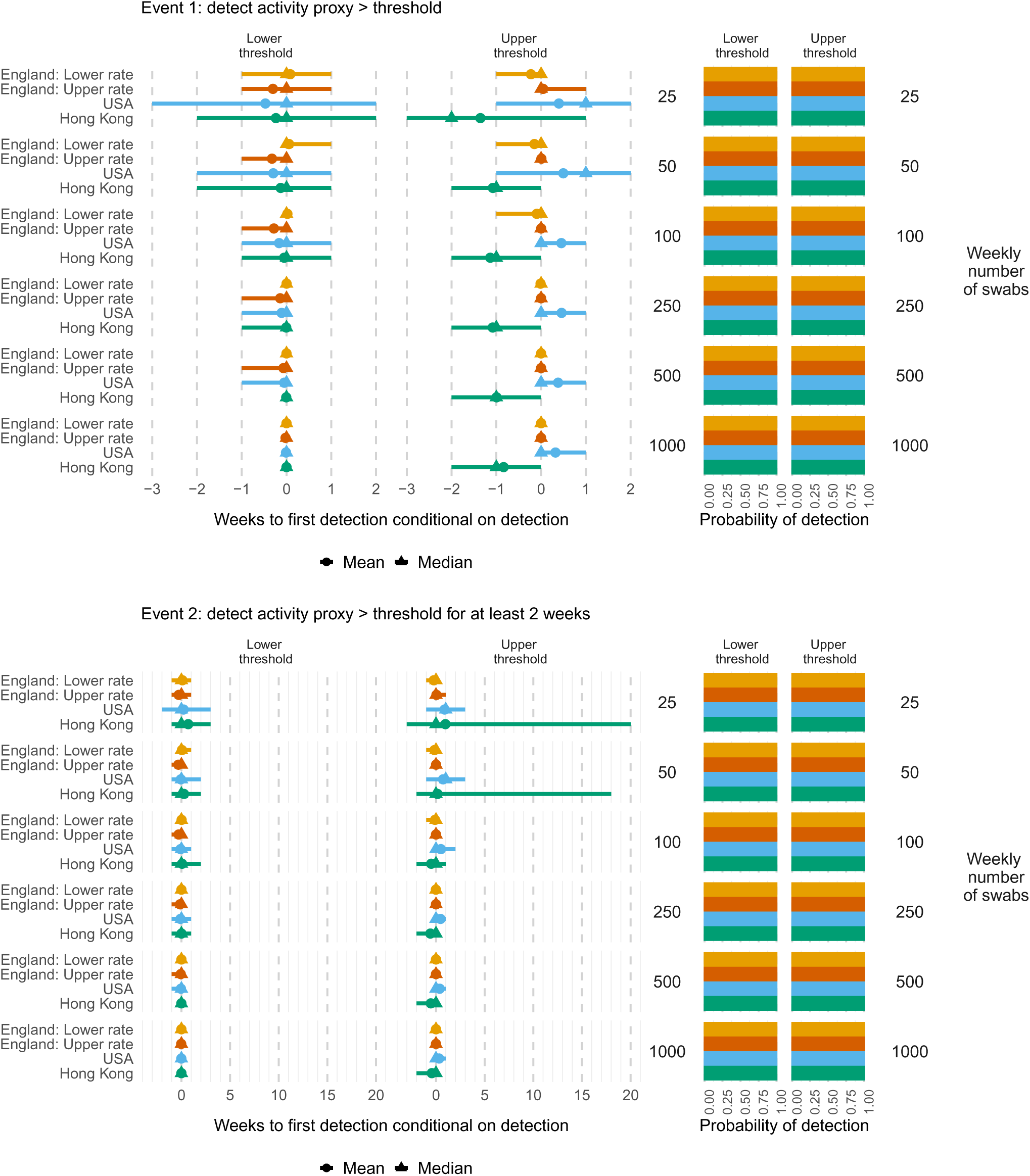
Times to (left) and probabilities of (right) detecting ILI+ activity greater than a country/region-specific lower or higher threshold (see Table B.1), by Event (1 top; 2 bottom), weekly number of swabs tested, threshold and country/region. Mean (circles), median (triangles) and 2.5 and 97.5 percentiles (line ranges) of times to detection are conditional on detection occurring. The probabilities of detection are obtained as the proportion of simulated datasets where the event occurs.

For Events 5 and 6, at least 3 consecutive weekly increases/decreases in ILI+ (Figure D.3), although the smallest sample size of 25 swabs a week tested is sufficient for 100% detection probabilities for England, 50 swabs a week are required to achieve greater than 75% detection rates for the USA and Hong Kong. For all three countries/regions, the detection times and their uncertainty reduce substantially with larger sample sizes. For example, for consecutive increases (Event 5), the mean (2.5 and 97.5 percentiles) detection times for England halve from 4 (0-8) weeks at 25 swabs per week to 2 (0-4) weeks at 1000 swabs per week. The reduction is even more dramatic for Hong Kong, from 17.5 (5-34) weeks to 2.5 (-2,6) weeks.

Detecting current growth/decline rates crossing thresholds (Events 9 and 11), based only on the last four weeks of data, is straightforward at all sample sizes and thresholds (Figure D.5). For detecting the corresponding Events 10 and 12 with significance (Figure 3), the detection probabilities increase and the detection times decrease with increasing sample size. Detecting a growth rate significantly larger than a high threshold (e.g. 1.1) is also clearly more challenging than for lower thresholds, and vice-versa for the decline rates. For England (yellow), a sample size of 50 swabs a week appears sufficient for any of the thresholds considered, with close to 100% detection probabilities and detection times less than a week. However, for the USA (blue) and Hong Kong (green), at least 250 swabs a week are needed to attain at least 75% detection probability that a current increase in ILI+ is detected above the country-specific threshold.

**Figure 3:**
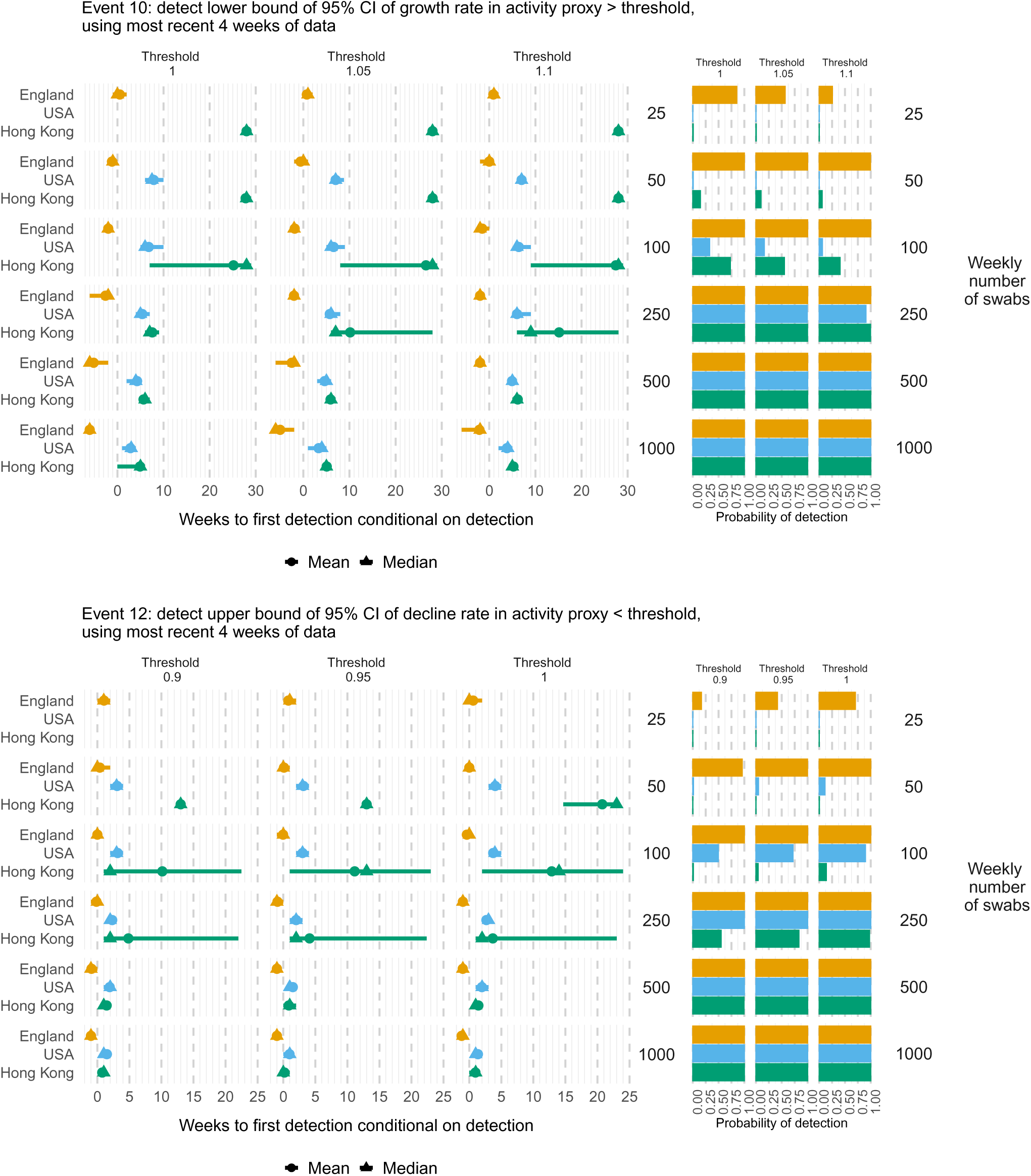
Times to (left) and probabilities of (right) detecting at a growth rate (top) or decline rate (bottom) in ILI+ activity significantly greater or smaller respectively than specified thresholds, based on the last four weeks of data, by weekly number of swabs tested and country/region. Mean (circles), median (triangles) and 2.5 and 97.5 percentiles (line ranges) of times to detection are conditional on detection occurring. The probabilities of detection are obtained as the proportion of simulated datasets where the event occurs. Note that when either the event does not occur in the ground truth data or in any of the simulated datasets, then a distribution of detection times is not available.

### 3.2 ARI-based activity

Here we compare sample sizes needed for sufficient detection for the ARI+ (consultation rates among those positive for at least one pathogen), ARI-flu (influenza-specific ARI consultation rates) and ARI-SC2 (SARS-CoV-2-specific ARI consultation rates) activity metrics.

Figure 4 demonstrates that for the thresholds chosen (Table B.1), any considered sample size is sufficient for detecting ARI+ or ARI-flu activity significantly greater than any threshold considered (Event 3), for at least two weeks (Event 4), with over 75% probability and in reasonable times, although 50 or more swabs a week reduces the time and uncertainty in the time to detection. However, for the ARI-SC2 activity proxy, given the much lower and less peaked rate of ground truth SARS-CoV-2 activity in the 2022/23 season (Figure A.2), the event never occurred for the higher thresholds in either the ground truth data or in any of the simulated datasets. For the lower thresholds, the familiar pattern of increasing detection probability and decreasing detection time with increasing sample size is observed, with 250 swabs a week required to attain at least 75% detection probability.

**Figure 4:**
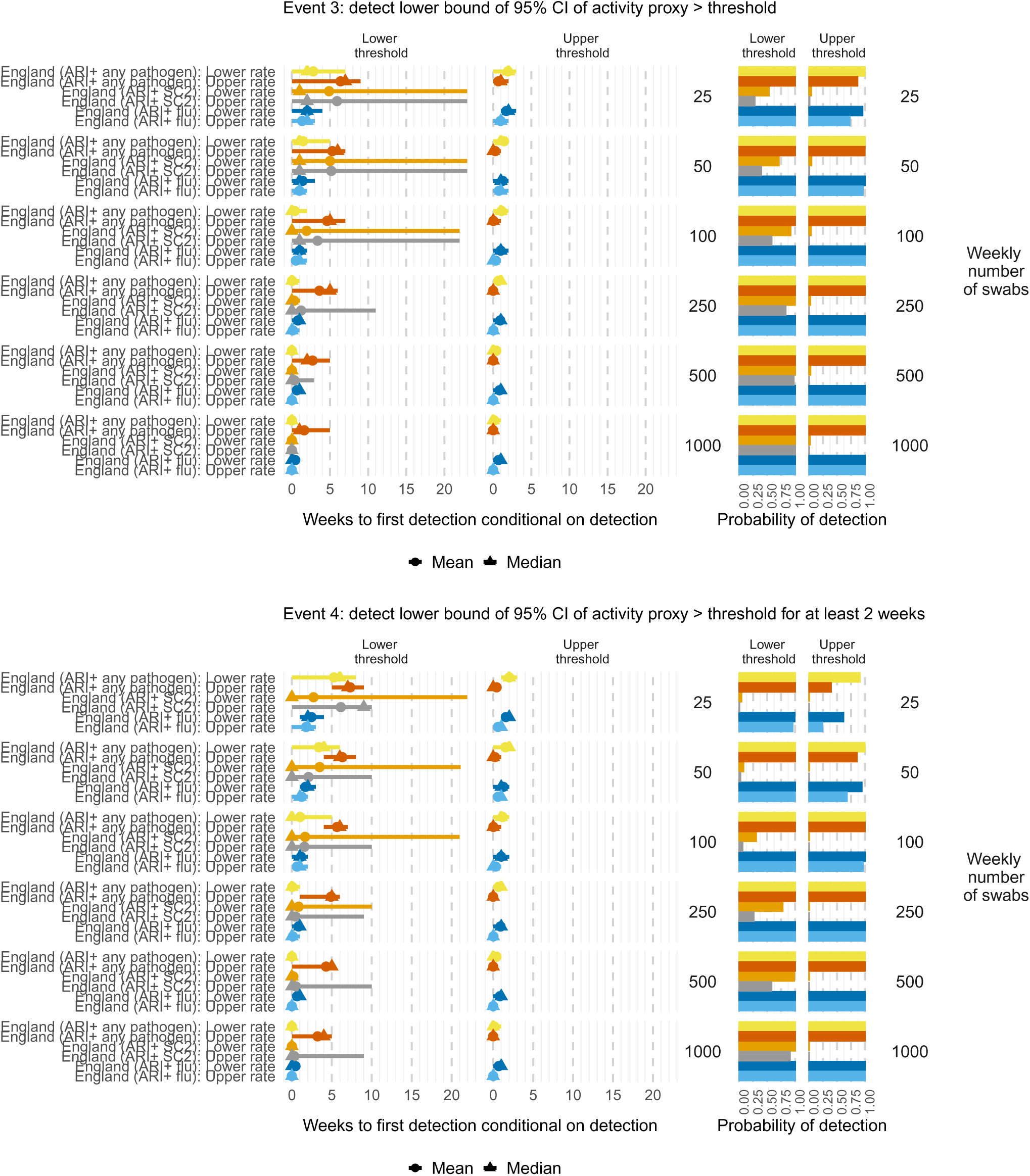
Times to (left) and probabilities of (right) detecting ARI-based activity metrics significantly greater (top), for at least two weeks (bottom), than lower or higher thresholds (Table B.1), by metric, weekly number of swabs tested and threshold. Mean (circles), median (triangles) and 2.5 and 97.5 percentiles (line ranges) of times to detection are conditional on detection occurring. The probabilities of detection are obtained as the proportion of simulated datasets where the event occurs.

Similar patterns are seen with other events for the ARI-based activity metrics. For example, Figure D.8 shows that detecting any growth rate (threshold 1) or 5% growth (threshold 1.05) or 10% growth (threshold 1.1) in the ARI-flu activity rate (Event 7) is straightforward at any sample size, with 100% detection probabilities and detection times for threshold 1.05 decreasing from a median of 2 weeks at 25 swabs a week to 0 weeks at 250 swabs a week. A sample size of at least 250 per week is needed to detect ARI+ growing at greater than 10% (threshold 1.1) with more than 75% probability; whereas 25 swabs is sufficient for detecting ARI+ growing at more than 5% rate with *>* 75% probability. For ARI-SC2, the event does not occur for any of the three thresholds, again due to the lack of signal in the ground truth data. In contrast, significant declines in ARI-SC2 (Event 8, Figure D.8) are detectable with reasonable power from 100 samples a week and up, but with a 5% decline requiring at least 1, 000 samples a week for more than 75% detection probability.

Figure 5 shows that detecting current (last 4 weeks) growth rates in the ARI-flu activity rate (green) significantly greater than 1 requires at least 100 swabs a week to attain more than 75% probability of detection and detection times less than 2 weeks; and 250 swabs a week for detecting current growth greater than 10% (threshold 1.1). A sample size of at least 500 per week is needed to detect ARI+ (yellow) growing at significantly greater than 10% with more than 75% probability; whereas 250 swabs is sufficient for detecting ARI+ growing at more than 5% rate with *>* 75% probability. For ARI-SC2 (blue), significant current growth is not detected at any sample size considered, with Event 10 not occurring for any of the three thresholds. In contrast, significant current decline of 10% (threshold 0.9) in ARI-SC2 (blue) is detectable with more than 75% probability, but only at 1,000 swabs per week.

**Figure 5:**
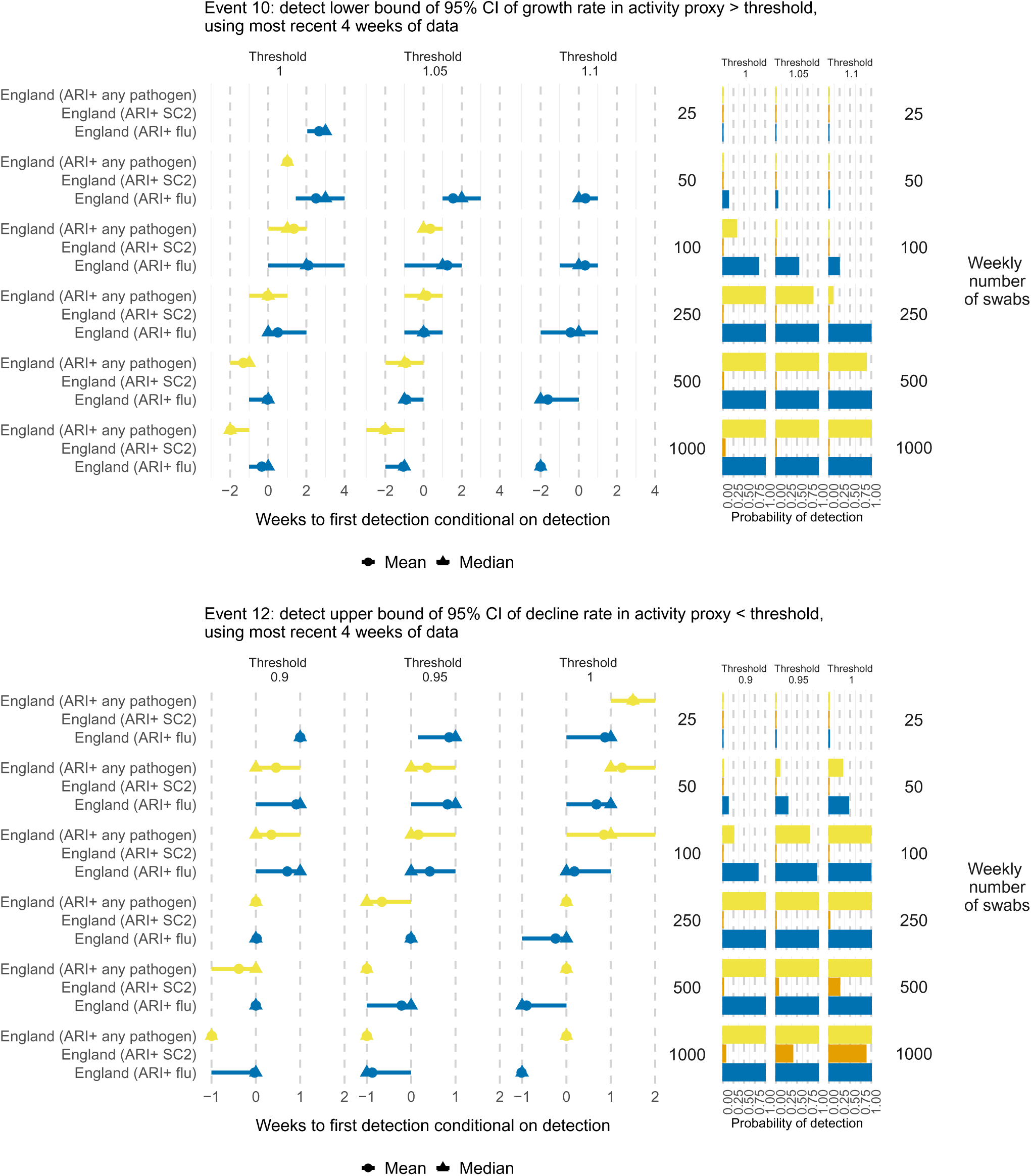
Times to (left) and probabilities of (right) detecting at a growth rate in ARI-based activity metrics significantly greater (top) or smaller (bottom) than specified thresholds, based on the last four weeks of data, by weekly number of swabs tested and country/region. Mean (circles), median (triangles) and 2.5 and 97.5 percentiles (line ranges) of times to detection are conditional on detection occurring. The probabilities of detection are obtained as the proportion of simulated datasets where the event occurs.

### 3.3 Comparing ARI-flu and ILI+ activity

Given the difference in scale between the ILI+ and ARI-flu ground truth data (Figure A.4), fair comparison of the power and time to detect events is challenging, because using common thresholds for the threshold-based Events 1-4 could result in either all simulations not experiencing crossing the thresholds or the ground truth data not crossing the threshold. The results presented here are therefore for different thresholds depending on whether ARI or ILI is used as the consultation rate (Table B.1). For a comparison for Events 1-4 using one common threshold (0.25) for the proportion positive for influenza out of either ILI or ARI/all swabs tested, see Appendix D.3.2.

For both the ILI+ and ARI-flu activity proxies for influenza circulation, Events 1, 2, 7, 9 and 11 (Figures D.10, D.13 and D.15) are equally straightforward to detect (bearing in mind the different thresholds used), with mostly 100% detection probabilities and short detection times at any sample size. However, the other Events had lower detection probabilities when using the ARI-flu metric than when using the classic ILI+ metric, assuming the same sample sizes. For example, Figure D.11 shows lower detection probabilities and longer detection times for ARI-flu compared to ILI+ for significant crossing of thresholds (Events 3 and 4). These differences likely reflect a lower specificity for influenza of the broader ARI symptom profile, thus requiring higher total number swabbed to enable observation of sufficient numbers of influenza cases.

Figure 6 demonstrates that while significant current growth or decline (Events 10 and 12) in the ILI+ metric (yellow) is detectable with over 75% probability at almost any sample size apart from the lowest 25 per week, the ARI-flu metric (blue) requires sample sizes of at least 100 a week to detect crossing the least extreme thresholds, and 250 a week to detect greater growth or decline. As expected, detection times reduce with increasing sample size for both metrics.

**Figure 6:**
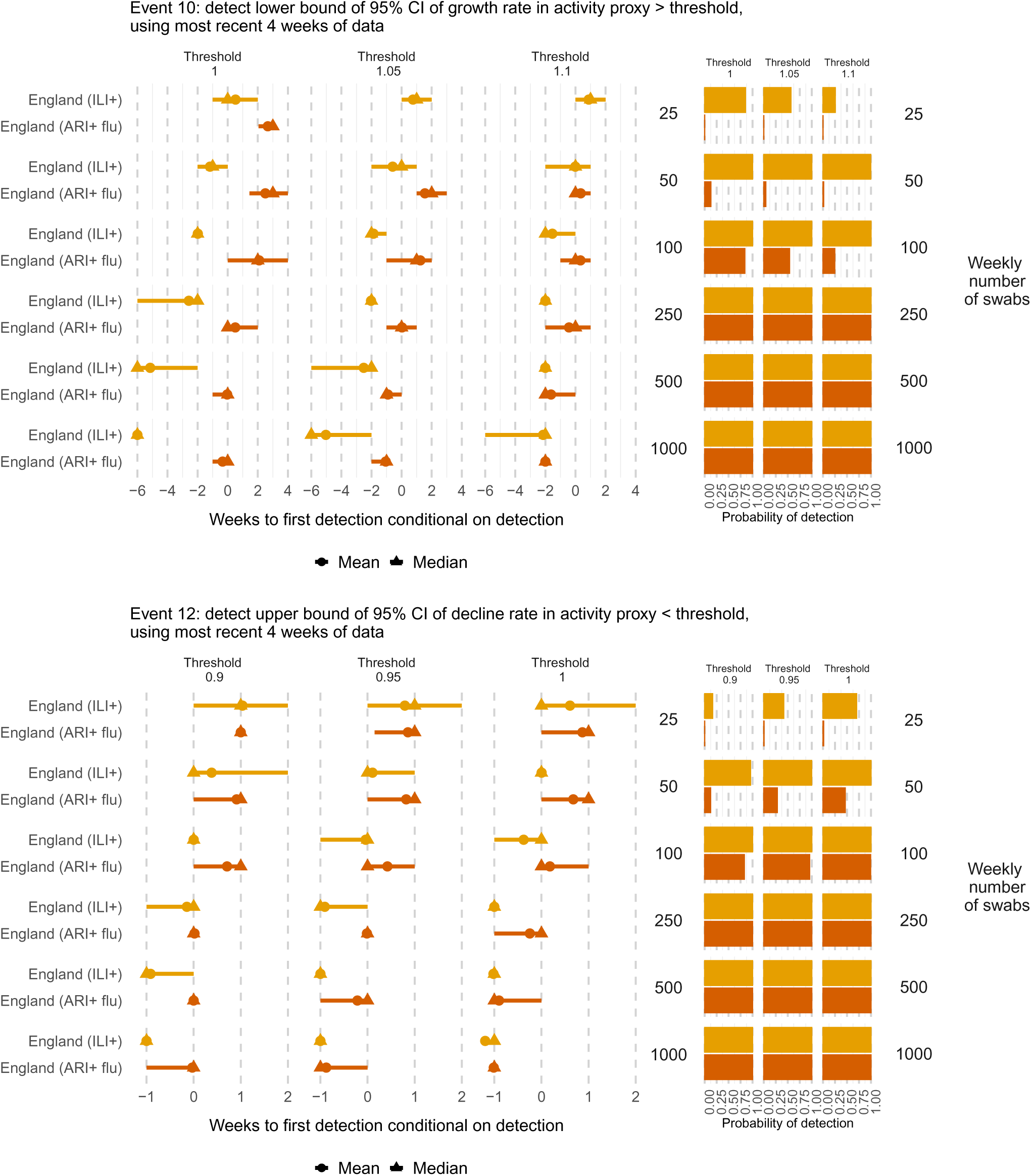
Times to (left) and probabilities of (right) detecting at a growth rate (top) or decline rate (bottom) in ILI+ activity significantly greater or smaller respectively than specified thresholds, based on the last four weeks of data, by weekly number of swabs tested and country/region. Mean (circles), median (triangles) and 2.5 and 97.5 percentiles (line ranges) of times to detection are conditional on detection occurring. The probabilities of detection are obtained as the proportion of simulated datasets where the event occurs.

### 3.4 Multiple pathogens integrated analyses

Figure 7 demonstrates that for detecting current growth/decline rates in the proportion positive that are significantly different from 1 in at least one pathogen, based on the most recent four weeks of data, sample sizes of at least 250 a week are needed to achieve *>* 75% detection probabilities. For detecting more extreme growth rates (thresholds 1.05 and 1.1), at least 500 or 1000 swabs a week are required. Influenza is almost always the first pathogen where these events are detected, when the event occurs at all in the simulated datasets.

**Figure 7:**
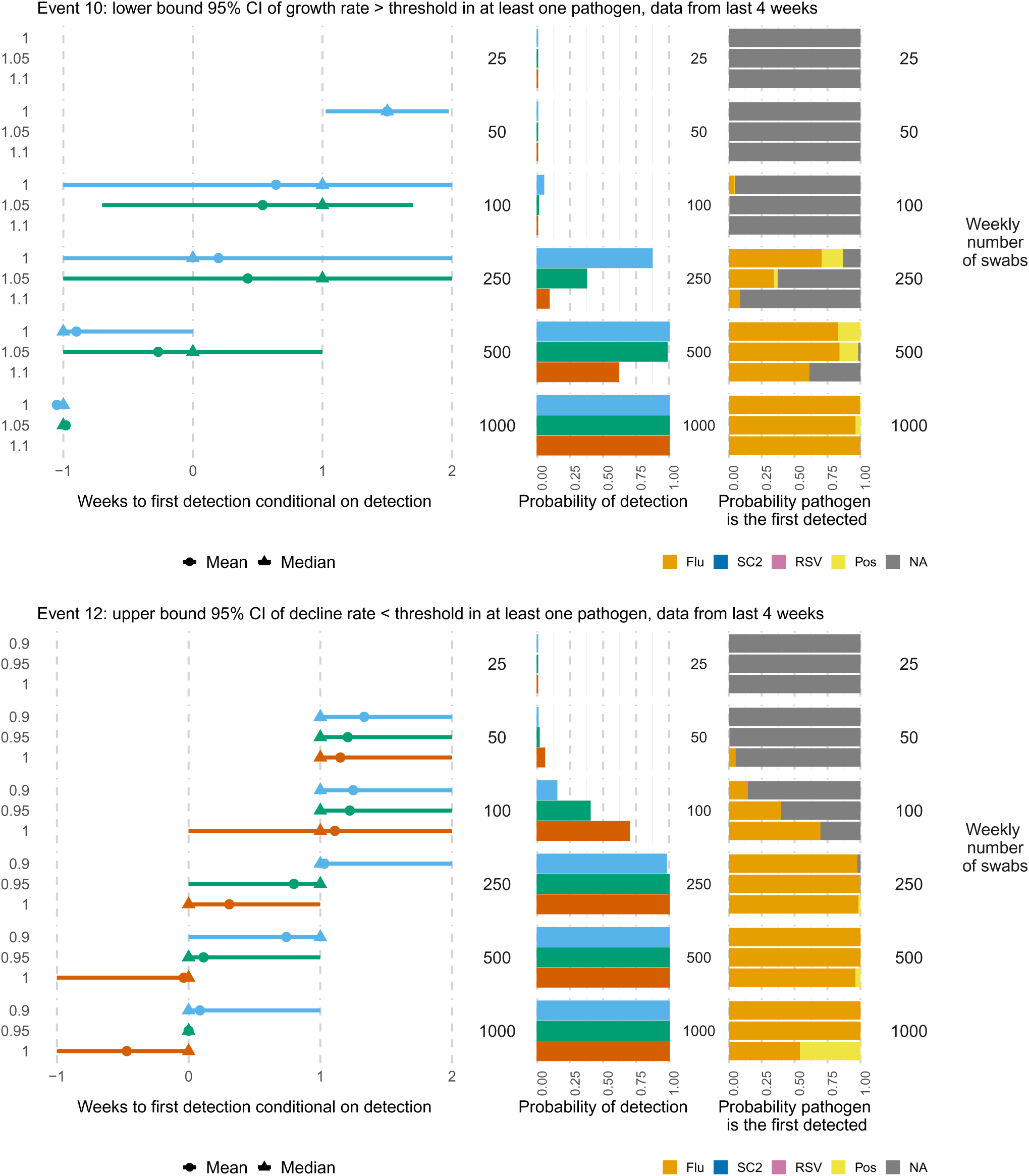
Times to (left) and probabilities of (middle) detecting at a growth rate (top) or decline rate (bottom) in proportion positive significantly greater/less than specified thresholds in at least one pathogen, based on the last four weeks of data, by weekly number of swabs tested. The right-hand panel shows the probability each pathogen is the first one detected, where Flu denotes influenza, SC2 denotes SARS-CoV-2, RSV denotes respiratory syncytial virus and *Pos* refers to positive for any other tested virus other than Flu, SC2 and RSV. NA denotes missing information due to the event not occurring. Mean (circles), median (triangles) and 2.5 and 97.5 percentiles (line ranges) of times to detection are conditional on detection occurring. The probabilities of detection are obtained as the proportion of simulated datasets where the event occurs. Note that when either the event does not occur in the ground truth data or in any of the simulated datasets, then a distribution of detection times is not available.

### 3.5 Comparison to actual sample sizes

Figure A.5 in Appendix A demonstrates that for the USA and Hong Kong, the sample sizes for swabs taken for testing across each country/region each week during the seasons of interest were well above the level needed for adequate detection probabilities and times to detection for each of the events assessed. Note that if considering sub-populations, e.g. sub-regions (states or districts) or age groups, the observed sample sizes will have been smaller. For England, the ILI swabs tested in the 2022/23 seasons considered in isolation may not have been adequate in every week for detecting all events for the ILI+ metric with sufficient probability and reasonable times, particularly at the start and end of the season and when considering sub-groups; however, the revised criteria for swabbing ARI cases is likely to have allowed a sufficient total number of swabs tested for adequate detection of most events for both ILI+ (if using all swabs tested, regardless of symptom profile as the denominator for the proportion positive for influenza in defining ILI+) and for the ARI-based activity proxies.

### 3.6 Supplementary analyses

Detection summaries for the proportion positive metric for influenza alone are shown in Appendix D.3.1. Results using the alternative all swabs (rather than ILI) denominator for the proportion positive for influenza in England are shown in Appendix D.3.2. The assessments of the probabilities of and times to detection show that detecting the proportion positive crossing the common threshold (Events 1-4) of 0.25 (lower threshold for the ILI denominator, higher threshold for the ARI/all swabs denominator) is more challenging using the larger denominator than the smaller ILI denominator, due to the smaller proportions positive.

## 4 Discussion

We have demonstrated how to define a range of different events to detect, fulfilling various surveillance objectives, including epidemic detection, situational awareness and intensity evaluation for respiratory viruses. We have shown how to choose sample sizes based on comparing two target quantities under different numbers of swabs to test, the probability of detection (power) and time to detection of an event. We have considered three different epidemic patterns based on real-world data from which to simulate, with our example scenarios including epidemics in both temperate and non-temperate climates mimicking the 2022/23 season or 2023 year. We have used a range of metrics, including the ILI+ influenza activity metric, ARI-based activity metrics and the integrated metric of proportions positive for multiple pathogens as illustration, demonstrating how to determine sample sizes for detecting events to achieve integrated surveillance objectives, such as detecting a positive trend in at least one of several monitored pathogens.

As might be expected, we found that more complex events, with definitions that required more information (e.g. events defined by confidence intervals crossing thresholds compared to those defined by point estimates crossing thresholds), require larger sample sizes than simpler events. We confirmed that for events involving comparison of a metric to a threshold, perhaps to evaluate the “intensity” of a wave of infection compared to a historical baseline, sample sizes required to achieve a particular power (probabilities of detection) varied depending on the level of the threshold, as expected. Although the power, detection times and corresponding sample sizes depend on threshold sizes, an advantage of the approach is that sample sizes can be chosen to ensure accurate and timely detection of threshold-based events where the thresholds have been chosen to represent meaningful expected trends and changes in the metrics used to monitor circulating viruses.

For the ILI+ activity rate metric, probabilities of detection were generally large, even at small sample sizes, but larger sample sizes were needed to have a substantive effect on reducing times to detection of some events, for example for having confidence in large current growth or decline rates (Events 10 and 12, Figure 3). Similar patterns were seen for the ARI+ activity metric (a proxy for incidence of all respiratory pathogens tested), but with larger sample sizes needed for more complex events to be detected with greater than 75% probability and with shorter, more certain, detection times. In contrast, it was easier to detect events adequately for the ARI-flu activity proxy than for ARI+, even at low sample sizes. However, when comparing the ARI-flu activity proxy to the more common ILI+ activity proxy, some events concerning ILI+ were easier to detect than for ARI-flu. This might be explained by the “peakier” shape of the 2022/23 ground truth ILI+ compared to ARI-flu activity rates when plotted on the log-scale (Figure A.4, Appendix A), likely reflecting higher specificity for influenza of ILI than the broader ARI symptom profile. However, the very different scales of the two metrics required setting different magnitudes of thresholds for the threshold-based events to detect, and setting these to be comparable across metrics is challenging. For the ARI-SC2 activity metric, the low signal provided by a ground truth metric that did not display substantial increases or decreases over the 2022/23 season meant that in general, larger sample sizes were required to detect events with reasonable power and in short enough times. However, declines were easier to detect than increases, and the results nevertheless indicate the potential for using ARI as a more suitable base than ILI in activity proxies for monitoring SARS-CoV-2 circulation, in the interest of integrated surveillance. When considering the integrated analysis of proportions positive for at least one tested pathogen, detection probabilities were generally high at all sample sizes for the less complex events (e.g., comparing a metric to a baseline threshold), but detection times reduced both in terms of the mean times and in terms of uncertainty with increasing sample size. More complex events such as confidence in the current growth/decline rate detections (Events 10 and 12, Figure 7) required sample sizes of at least 250 a week to detect them with high power and within reasonable times.

When considering the proportion positive metric on its own for a single pathogen such as influenza (Appendix D.3.1), detection probabilities increased with sample sizes, requiring numbers of weekly swabs tested to be at least 100 to achieve 75% detection probabilities for peaked epidemics such as those experienced by England and the US in 2022/23 (see also the revised GISRS integrated surveillance guidance, World Health Organization 2024a, who suggest 50 samples per week at a minimum, but with 150 samples recommended for more precise monitoring of proportion positive). However, for countries/regions with less clear epidemics, such as Hong Kong, usually at least 250 samples a week are needed for 75% power. These findings suggest that combining the proportion positive data with the much larger ILI or ARI consultation rates into activity proxies allows sufficient detection of events of interest even at low virology testing sample sizes.

When comparing required sample sizes for large probabilities of and small times to detection to the actual sample sizes observed during the 2022/23 or 2023 seasons used, we found that at least at national level, the surveillance systems for England, the USA and Hong Kong are sufficient for adequate detection of each event for both the ILI+ activity metric and the proportion positive for influenza, particularly if using the broader ARI criteria for selection of swabbing for England. However, when restricting to samples collected due to ILI criteria in the 2022/23 season in England, weekly sample sizes were mostly smaller than 100, apart from at the peak. These numbers of ILI swabs would not have been sufficient to detect some events with high probability or in a reasonable time: for example, detecting a growth rate significantly greater than 10% in the ILI+ activity proxy based on data in the last four weeks (Figure 3).

In summary, these results suggest that the ARI+ metric might be useful, e.g. for an initial indication of respiratory activity, even when using relatively low sample sizes. However, for effective integrated pathogen-specific monitoring using ILI+ and the pathogen-specific ARI metrics, we would require larger sample sizes of at least 250 a week, unless an outbreak is particularly large.

Although our results are specific to the countries/regions considered and the context of 2022/2023 circulation, the approach can be generalised to (a) different country and climate contexts; (b) more complex surveillance objectives and events, such as estimating incidence of infection, case- or infection-severity risks and vaccine effectiveness; (c) more sophisticated sampling models than the independent binomial and multinomial sampling distributions used here, accounting e.g. for over-dispersion and clustering; and (d) trading off multiple surveillance objectives against logistical and cost constraints, e.g. understanding whether to change the number of sentinel sites in a surveillance system. For (a), other countries could adopt the approach taken here for their specific country and sub-national areas; or the exercise could be repeated for a range of different example season/epidemic types (e.g. low incidence, fast-growing epidemics, slow-growing epidemics, etc.), and countries could match the types of epidemics they see to these examples to decide on sample sizes suitable for their context. For (b), generalising the approach would entail formulating mathematically the objectives, such as estimation or prediction, in order to rightsize surveillance to achieve them. The extension in (c) is an approach proposed by Cheng et al. 2020, who suggest that surveillance design requires four components: (i) understanding of the disease process; (ii) understanding of the surveillance process, including the objectives of surveillance and the data available to achieve these objectives; (iii) a mathematical definition of target quantities (“objective functions”) to optimise (as also advocated by Fearon et al. 2026); and (iv) a method to perform that optimisation, possibly subject to logistical and/or cost constraints. Here we have taken the first steps towards this approach, concentrating on steps (ii) and (iii), and proposing how to define multiple targets and events to detect for multiple surveillance objectives. We have taken a simulation approach to comparing and choosing sample sizes for detecting each of these events and objectives. However, a next step for future work will be to formally optimise multiple target quantities for multiple events and objectives *simultaneously*, moving towards a trade-off approach where we can also optimise subject to constraints.

Our illustrative examples here have used country/region-specific thresholds based either on historical data, using the Moving Epidemic Method (Vega et al. 2013), its variations, standard deviation-based approaches or expert knowledge to determine thresholds (Table B.1, Appendix B). During the SARS-CoV-2 pandemic, both non-SARS-CoV-2 respiratory pathogen circulation and monitoring of metrics such as ILI and ARI rates and the composite ILI+ activity proxy were substantially disrupted by the pandemic and its associated public health and social measures (Cobb et al. 2022; Fricke et al. 2021; Grosso et al. 2021; Zhang et al. 2023). Further work is therefore needed on how to set thresholds based on historical data for the ILI, ILI+ and SARS-CoV-2 metrics (Sinnathamby et al. 2024), and for alternative metrics such as the new ARI-based metrics we have proposed in England (Elson et al. 2024; Gu et al. 2024).

A key consideration is the challenging interpretation of the classic metrics used for monitoring respiratory pathogen circulation, particularly the proportion positive, given time-varying testing strategies and co-circulation (Eales et al. 2023), highlighting the need for integrated surveillance, the assessment of objectives in the context of multiple pathogens as we have illustrated, and more widespread adoption of the ILI+ and ARI-based activity metrics (Eales et al. 2024). Constructive further research would include assessing the representativeness of sentinel surveillance systems and the feasibility of random sampling, accounting for time-varying sampling and co-circulation, assessing how well ARI-based activity metrics approximate incidence, and estimating multi-pathogen incidence in an integrated surveillance framework.

Our examples have used the influenza activity proxies ILI+ and ARI-flu and one for SARS-CoV-2 (ARI-SC2), but sample size determination for activity proxies for other pathogens such as RSV could also be carried out with some careful choice of the underlying consultation rate to use. An activity proxy for RSV is typically defined as RSV positivity among bronchitis or bronchiolitis cases in children aged *<* 5 years old times the consultation rate for bronchitis/bronchiolitis in the same age group (Lusignan et al. 2025; Reeves et al. 2019). However, in some countries or regions, the challenge of obtaining swab samples from young children may limit the precision of the proportion positive, further emphasising the need to balance surveillance objectives with logistical constraints. For SARS-CoV-2, further work is needed to understand whether ARI consultations or a more precise subcategory based on clinical indication of lower respiratory tract infection (LRTI) may be more suitable for an activity proxy. When considering integrated surveillance of all respiratory pathogens tested, we did not consider an activity proxy given the potential need for different consultation denominators for different pathogens.

Nevertheless, the consideration of multiple surveillance objectives, activity metrics, events and pathogens demonstrated here is an important step toward an efficient integrated respiratory pathogen surveillance system.

## Data Availability

The simulated data that support the findings of this study are openly available in the repository gp-resp-surv-simulations at https://gitlab.developers.cam.ac.uk/amp62/gp-resp-surv-simulations.git. Aggregate data used to simulate from are publicly available as detailed in Section 2.1. Individual-level pseudo-anonymised data underlying these aggregate data are not publicly available, see Section 2.1.

## Acknowledgments

The authors are grateful to patients, general practices, Magentus (formerly Wellbeing) and Optum (formerly EMIS) for sharing pseudonymised data with the Royal College of General Practitioners Research and Surveillance Centre. We also thank colleagues at the University of Oxford, UK Health Security Agency and Royal College of General Practitioners involved in obtaining and/or processing primary care and virology swab data. We thank Aspen Hammond and Kaat Vandemaele from the Global Influenza Programme at WHO and members of the WHO technical working group for the GISRS revised integrated surveillance guidance for useful discussions and feedback.

## Funding

AMP, TN and DDA were funded by the UK Medical Research Council programme MC_UU_00040/04. SSV was funded by the UK Medical Research Council programme MC_UU_00040/03

## Glossary

**Table 2.** summarises notation and acronyms used in this paper. Notation and acronyms used throughout the paper

| Notation | Description |
| --- | --- |
| ILI | Influenza-Like Illness |
| ARI | Acute Respiratory Infection |
| Flu | Influenza |
| SC2 | SARS-CoV-2 |
| RSV | Respiratory syncytial Virus |
| Pos | positive for any other pathogen tested (other coronaviruses, human metapneumovirus, adenovirus, enterovirus, human rhinovirus) |
| Neg | negative for all pathogens tested |
| $c$ | Indexes country/region/subpopulation |
| $t$ | Indexes time/week |
| $s$ | Indexes symptom profile/consultation type $s = \text{ILI}$ or $s = \text{ARI}$ or $s = \text{ALL}$ |
| $p$ | Indexes pathogen |
| $P$ | Denotes population |
| $S$ | Denotes sentinel surveillance |
| $N_{P,t}$ | Total population size at week $t$ |
| $g_{P,t,s}$ | Number of GP or outpatient consultations in the population at week $t$ for symptom profile $s$ |
| $N_{S,t}$ | Population size covered by sentinel surveillance at week $t$ |
| $g_{S,t,s}$ | Number of GP or outpatient consultations at week $t$ observed in sentinel surveillance for symptom profile $s$ |
| $n_{t,s}$ | Number of swabs taken in week $t$ from consultations for symptom profile $s$ |
| $y_{t,p}$ | Number of swabs testing positive for pathogen $p$ in week $t$ |
| ILI+ | influenza activity rate proxy |
| ARI+ | ARI-based activity rate proxy for at least one tested pathogen |
| ARI-flu | ARI-based influenza activity rate proxy |
| ARI-SC2 | ARI-based SARS-CoV-2 activity rate proxy |
| CI | confidence interval |
| $\alpha$ | country/region-specific scale for the consultation rates (100,000; 100; 1,000 for England, USA & Hong Kong respectively) |
| $d$ | Indexes the sample sizes $n_{t,s} \in \{25, 50, 100, 250, 500, 1000\}$ |
| $I$ | Number of simulated surveillance datasets per country/region and sample size |
| $T$ | Number of surveillance weeks simulated |
| $q_{t,p,s}$ | unsmoothed observed proportion positive for pathogen $p$ in week $t$ among swabbed consultations with symptom profile $s$ |
| $\pi_{t,p,s}$ | smoothed proportion positive for pathogen $p$ in week $t$ among swabbed consultations with symptom profile $s$, used as ground truth |
| $y_{t,p,d,i}$ | number of tests positive for pathogen $p$ in week $t$ in the $i$'th simulated dataset under sample size $d$ |
| $f_{t,Flu,d,i}$ | ILI+ value in week $t$ in the $i$'th simulated dataset under sample size $d$ |
| $a_{t,p,d,i}$ | ARI-based activity proxy value in week $t$ in the $i$'th simulated dataset under sample size $d$ ($p$ is either + for ARI+ or *Flu* or *SC2* for the pathogen-specific proxies) |

## Appendices

### A Ground truth surveillance data

ILI and ARI consultation rates, on both the original and log scale, are shown in Figure A.1.

**Figure A.1:**
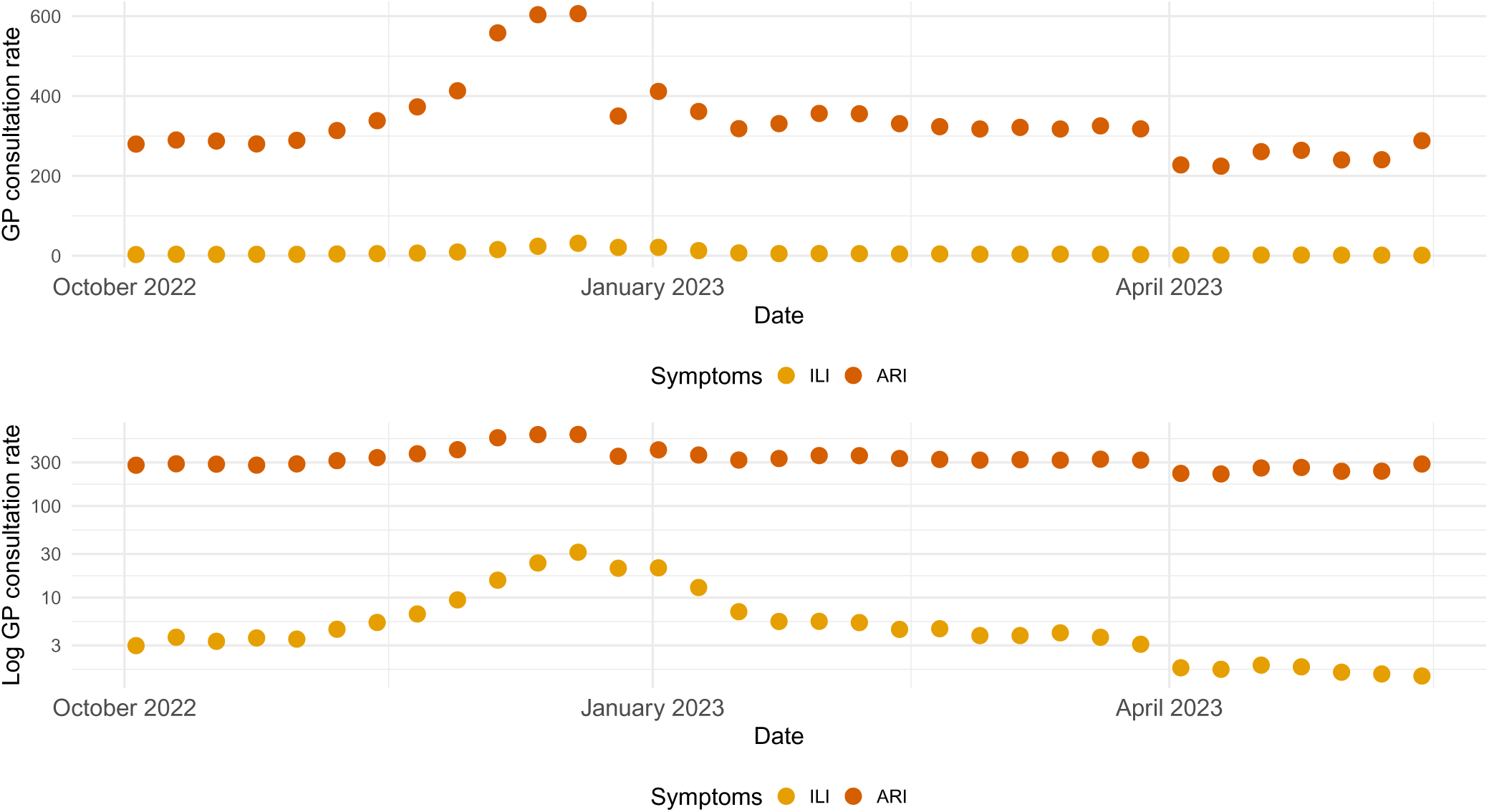
Consultation rates per 100,000 population in England, on original scale (top) or log scale (bottom), by symptom phenotype, ILI or ARI.

**Figure A.2:**
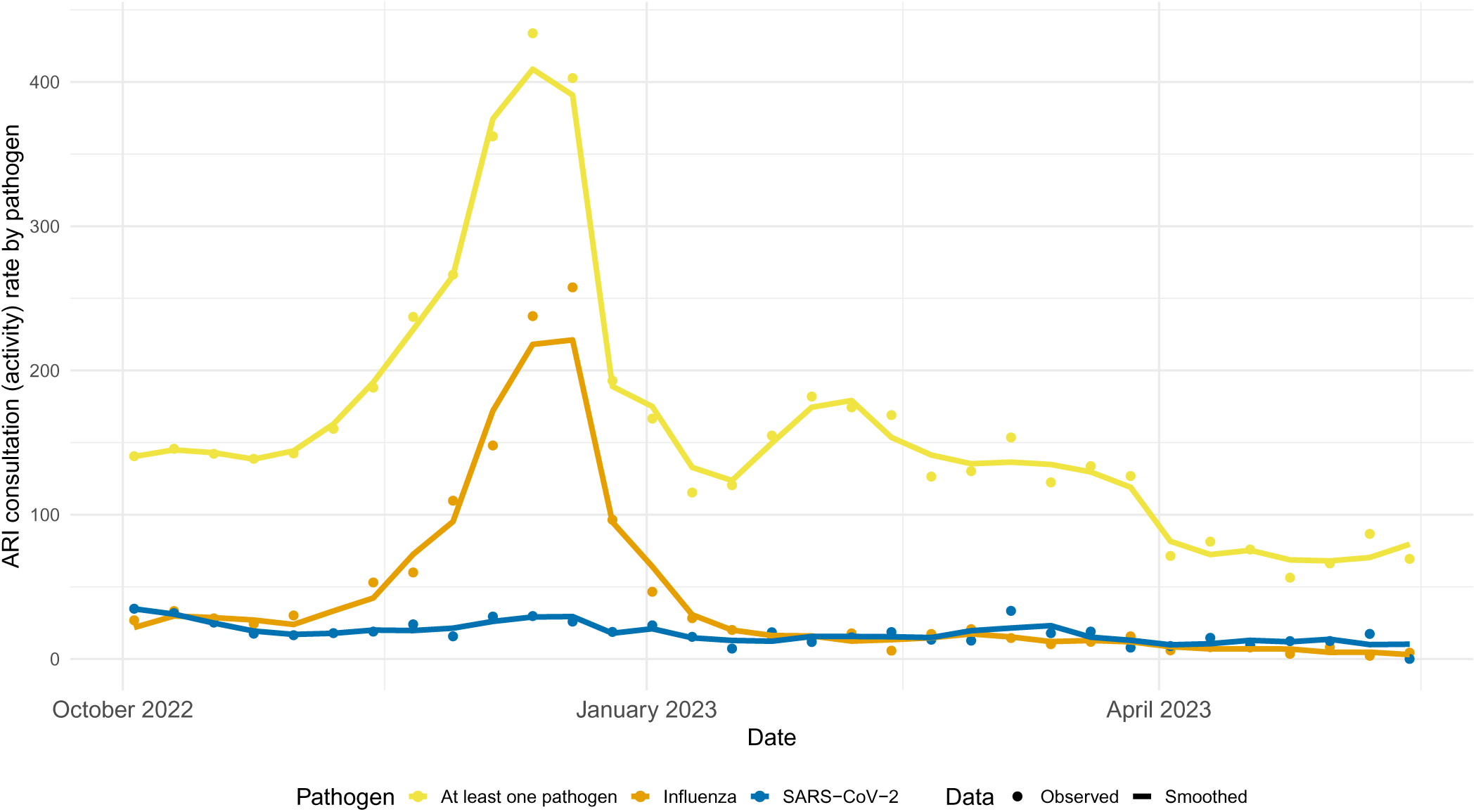
ARI-based activity proxies, by pathogen (at least one pathogen, influenza or SARS-CoV-2), in England, with the smoothed version using 3-week moving averages.

**Figure A.3:**
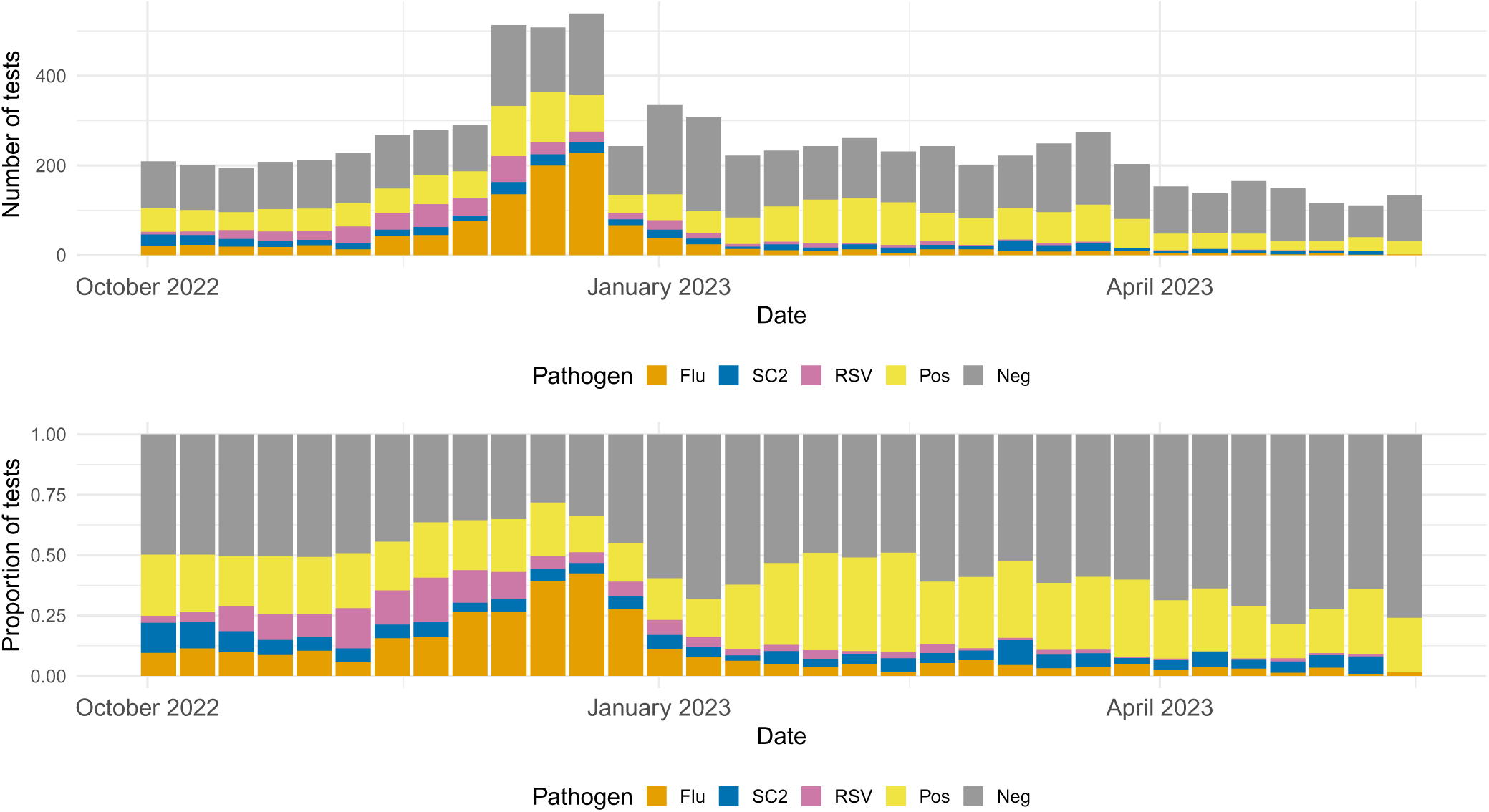
Numbers and proportions of tests by test results (positive for different pathogens or negative) in England, when using ARI/all swabs tested as a denominator.

**Figure A.4:**
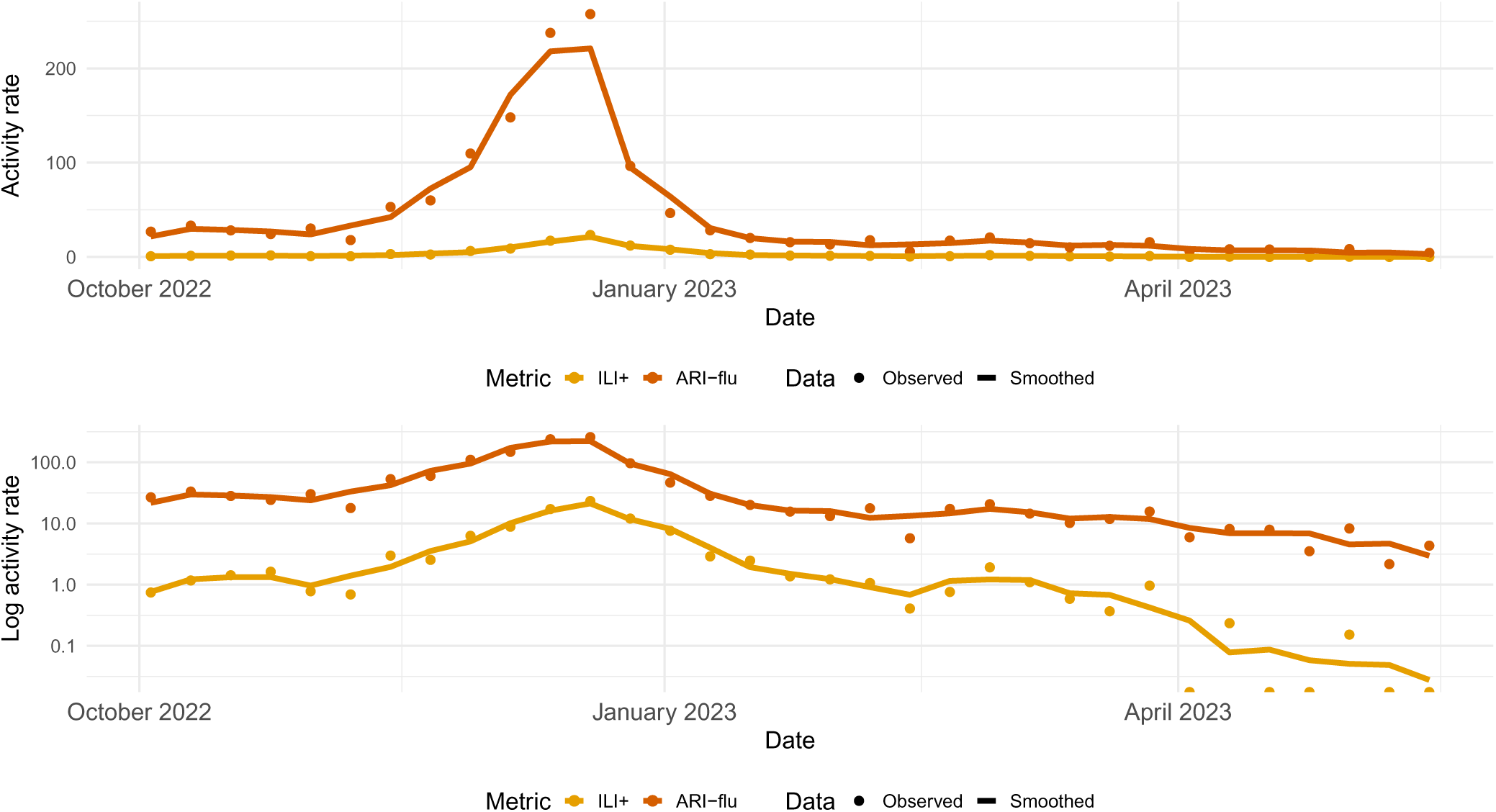
Influenza-specific consultation (activity) rates per 100,000 population in England, by symptom profile (ILI or ARI).

**Figure A.5:**
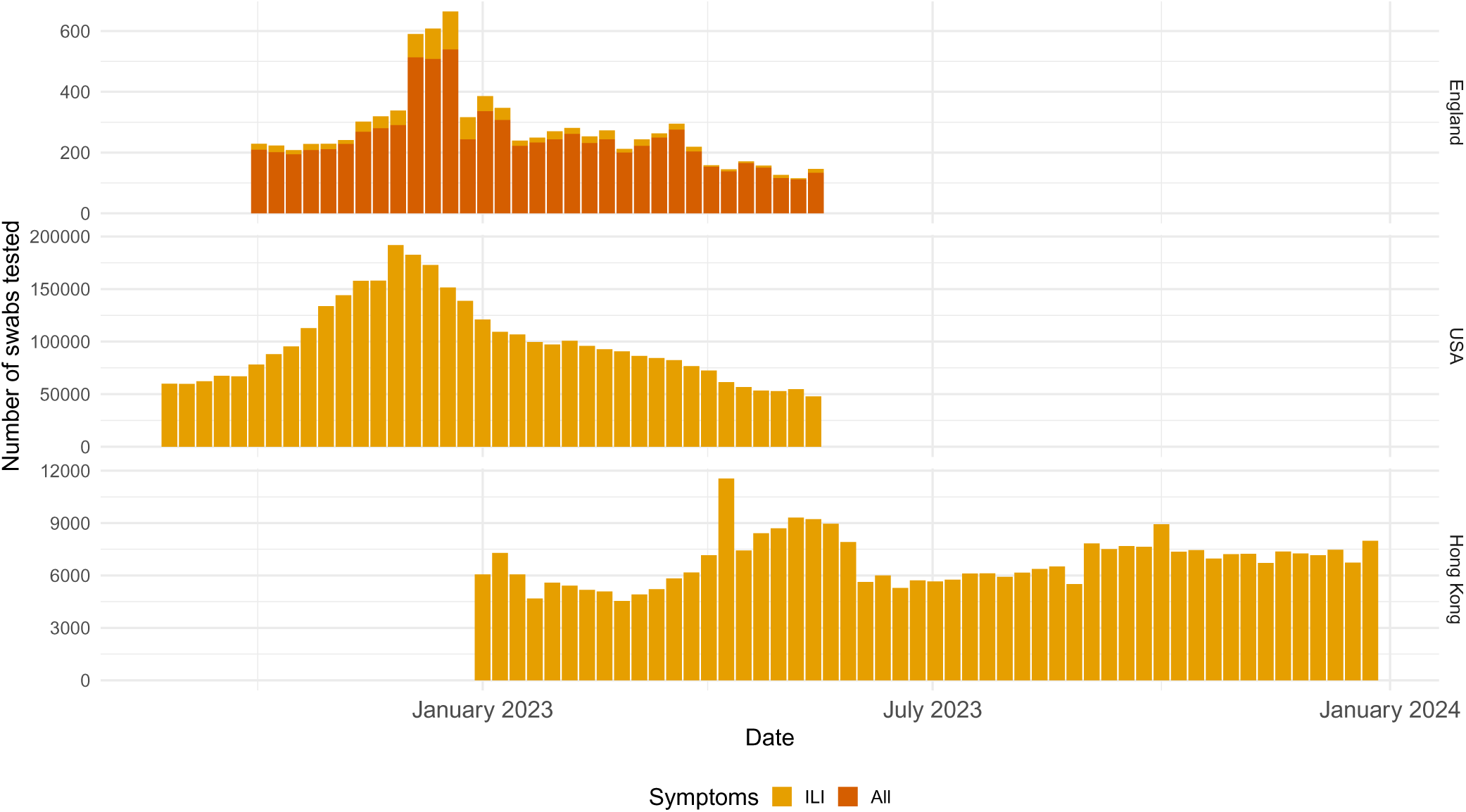
Weekly number of swabs tested, by country/region, and for England, by whether only ILI symptom-based swabs or all swabs were tested.

The ARI-based activity proxies are shown in Figure A.2.

Figure A.3 shows the distribution of test results when using ARI/all swabs tested as the denominator A comparison of the influenza activity proxies using either ILI or ARI consultation rates as the pool from which swabs are collected is shown in Figure A.4.

The numbers of swabs tested, based on either ILI or ARI/all consultations, are shown in Figure A.5.

### B Thresholds

The thresholds used in our simulation approach are given in Table B.1.

**Table B.1:** Thresholds by country/region and metric. Most of the metrics have two different surveillance thresholds specified.

| Country/region | Metric | Threshold |  | Source |
| --- | --- | --- | --- | --- |
|  |  | Lower | Upper |  |
| England | Proportion positive for influenza out of ILI swabs | 0.25 | 0.50 | Expert knowledge |
|  | ILI consultation rate (per 100,000 population) | 11.47 | 15.06 | MEM baseline and low thresholds |
|  | ILI+ activity rate using lower proportion positive for influenza | 2.87 | 3.77 | ILI rate * lower proportion positive |
|  | ILI+ activity rate using upper proportion positive for influenza | 5.74 | 7.53 | ILI rate * upper proportion positive |
|  | Proportion positive for influenza out of all swabs | 0.10 | 0.25 | Expert knowledge |
|  | ARI consultation rate (per 100,000 population) | 350.00 | 450.00 | Expert knowledge |
|  | ARI-influenza activity rate using lower proportion positive for influenza | 35.00 | 45.00 | ARI rate * lower proportion positive |
|  | ARI-influenza activity rate using upper proportion positive for influenza | 87.50 | 112.50 | ARI rate * upper proportion positive |
|  | Proportion positive for SARS-CoV-2 out of all swabs | 0.05 | 0.10 | Expert knowledge |
|  | ARI-SC2 activity rate using lower proportion positive for SC2 | 17.50 | 22.50 | ARI rate * lower proportion positive |
|  | ARI-SC2 activity rate using upper proportion positive for SC2 | 35.00 | 45.00 | ARI rate * upper proportion positive |
|  | Proportion positive for at least one pathogen out of all swabs | 0.30 | 0.60 | Expert knowledge |
|  | ARI-AL1 activity rate using lower proportion positive for at least one pathogen | 105.00 | 135.00 | ARI rate * lower proportion positive |
|  | ARI-AL1 activity rate using upper proportion positive for at least one pathogen | 210.00 | 270.00 | ARI rate * upper proportion positive |
| USA | Proportion positive for influenza out of ILI swabs | 0.10 | 0.15 | Expert knowledge |
|  | ILI consultation rate (per 100 outpatient consultations) | 2.50 |  | CDC 2-year historic baseline |
|  | ILI+ activity rate using lower proportion positive for influenza | 0.25 |  | ILI rate * lower proportion positive |
|  | ILI+ activity rate using upper proportion positive for influenza | 0.38 |  | ILI rate * upper proportion positive |
| Hong Kong | Proportion positive for influenza out of ILI swabs | 0.09 | 0.15 | CHP 5-year historic baseline and expert knowledge |
|  | ILI consultation rate (per 1,000 outpatient consultations) | 30.00 |  | Expert knowledge |
|  | ILI+ activity rate using lower proportion positive for influenza | 2.76 |  | ILI rate * lower proportion positive |
|  | ILI+ activity rate using upper proportion positive for influenza | 4.50 |  | ILI rate * upper proportion positive |

### C Mathematical definitions

Let *c* denote the country/region/subpopulation of interest. Table 2 summarises notation and key acronyms in the main text, Table C.1 summarises additional notation in this section.

**Table C.1:** Notation and acronyms used in appendices

| Notation | Description |
| --- | --- |
| $\mu_{c,t,d,i}$ | Generic notation for simulated value of a metric for country/region $c$ in week $t$ in the $i$'th simulated dataset under sample size $d$ |
| $\mu_{c,t}^{TRUE}$ | Equivalent generic notation for the true value of a metric for country/region $c$ in week $t$ |
| $\tau$ | threshold |
| $T_{c,e}^{TRUE}$ | True event time for event $e$ in country/region $c$ |
| $T_{c,e,d,i}$ | Simulated event time for event $e$ in country/region $c$ in the $i$'th simulated dataset under sample size $d$ |
| $T_{c,e,d,i}^D$ | Detection time (difference between simulated and true event times) for event $e$ in country/region $c$ in the $i$'th simulated dataset under sample size $d$ |
| $Pr(T_{c,e,d}^D < \infty)$ | Power to detect event $e$ in country/region $c$ under sample size $d$, equal to the proportion of the $I$ simulated datasets where the event occurs |

#### C.1 Events 1 to 4: a metric crossing a threshold

Denote by *µ_c,t,d,i_*the metric of interest for country/region *c* in week *t* in the *i*th simulated dataset under sample size *d*, for example the proportion positive *π_c,t,d,i_* or the influenza activity proxy ILI+ *f_t,Flu,d,i_*, or similarly for the ARI-based activity proxies. Denote the equivalent “ground truth” metric as *µ_c,t_^TRUE^*.

Denote the threshold of interest by *τ* . Denote by *T_c,_*_1_*^TRUE^* = arg min*_t_* [*µ_c,_*_t_*^TRUE^*≥*τ*] the true time the threshold is first crossed (Event 1, denoted by the index 1) and the equivalent time in the *i*th simulated dataset by

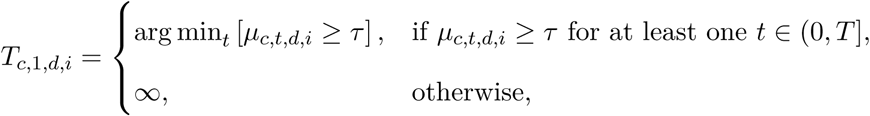

i.e. it is defined conditional on the event occurring in both the ground truth and the simulated dataset *i* and set to infinite otherwise.

Then the empirical distribution of the time to detection of the event *T*_c,1,*d,i*_*^D^* can be constructed from the set of time differences:

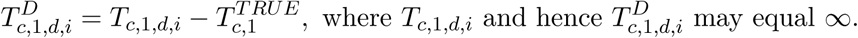

The distribution of the detection times conditional on the event occurring is therefore given by the subset of times *T*_c,O1,*d,i*_*^D^ <* ∞ and the probability a metric crosses the threshold at least once in the time period (0*, T* ] is the proportion of simulated datasets where the event occurs:

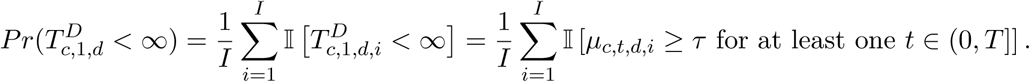

The detection time distribution can be summarised using the mean, median and 2.5 and 97.5 percentiles of the times *T*_c,1,*d,i*_*^D^ <* ∞.

Since the estimates of *µ_c,t,d,i_*are centred around the true metrics on average regardless of sample size, these probabilities are not expected to vary across sample sizes *d* (see e.g. Figure 2). Instead, we can consider slightly more complex events to detect. Event 2 is a metric being greater than a threshold for at least two consecutive weeks. The true and simulated times this event first occurs are given by:

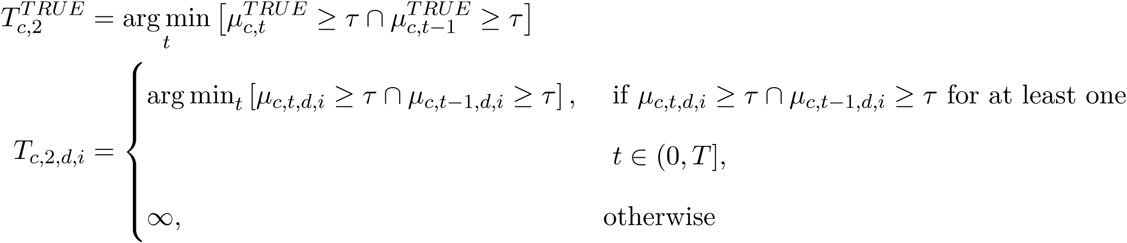

The conditional times to detection and the probability of detection are defined analogously to those for Event 1.

Events 3 and 4 are detected when a metric is statistically significantly above a threshold for the first time (Event 3) and for at least two consecutive weeks (Event 4), as determined by the lower bound of a 95% CI being above the threshold. The times when the true metric crossed the threshold first and for two consecutive weeks are defined 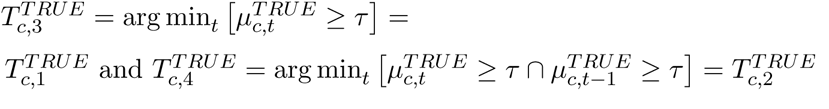.

Let the lower CI bound for the proportion positive be denoted as *λ_c,t,d,i_* for the *i*th simulated dataset. It is obtained using the Clopper-Pearson exact binomial confidence interval. The corresponding lower bound for the influenza activity proxy ILI+, in the case that ILI consultation rates are considered fixed, is just the positivity lower bound multiplied by the ILI consultation rate. The simulated times to the lower bound crossing a threshold, followed by the equivalent for being above the threshold for at least two weeks are defined as:

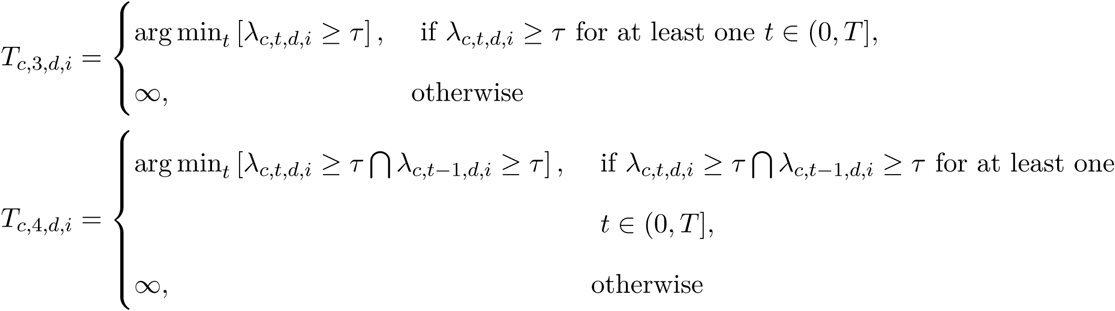

The conditional times to detection and the probability of detection are defined analogously to those for Event 1.

#### C.2 Events 5 & 6: three consecutive weeks of increase or decrease in a metric

These events are detected if 3 monotonic increases or decreases respectively in a metric *µ_c,t_*occur. The true and simulated times that such a monotonic change is observed are defined as

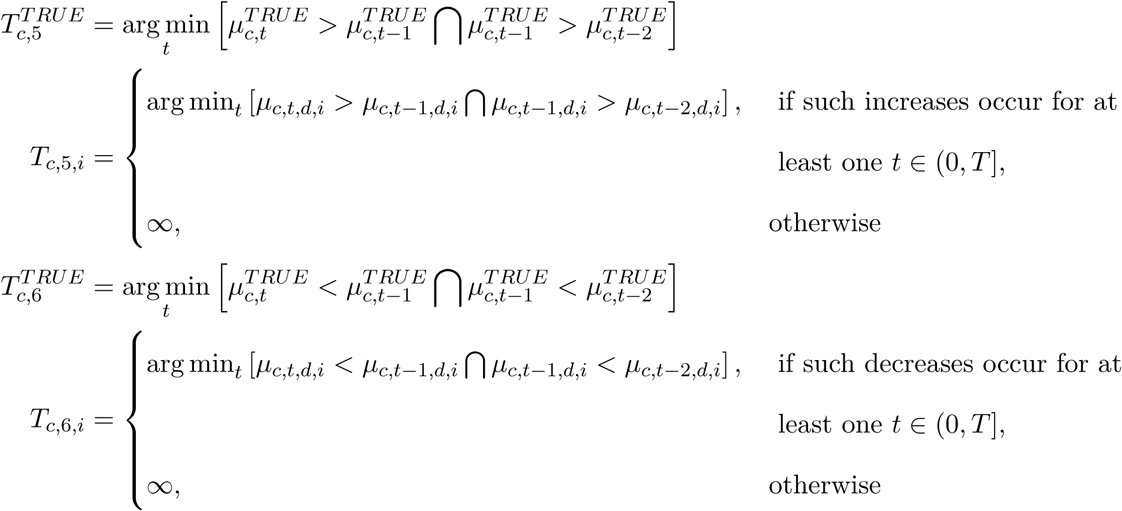

The conditional times to detection and the probability of detection are again defined analogously to those for previous events.

#### C.3 Events 7 and 8: growth or decline rate in a metric statistically significantly greater or less than a threshold

For a true metric *µ_c,t_^TRUE^* for country/region *c* in week *t*, denote the corresponding numerator and denominator for the proportion or rate by *y_c,t_^TRUE^* and *n_c,t_^TRUE^*. Similarly, for each of the *I* simulated datasets, denote as above the corresponding numerator and denominator as *y_c,t,d,i_* and *n_c,t,d_* where *d* indexes the sample sizes and *i* = 1*, . . . I* indexes the simulated datasets.

For each week in the season apart from the first week, *t* ∈ (1*, T* ], we fit a separate log-linear Poisson regression of the ground truth metric’s numerator on time, offset by its denominator, using the cumulative data up to week *t*:

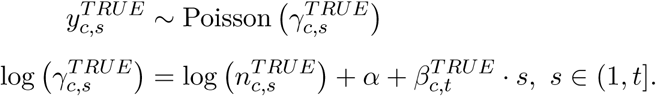

Based on the model, the growth rate up to week *t* is *ρ_c,t_^TRUE^* = exp(*β_c,t_^TRUE^*) and denote the corresponding exponentiated 95% CI by (*λ_c,t_^TRUE^*, *υ_c,t_^TRUE^*). The true time at which a growth rate is first significantly greater than a threshold *τ* in the time period under consideration, (0*, T* ], is defined as the first week *t* when the estimated lower bound of the CI *λ^_c,t_^TRUE^* using the ground truth data up to week *t* is larger than *τ* :

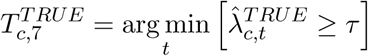

Denote by 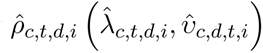 the equivalent estimated growth rate (95% CI) based on a log-linear model of each simulated metric’s numerator *y_c,t,d,i_* on the weeks of time in (0*, t*] offset by the denominator *n_c,t,d_*. Denote by 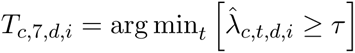 the equivalent earliest time at which a growth rate significantly greater than *τ* is estimated in the *i*th simulated dataset, conditional on that observation occurring. If the event does not occur in simulated dataset *i*, or if it does not occur in the ground truth data, then *T_c,_*_7_*_,d,i_* is set to be infinite.

Then the empirical distribution of the time to detection of the event is given by the set of time differences:

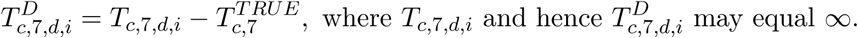

The distribution of the detection times conditional on the event occurring and the probability of detection are then defined analogously to previous events.

Sample sizes to detect Event 8 are assessed equivalently, but by comparing the upper bound of the 95% CI for the decline rate estimate *υ*^*_c,t,d,i_* to the threshold *τ* , rather than the point estimate, for both the ground truth and each simulated dataset.

Note that when the metric of interest is an activity proxy, and when assuming that the corresponding ILI or ARI consultation rate is fixed, so that all variability comes from the proportion positive, we consider the number of positive tests for the pathogen of interest to be the numerator *y_c,t,d,i_*; and the offset *n_c,t,d_*to be equal to the number of swabs tested divided by the ILI or ARI consultation rate. This consideration ensures the growth or decline rate estimated from the log-linear model is indeed for the ILI+ or ARI-based activity rate, but also allows a correct reflection of the variability in the proportion positive data.

#### C.4 Events 9-12: growth or decline rate in a metric over the most recent 4 weeks (statistically) greater or less than a threshold

These events are similar to Events 7-8 in that the growth/decline rate of a metric is estimated as the slope of a log-linear regression of the metric numerator on week *t*, offset by the denominator, but based only on the most recent four weeks of data, rather than the cumulative data up to week *t*. Note that since there is extra complexity in estimating growth/decline rates from short time series of data, we consider both detecting a growth/decline rate estimate itself being greater than/less than a threshold (Events 9 and 11), as well as detecting statistically significant departures from a threshold (Events 10 and 12).

For each week *t* ∈ (4*, T* ], the following log-linear regressions are fitted for the true and simulated datasets:

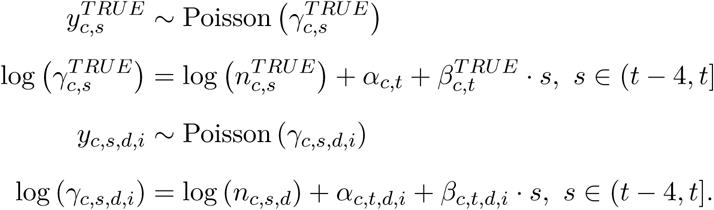

Denote by 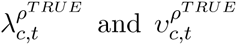 the lower and upper bounds of the 95% confidence interval estimated for 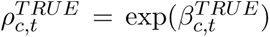 Similarly, let 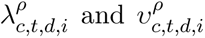 be the 95% confidence interval estimated for *ρ_c,t,d,i_* = exp(*β_c,t,d,i_*).

Then for a threshold *τ* , the true and simulated event times (i.e. times the growth/decline rate are greater or less than *τ* for Events 9 and 11; with statistical significance for Events 10 and 12) are defined as:

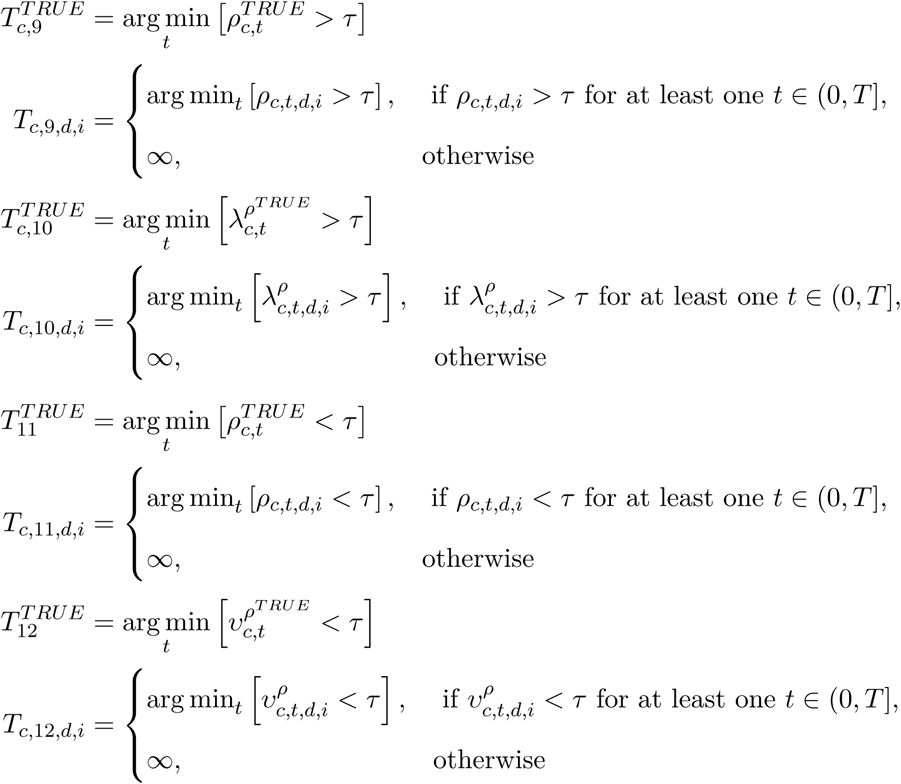

The conditional times to detection and the probability of detection are again defined analogously to those for previous events. When considering an activity rate as the metric of interest, the numerator and offset used are defined the same as for Events 7-8.

#### C.5 Integrated surveillance: Events 5-6 trend detection

Denote by *µ_c,t,p,d,i_*the metric of interest for for pathogen *p*, where *p* takes the values

{*Flu, SC*2*, RSV, Pos*} corresponding with influenza, SARS-CoV-2, RSV and all other tested respiratory pathogens. Denote the corresponding “ground truth” metric as *µ_c,t,p_^TRUE^*. Analogous with the single-pathogen simulations for Events 5 and 6, we consider detection of a monotonous increase or decrease over three consecutive weeks of *any* of the pathogens considered for integrated surveillance. The true and simulated times that such a monotonic change is first observed for at least one pathogen are defined as

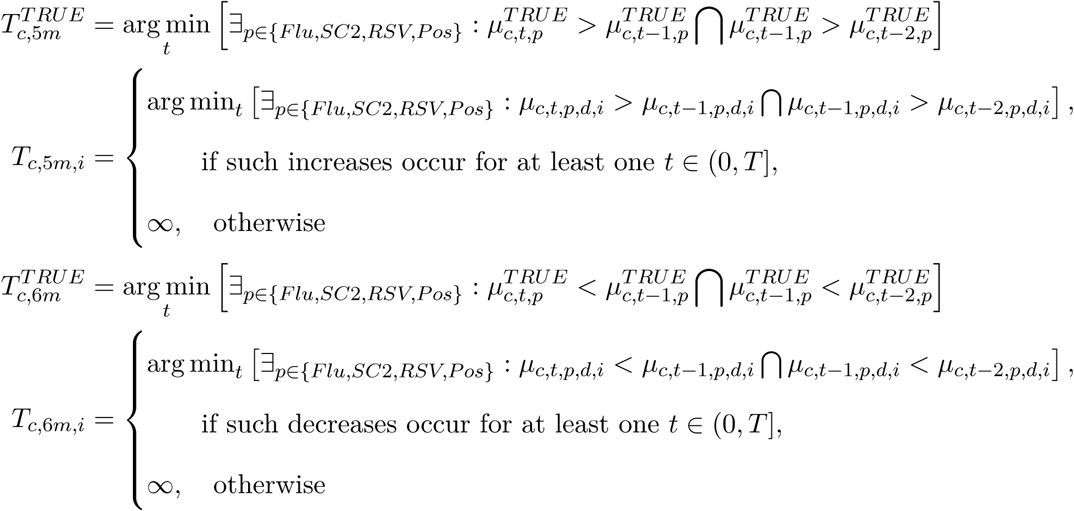

The conditional detection times and probabilities are again defined analogously to those for previous events.

#### C.6 Integrated surveillance: Events 7-12 growth/decline rate detection

These events and detection summaries are defined analogously to the single pathogen ones, but considering detection of the event (growth/decline rates greater/less than a threshold, potentially with significance) in at least one of the tested pathogens. However, since growth or decline rates in a pathogen may be correlated with the growth/decline rates in other co-circulating respiratory pathogens, confidence intervals for the growth/decline rates need to take account of such correlation.

The data on positive and negative test results given the number of swabs tested can be considered realisations of a multinomial distribution:

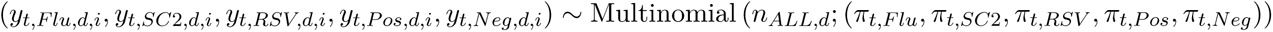

where *Pos* denotes positive for any other respiratory pathogen tested and *Neg* denotes negative for all pathogens tested.

However, rather than considering a multinomial regression of the positive tests on time, we can equivalently consider a series of conditional Poisson regressions for each pathogen, offset by a denominator that accounts for not having tested positive for a previous pathogen. In particular, we regress the influenza positive tests on time, offset by the total number of tests; then the SARS-CoV-2 positive tests on time, offset by the number tested minus the number that tested positive for influenza, i.e. conditional on being influenza-negative; third, we regress the RSV positive tests on time, conditional on not having tested positive for either influenza or SARS-CoV-2; and finally, the remaining positive tests on time, conditional on not having tested positive for influenza, SARS-CoV-2 or RSV. Mathematically, for each simulated dataset *i* ∈ 1*, . . . I* under each sample size indexed by *d*, we fit the following Poisson log-linear regressions:

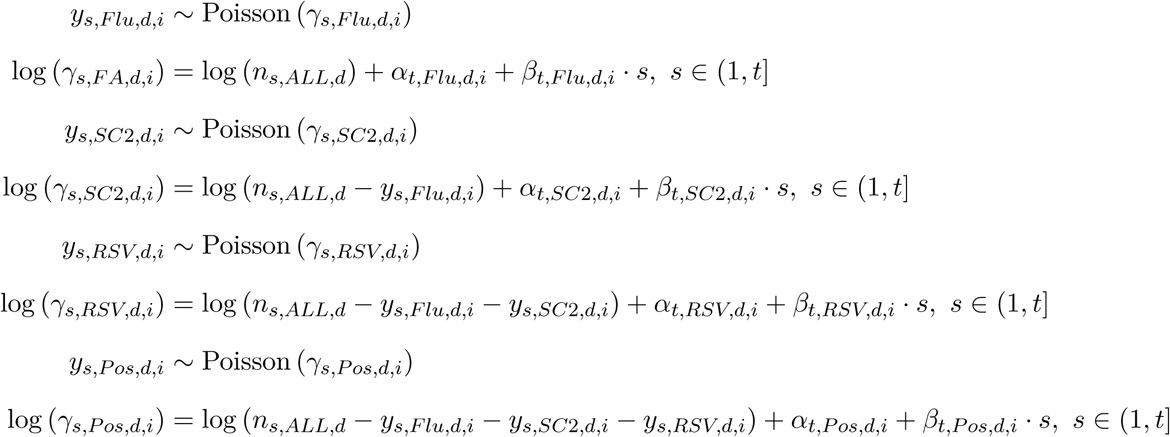

to obtain growth/decline rate estimates 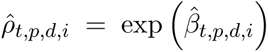 and corresponding 95% Cis 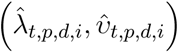 for each pathogen *p*.

The same conditional regressions are fit to the ground truth data, and the detection times and probabilities of detection are obtained as for previous events.

### D Results

#### D.1 Simulated datasets

Figure D.1 shows the median and 2.5 and 97.5 percentiles over the simulated ILI+ activity and proportion positive for influenza metrics, under different assumed swabbing sample sizes. These plots demonstrate how the proportion positive and therefore the ILI+ metric are simulated with increasing precision with increasing sample size.

#### D.2 Main analyses

##### D.2.1 ILI+ activity rate

Conditional on a growth or decline rate in ILI+ activity statistically significantly greater or less than a threshold occurring in both the ground truth data and at least one simulated dataset, detecting Events 7 and 8 is straightforward in terms of 100% detection rates, even at small weekly sample sizes (Figure D.4). However, the detection times, particularly for the USA and Hong Kong, decrease with sample size, e.g. for Event 7, from 8 (6-10) weeks at 25 swabs tested a week to 5 (0,6) weeks at 500 swabs a week in Hong Kong for growth rates greater than 1.1 (10%).

##### D.2.2 ARI-based activity rates

For detecting at least three weeks of increase or decrease in the ARI-based activity metrics (Events 5 and 6), Figure D.7 shows that 50 swabs a week is sufficient for ARI+ and ARI-flu, in terms of detection probabilities greater than 75%. However, detection times and particularly their uncertainty reduce with increasing sample size, with ARI+ requiring at least 1000 swabs a week to ensure the confidence interval for the detection time is no more than 3 weeks wide. For ARI-SC2, at least 500 swabs a week is needed to reach a 75% detection probability for Event 5, 3 weeks increase, whereas 250 a week is sufficient for detecting Event 6, 3 weeks decrease with more than 75% probability. It is notable that the uncertainty in the detection times for all three activity proxies is somewhat greater for Event 6 than for Event 5.

##### D.2.3 ARI-flu vs ILI+ activity rates

Figure D.14 shows that a decline more extreme than 0.9 (-10%) is not observed in either the ground truth or any of the simulated datasets for either metric, and a decline more extreme than 0.95 (-5%) is not observed in the ground truth, but is in some of the simulated datasets. While for the ILI+ metric the detection probabilities for threshold 0.95 are all at 100%, whatever the sample size, they increase with sample size for the ARI-flu activity metric, with 500 swabs per week needed to achieve more than a 75% detection probability. Detecting any decline (threshold 1) is achievable at 100% probability for any sample size for both metrics, with the corresponding detection times decreasing and becoming more certain with sample size. It is notable that the uncertainty in the detection times for ARI-flu is more pronounced than the uncertainty for ILI+.

##### D.2.4 Multiple pathogen proportion positive

When considering the joint proportions positive for multiple pathogens in an integrated analysis, the summaries for detecting Events 5 and 6 (at least three consecutive weeks of increase or decrease) for at least one pathogen among all those tested are shown in Figure D.16. The minimum sample size of 25 swabs a week is sufficient for high (*>* 75%) detection probabilities, but detection times improve with larger sample sizes. For this 2022/23 season, it is notable that the pathogen for which an increase is most often detected first is influenza, followed by the “Pos” category (positive for any other respiratory pathogen tested), whereas decreases are more often detected first for SARS-CoV-2 at higher sample sizes.

Figure D.17 shows the detection summaries for Events 7 and 8, growth or decline rates significantly greater or less than a threshold. Here it is noticeable that the detection probabilities increase with the number of swabs tested, e.g. for threshold 1.1 for the growth rates and threshold 0.9 for the decline rates. The detection times for significant growth also decrease with sample size, but from around 1 week to negative values, reflecting the variability when simulating proportions positive that results in events occurring earlier in the simulated datasets than in the ground truth data. For detecting growth rates significantly above a threshold, influenza is the pathogen always detected first (the NAs in the plot reflect the missing values for detection summaries when either the event does not occur in the ground truth data or does not occur in any of the simulated datasets). Whereas for detecting decline rates significantly lower than a threshold, the pathogen most often detected first varies with threshold and sample size: influenza for threshold 1 at smaller sample sizes; RSV for threshold 0.95 at every sample size; and influenza for threshold 0.9, although many simulations do not include this event at all for this threshold.

#### D.3 Supplementary analyses

##### D.3.1 Proportion positive for influenza, ILI swabs as denominator

##### **D.3.2** Alternative denominator (all swabs) for proportion positive for in-fluenza, England

Using the alternative denominator of *all* swabs tested, *y_t,ALL_* (rather than ILI swabs tested *y_t,ILI_* ) for the proportion of tests positive for influenza results in a generally lower proportion positive, more comparable with the proportions observed in the USA and Hong Kong (bottom panel of Figure A.3 compared to middle row of Figure 1.

The resulting assessments of the probabilities of and times to detection (Figures D.25 to D.30) therefore show that detecting the proportion positive crossing a common threshold of 0.25 (lower threshold for the ILI swab denominator, upper threshold for the ARI/all swabs denominator) is more challenging using the larger all swabs denominator than the smaller ILI denominator.

---

**Figure D.1:**
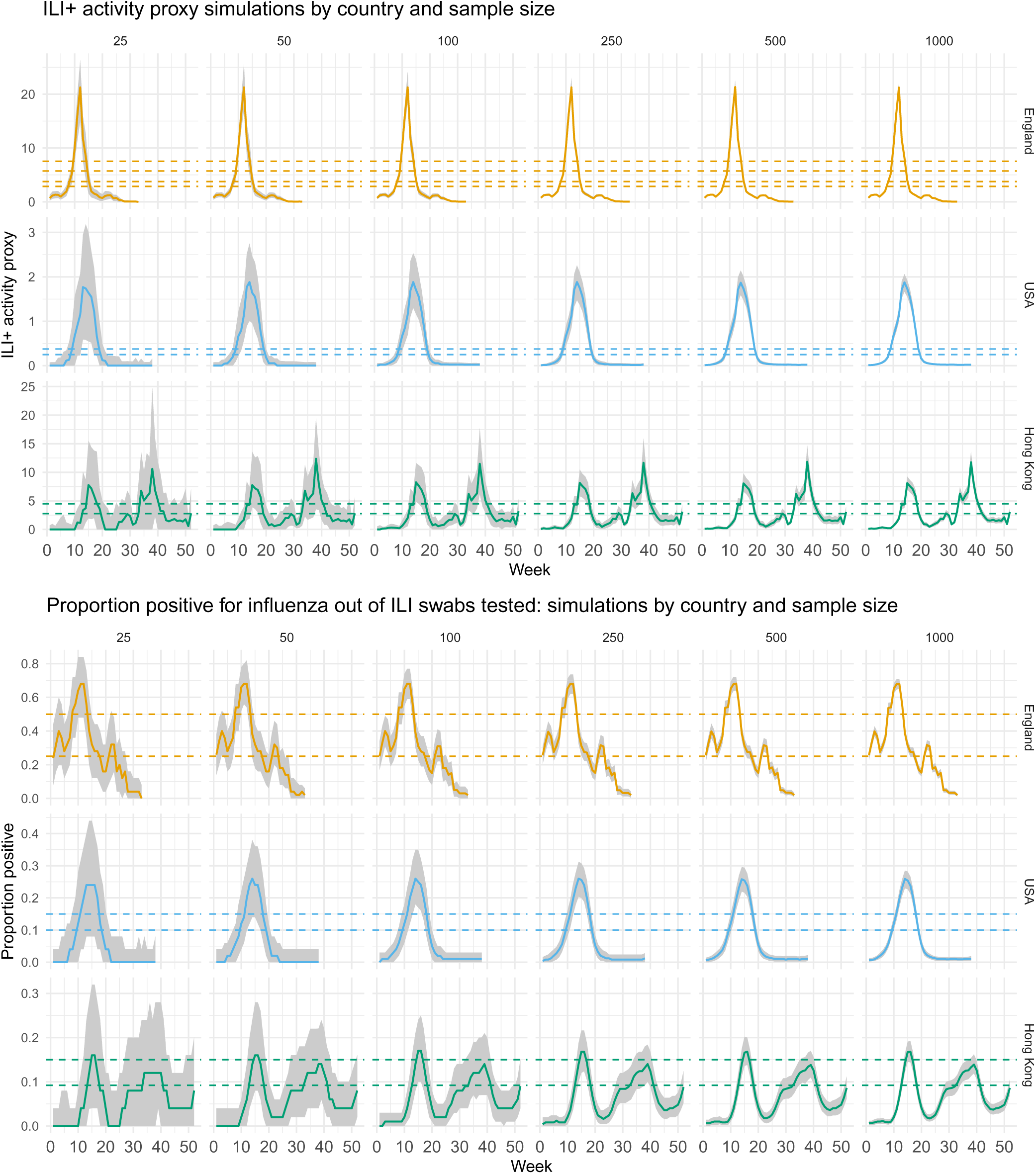
Median (black) and 95% CI (quantile-based, grey) influenza activity proxy (top) and proportion positive for influenza (bottom) across all simulated datasets, by country and number of samples swabbed/tested per week (positivity denominator). The horizontal dashed lines show the different values used for detecting crossing a threshold (Table B.1).

**Figure D.2:**
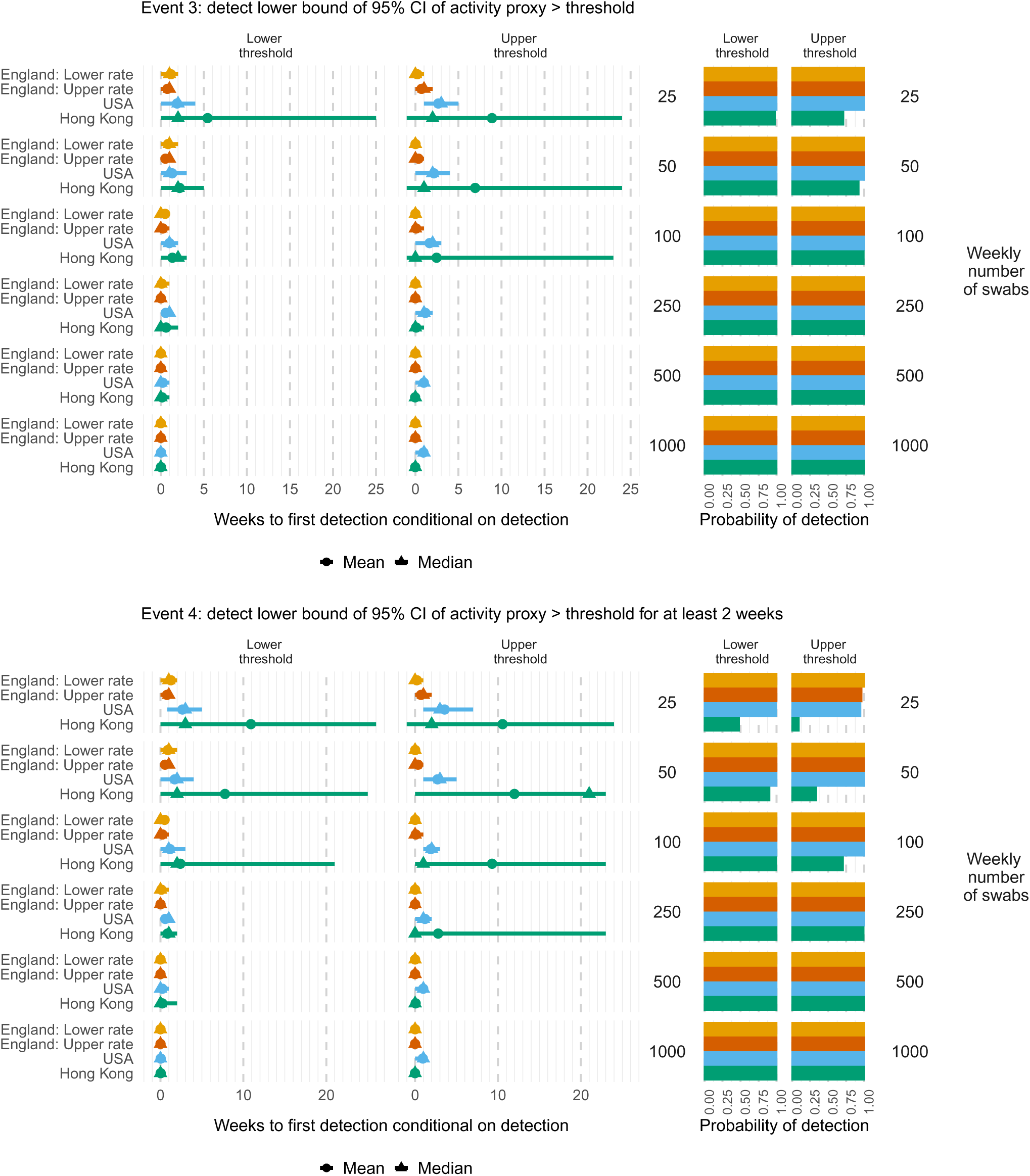
Times to (left) and probabilities of (right) detecting ILI+ activity significantly greater than a country/region-specific lower or higher threshold (see Table B.1) by Event (3 top; 4 bottom), weekly number of swabs tested, threshold and country/region. Mean (circles), median (triangles) and 2.5 and 97.5 percentiles (line ranges) of times to detection are conditional on detection occurring. The probabilities of detection are obtained as the proportion of simulated datasets where the event occurs.

**Figure D.3:**
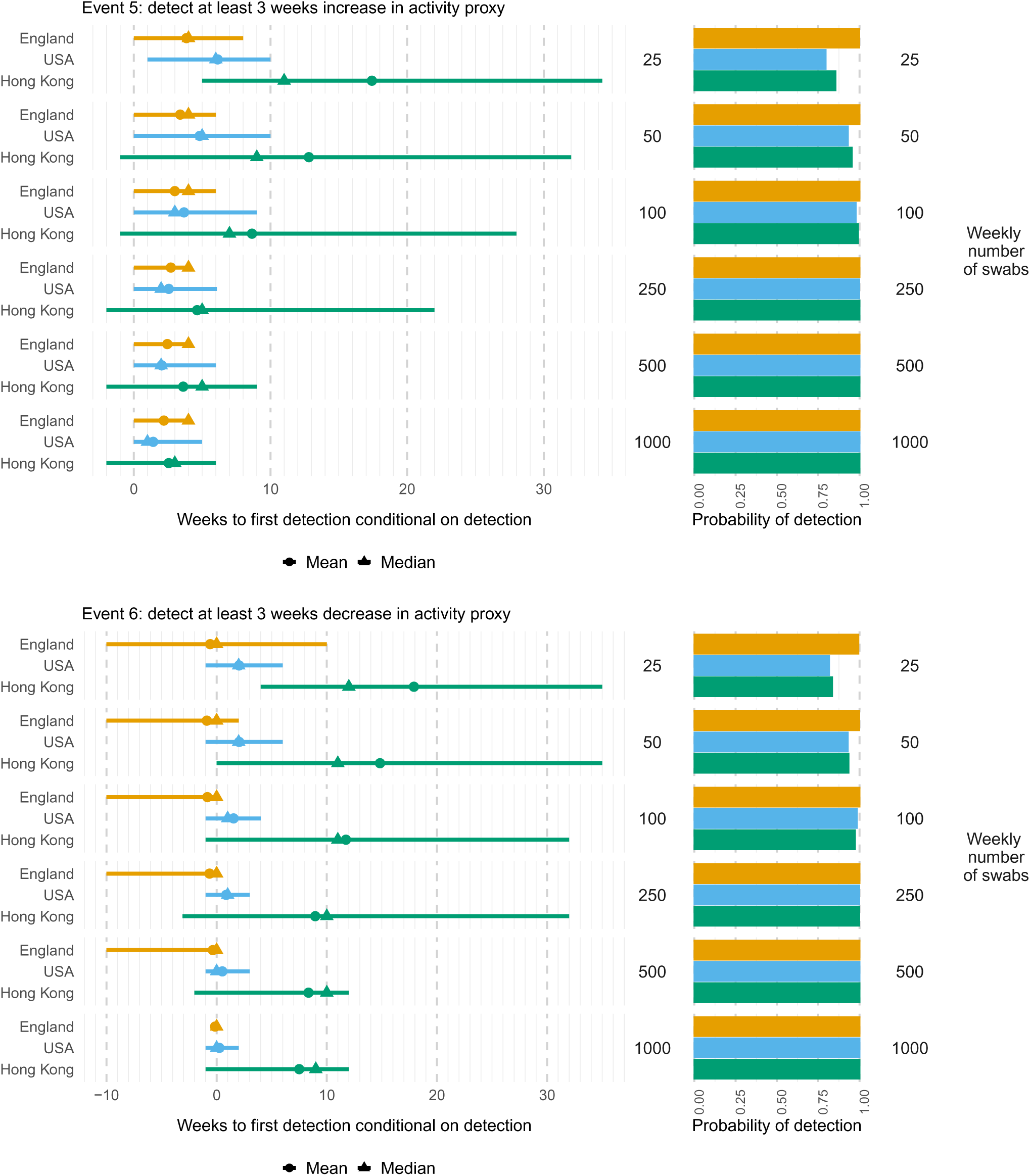
Times to (left) and probabilities of (right) detecting at least 3 consecutive weeks increase (top) or decrease (bottom) in ILI+ activity, by weekly number of swabs tested and country/region. Mean (circles), median (triangles) and 2.5 and 97.5 percentiles (line ranges) of times to detection are conditional on detection occurring. The probabilities of detection are obtained as the proportion of simulated datasets where the event occurs.

**Figure D.4:**
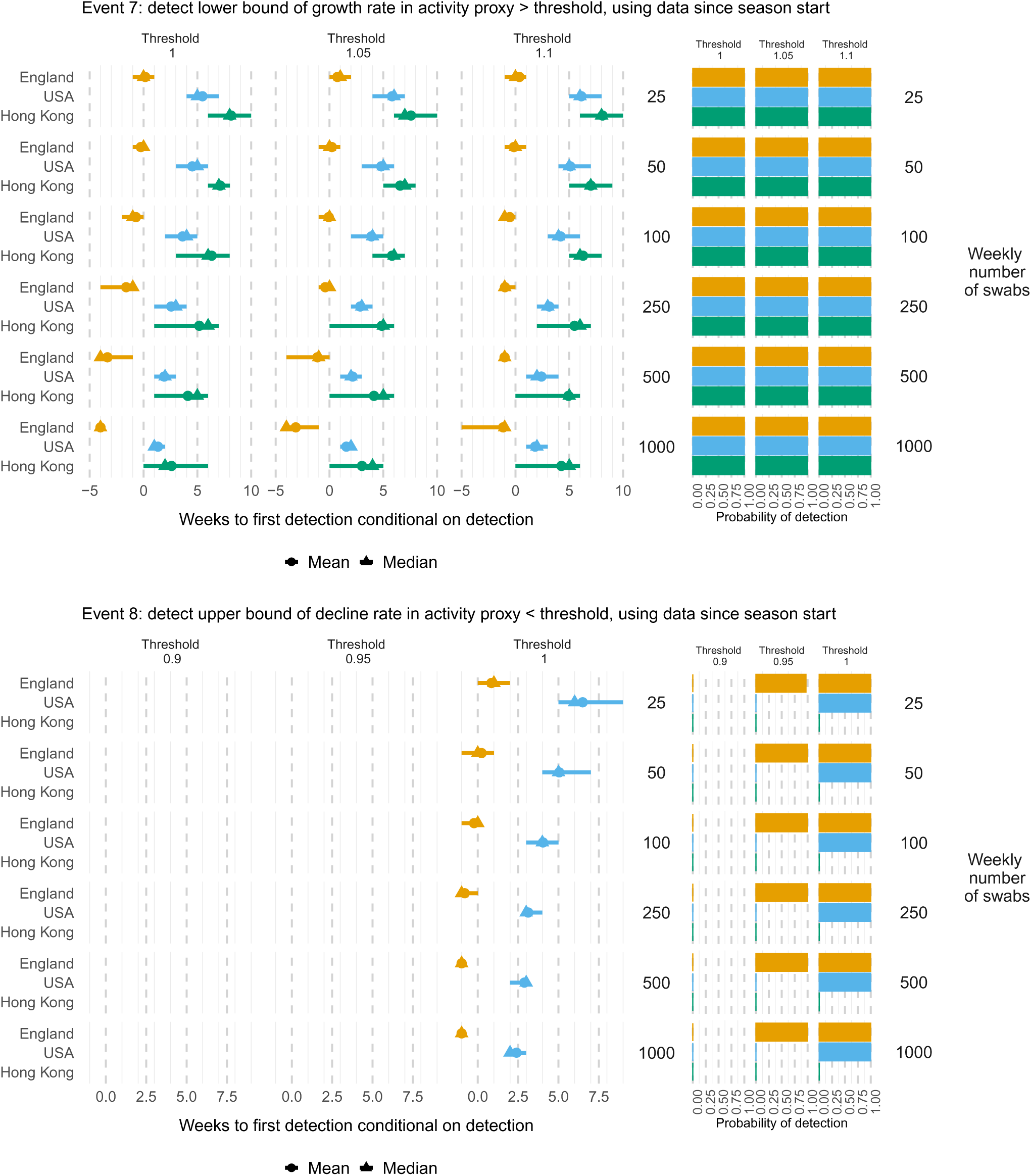
Times to (left) and probabilities of (right) detecting at a growth rate (top) or decline rate (bottom) in ILI+ activity significantly greater or smaller respectively than specified thresholds, based on all data since the start of the season, by weekly number of swabs tested and country/region. Mean (circles), median (triangles) and 2.5 and 97.5 percentiles (line ranges) of times to detection are conditional on detection occurring. The probabilities of detection are obtained as the proportion of simulated datasets where the event occurs. Note that when either the event does not occur in the ground truth data or in any of the simulated datasets, then a distribution of detection times is not available (e.g. for threshold 0.9 for Event 8).

**Figure D.5:**
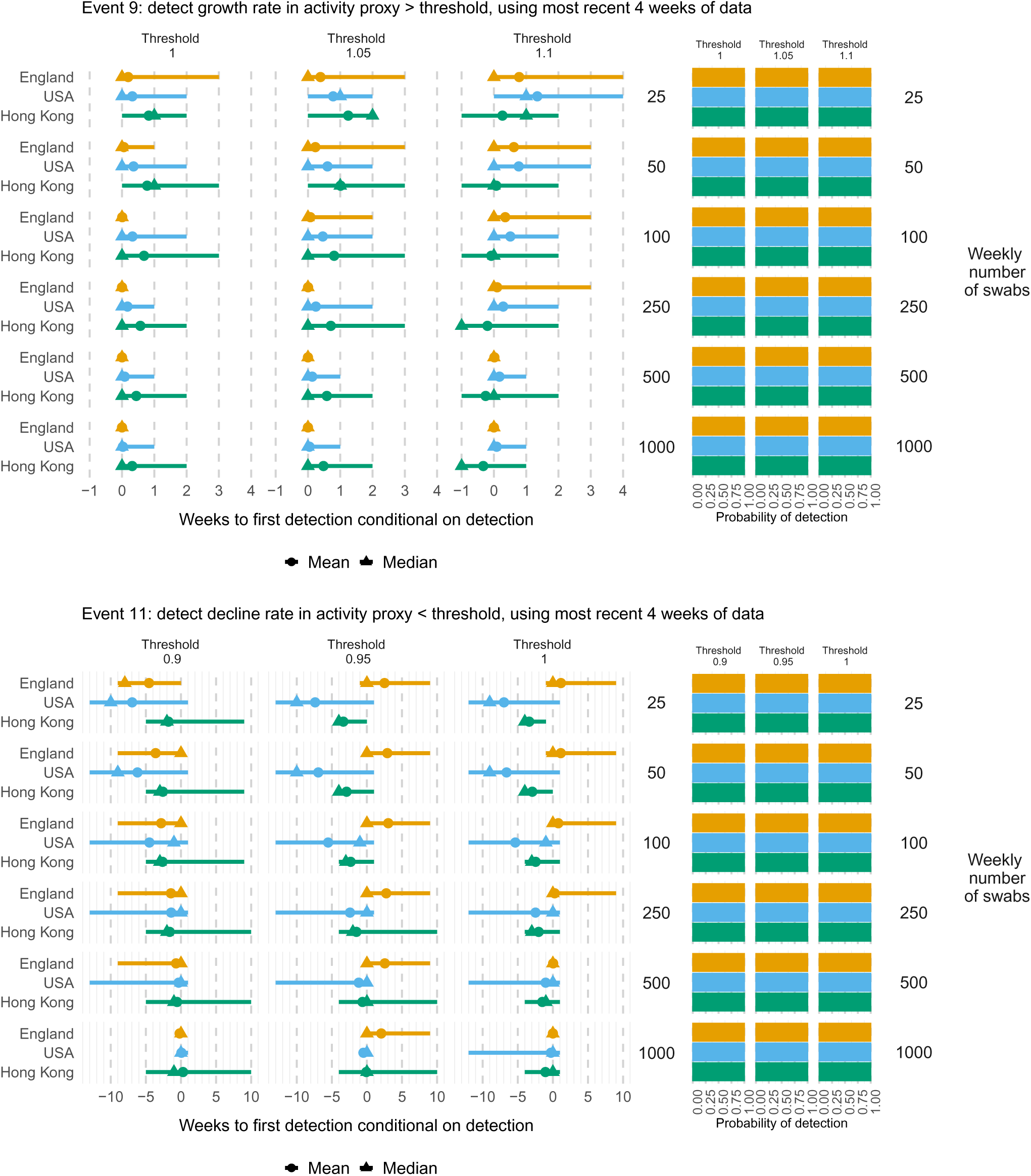
Times to (left) and probabilities of (right) detecting at a growth rate (top) or decline rate (bottom) in ILI+ activity greater or smaller respectively than specified thresholds, based on the last four weeks of data, by weekly number of swabs tested and country/region. Mean (circles), median (triangles) and 2.5 and 97.5 percentiles (line ranges) of times to detection are conditional on detection occurring. The probabilities of detection are obtained as the proportion of simulated datasets where the event occurs.

**Figure D.6:**
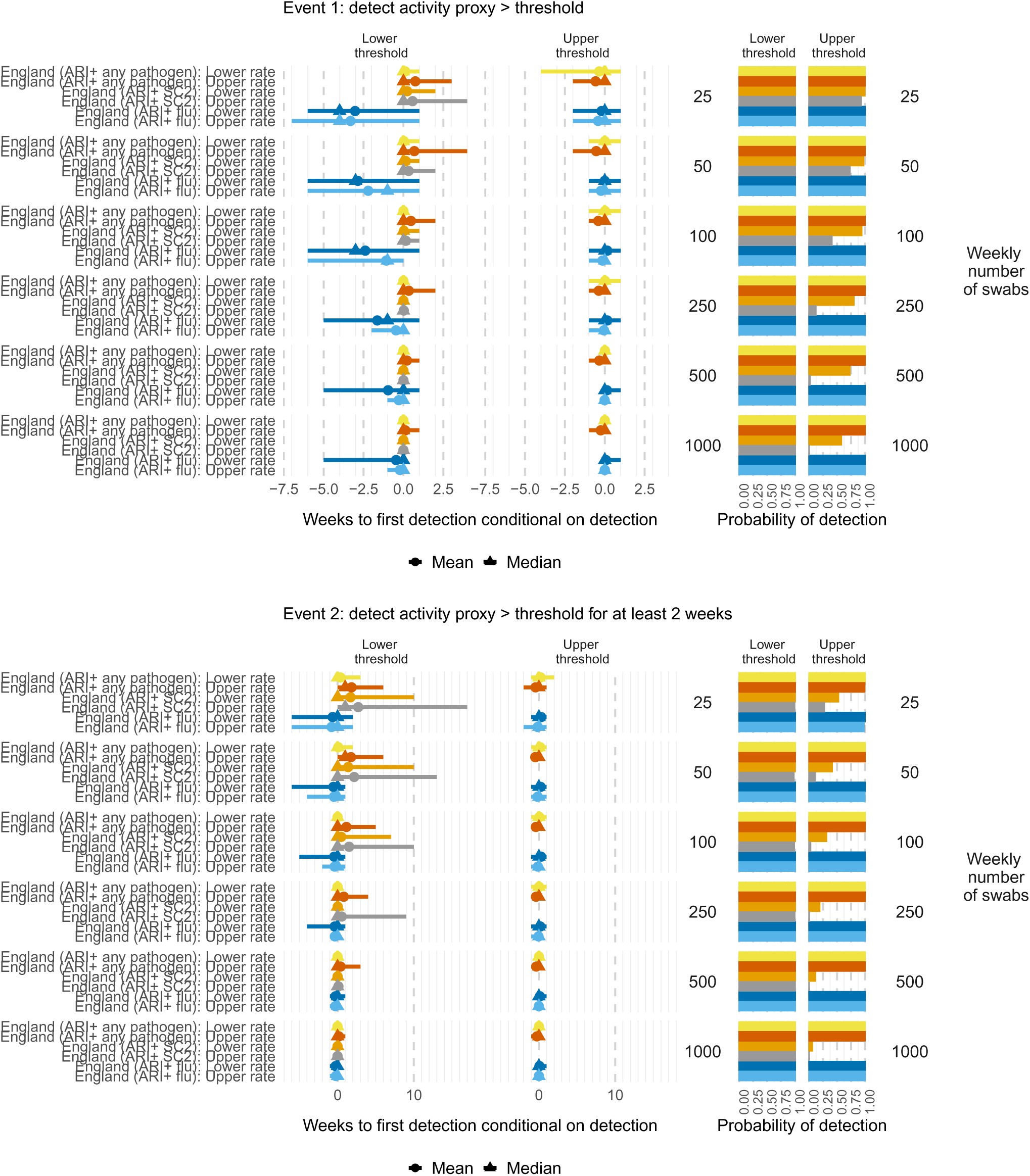
Times to (left) and probabilities of (right) detecting ARI-based activity metrics greater than specified thresholds (Event 1: top), for at least two weeks (Event 2: bottom), metric, weekly number of swabs tested and threshold (Table B.1). Mean (circles), median (triangles) and 2.5 and 97.5 percentiles (line ranges) of times to detection are conditional on detection occurring. The probabilities of detection are obtained as the proportion of simulated datasets where the event occurs.

**Figure D.7:**
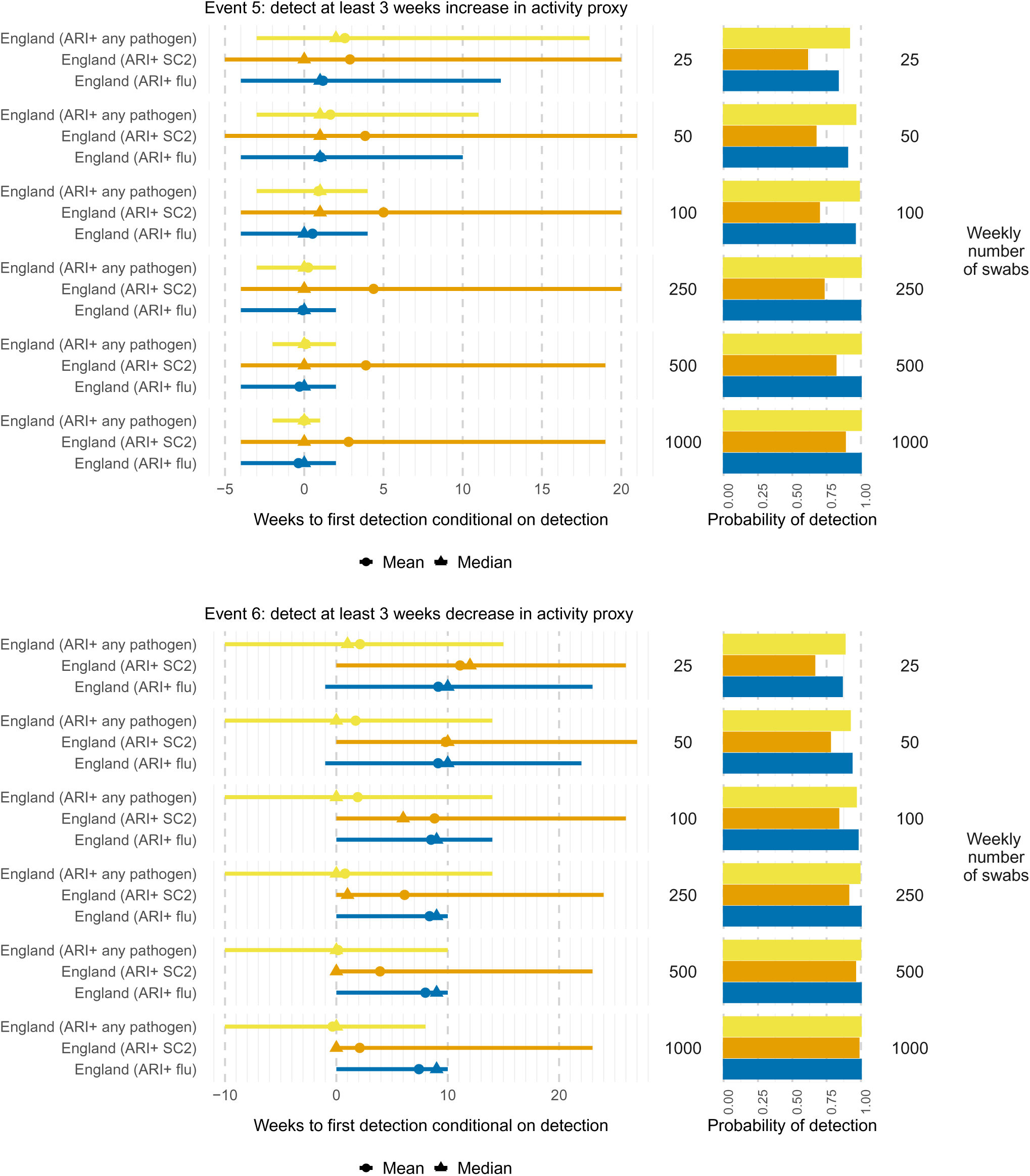
Times to (left) and probabilities of (right) detecting at least 3 consecutive weeks increase (top) or decrease (bottom) in ARI-based activity metrics, by weekly number of swabs tested and metric. Mean (circles), median (triangles) and 2.5 and 97.5 percentiles (line ranges) of times to detection are conditional on detection occurring. The probabilities of detection are obtained as the proportion of simulated datasets where the event occurs.

**Figure D.8:**
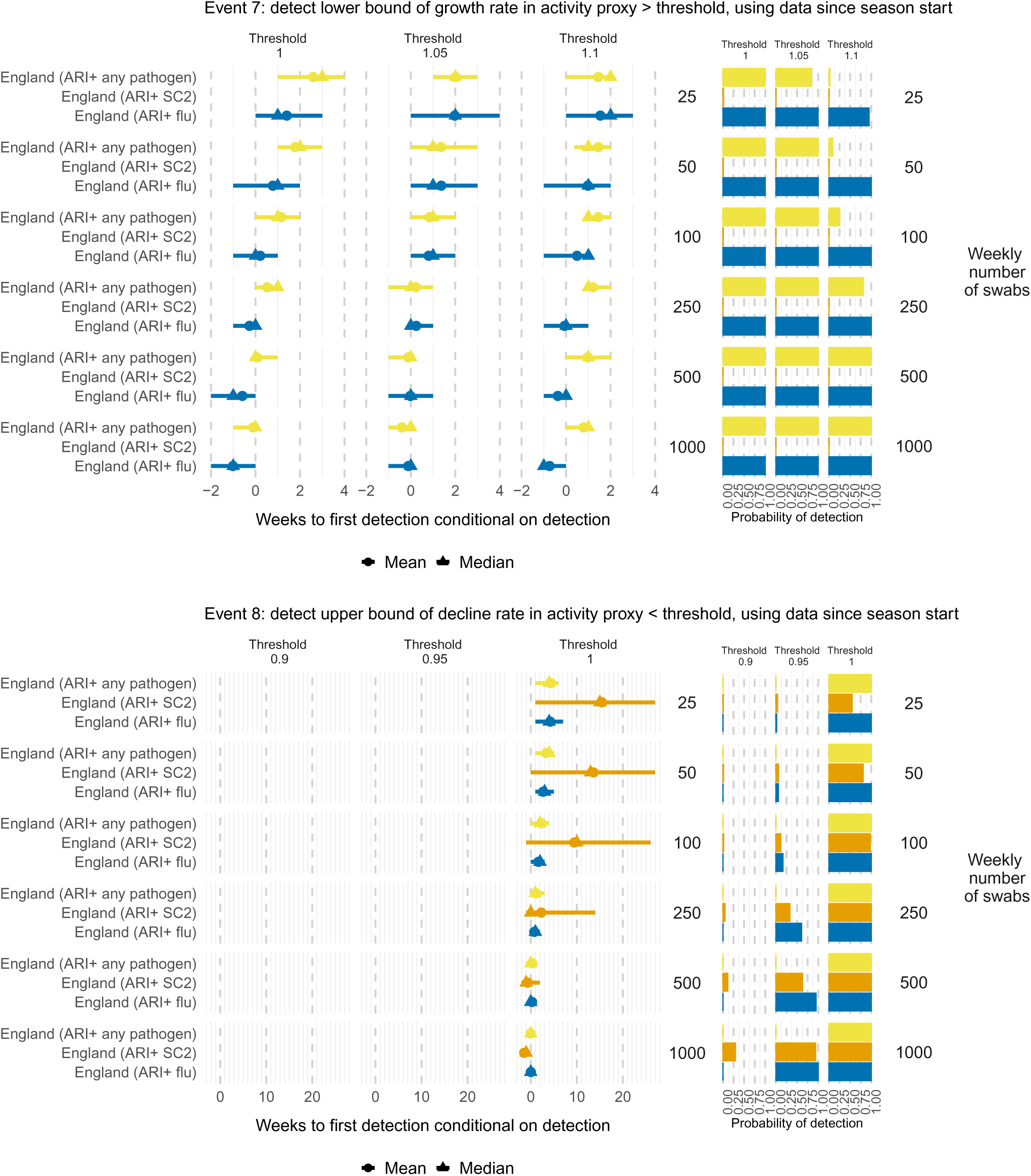
Times to (left) and probabilities of (right) detecting at a growth rate in ARI-based activity metrics significantly greater or lower than specified thresholds, based on all data since the start of the season, by weekly number of swabs tested and metric. Mean (circles), median (triangles) and 2.5 and 97.5 percentiles (line ranges) of times to detection are conditional on detection occurring. The probabilities of detection are obtained as the proportion of simulated datasets where the event occurs.

**Figure D.9:**
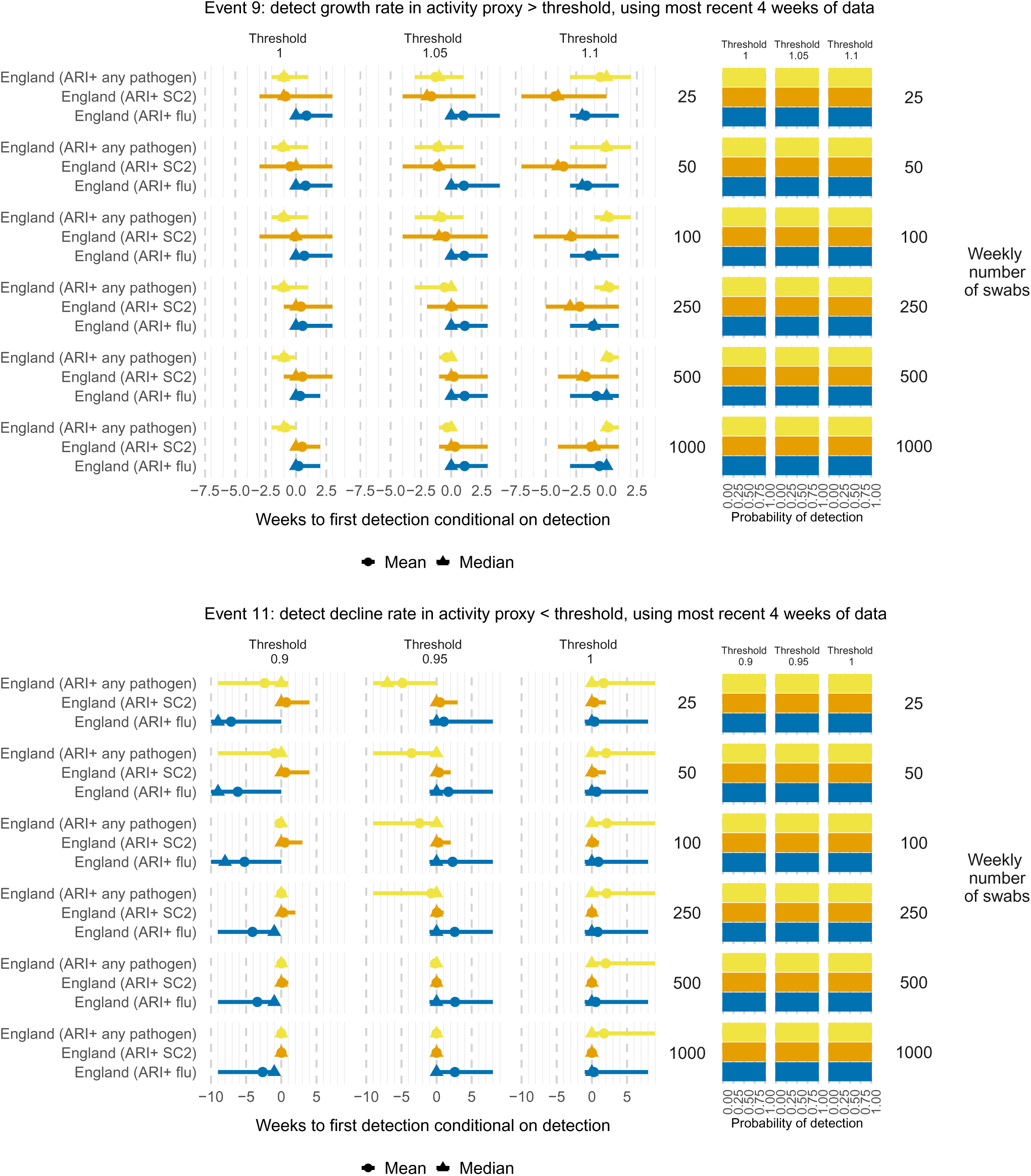
Times to (left) and probabilities of (right) detecting at a growth (top) or decline (bottom) rate in ARI-based activity metrics greater (top) or lower (bottom) than specified thresholds, based on the last four weeks of data, by weekly number of swabs tested and country/region. Mean (circles), median (triangles) and 2.5 and 97.5 percentiles (line ranges) of times to detection are conditional on detection occurring. The probabilities of detection are obtained as the proportion of simulated datasets where the event occurs.

**Figure D.10:**
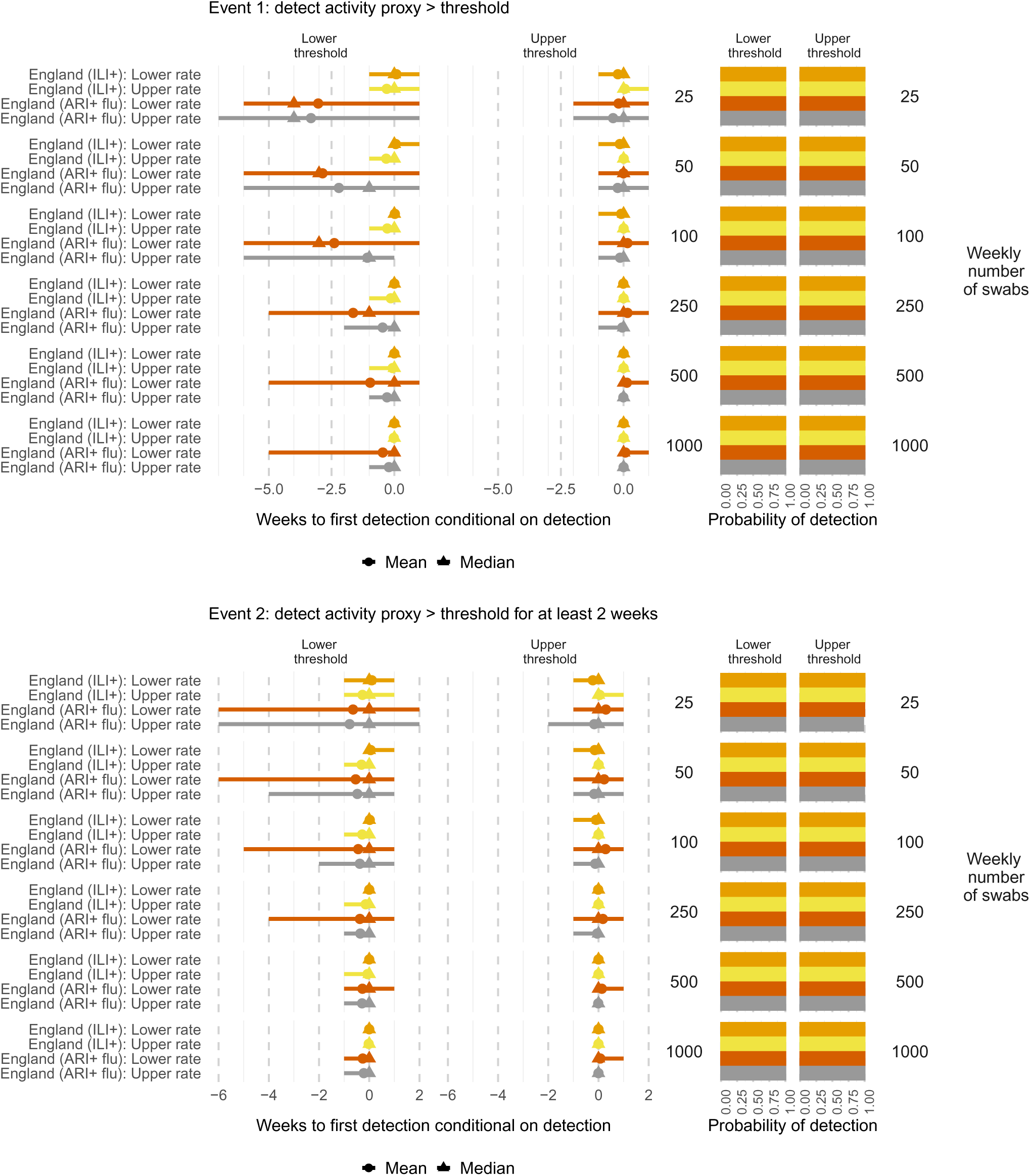
Times to (left) and probabilities of (right) detecting influenza activity greater than a threshold (Event 1, top), for at least two weeks (Event 2, bottom), by symptom profile for the consultation rate (ILI or ARI), weekly number of swabs tested and threshold (Table B.1). Mean (circles), median (triangles) and 2.5 and 97.5 percentiles (line ranges) of times to detection are conditional on detection occurring. The probabilities of detection are obtained as the proportion of simulated datasets where the event occurs.

**Figure D.11:**
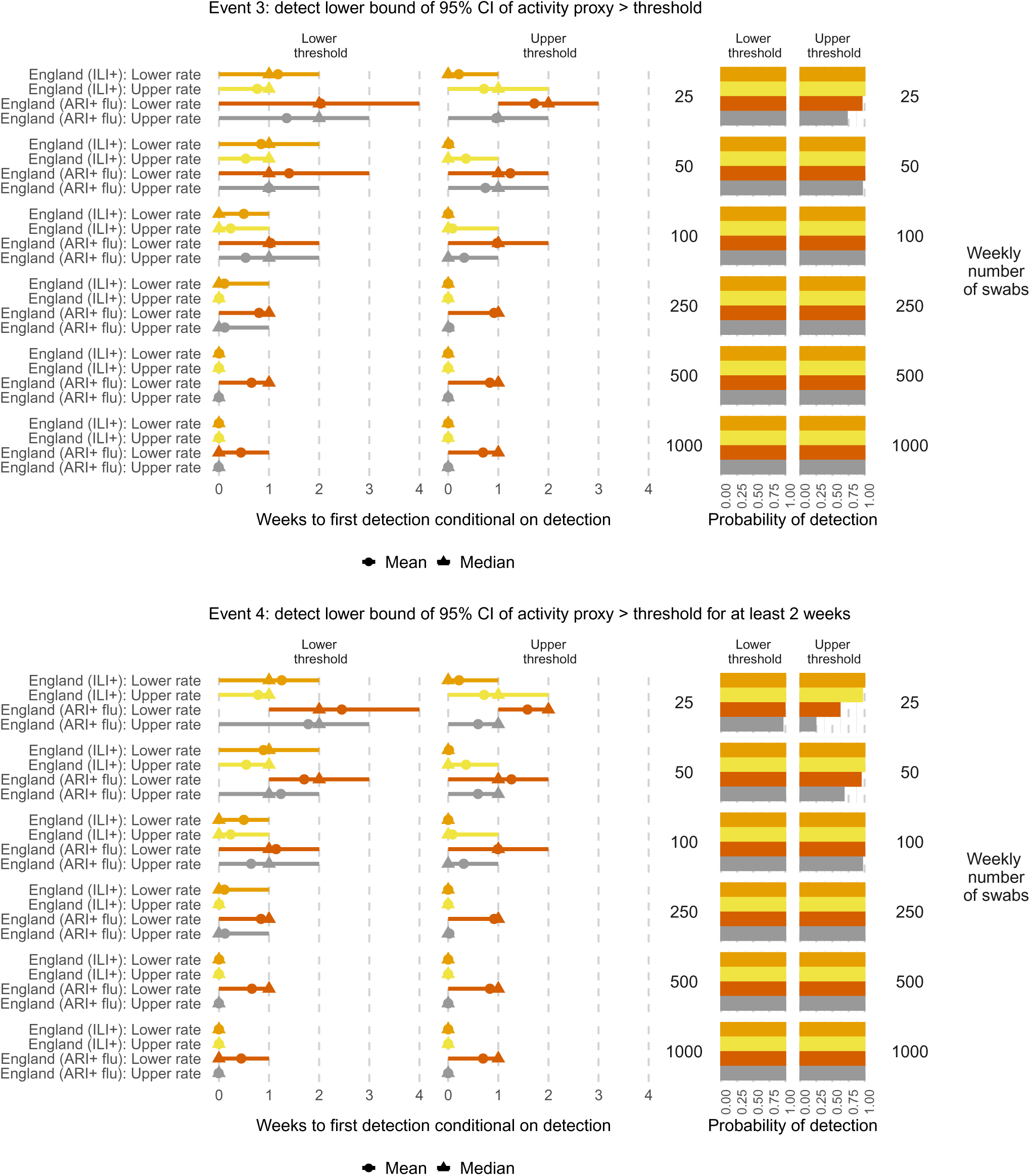
Times to (left) and probabilities of (right) detecting influenza activity significantly greater than a threshold (Event 3, top), for at least two weeks (Event 4, bottom), by symptom profile for the consultation rate (ILI or ARI), weekly number of swabs tested and threshold (Table B.1). Mean (circles), median (triangles) and 2.5 and 97.5 percentiles (line ranges) of times to detection are conditional on detection occurring. The probabilities of detection are obtained as the proportion of simulated datasets where the event occurs.

**Figure D.12:**
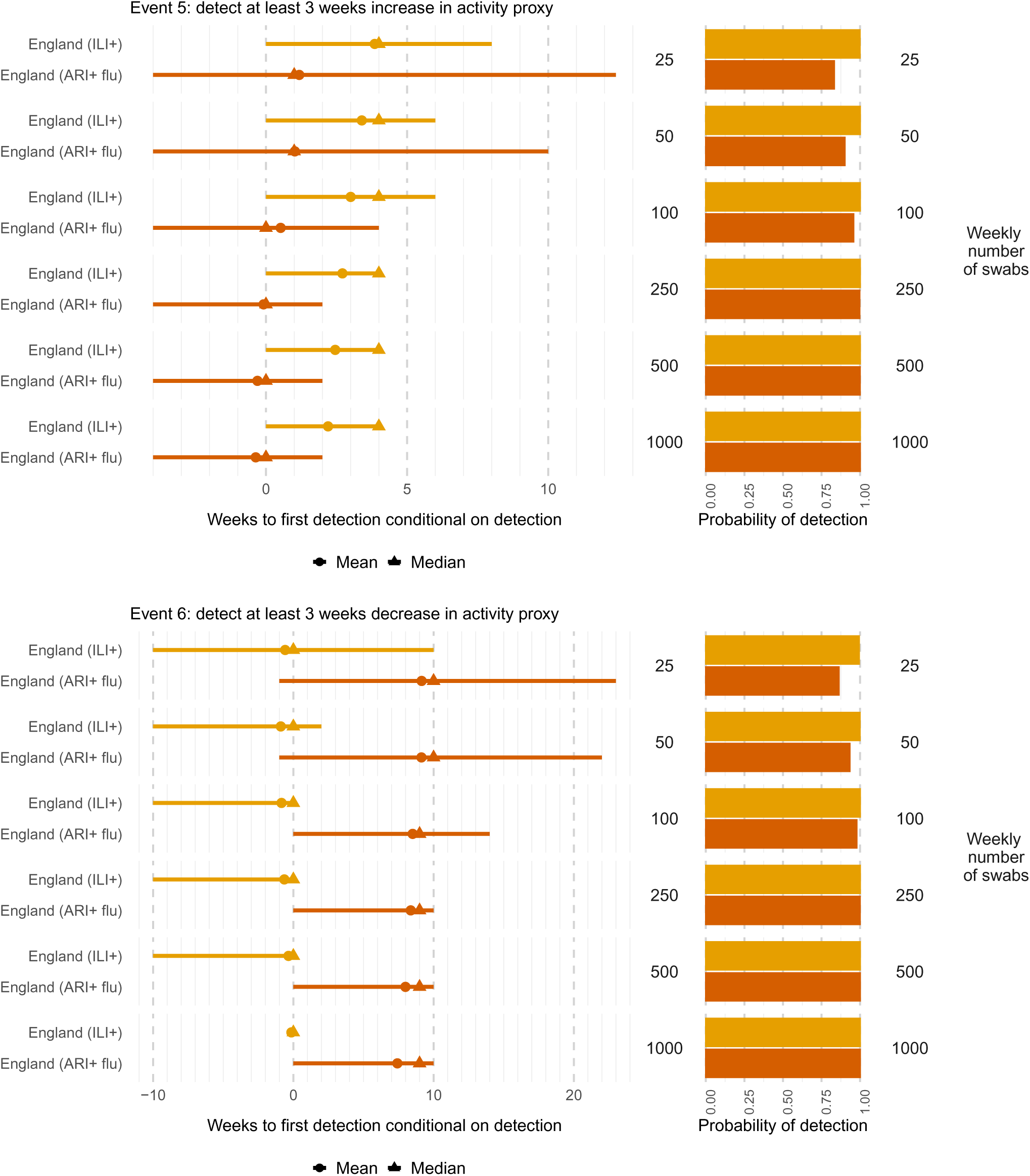
Times to (left) and probabilities of (right) detecting at least 3 consecutive weeks increase (top) or decrease (bottom) in influenza activity, by symptom profile for the consultation rate (ILI or ARI), weekly number of swabs tested and country/region. Mean (circles), median (triangles) and 2.5 and 97.5 percentiles (line ranges) of times to detection are conditional on detection occurring. The probabilities of detection are obtained as the proportion of simulated datasets where the event occurs.

**Figure D.13:**
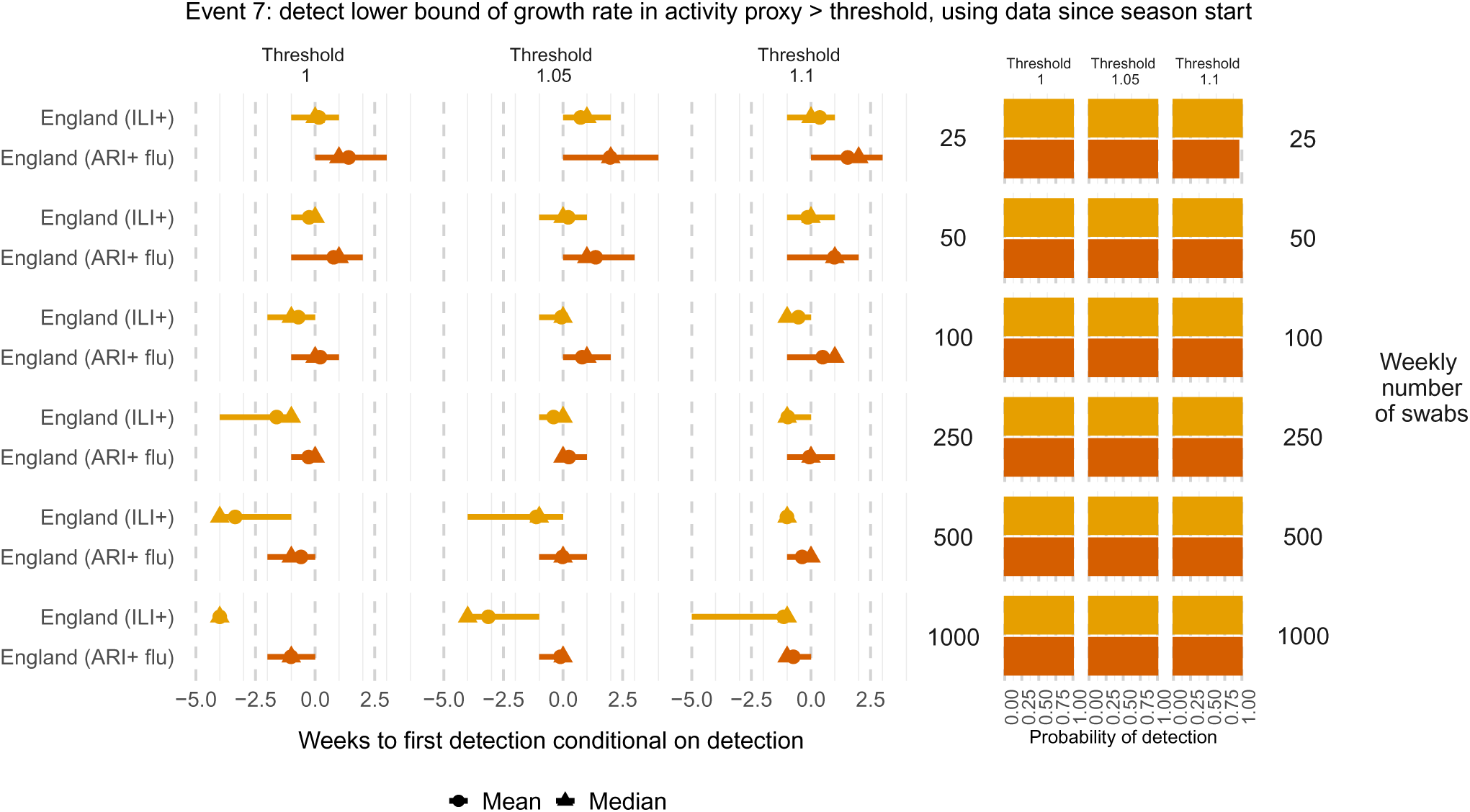
Times to (left) and probabilities of (right) detecting a decline rate in influenza activity smaller than specified thresholds, based on all data since the start of the season, by weekly number of swabs tested and symptom profile for the consultation rate (ILI or ARI). Mean (circles), median (triangles) and 2.5 and 97.5 percentiles (line ranges) of times to detection are conditional on detection occurring. The probabilities of detection are obtained as the proportion of simulated datasets where the event occurs.

**Figure D.14:**
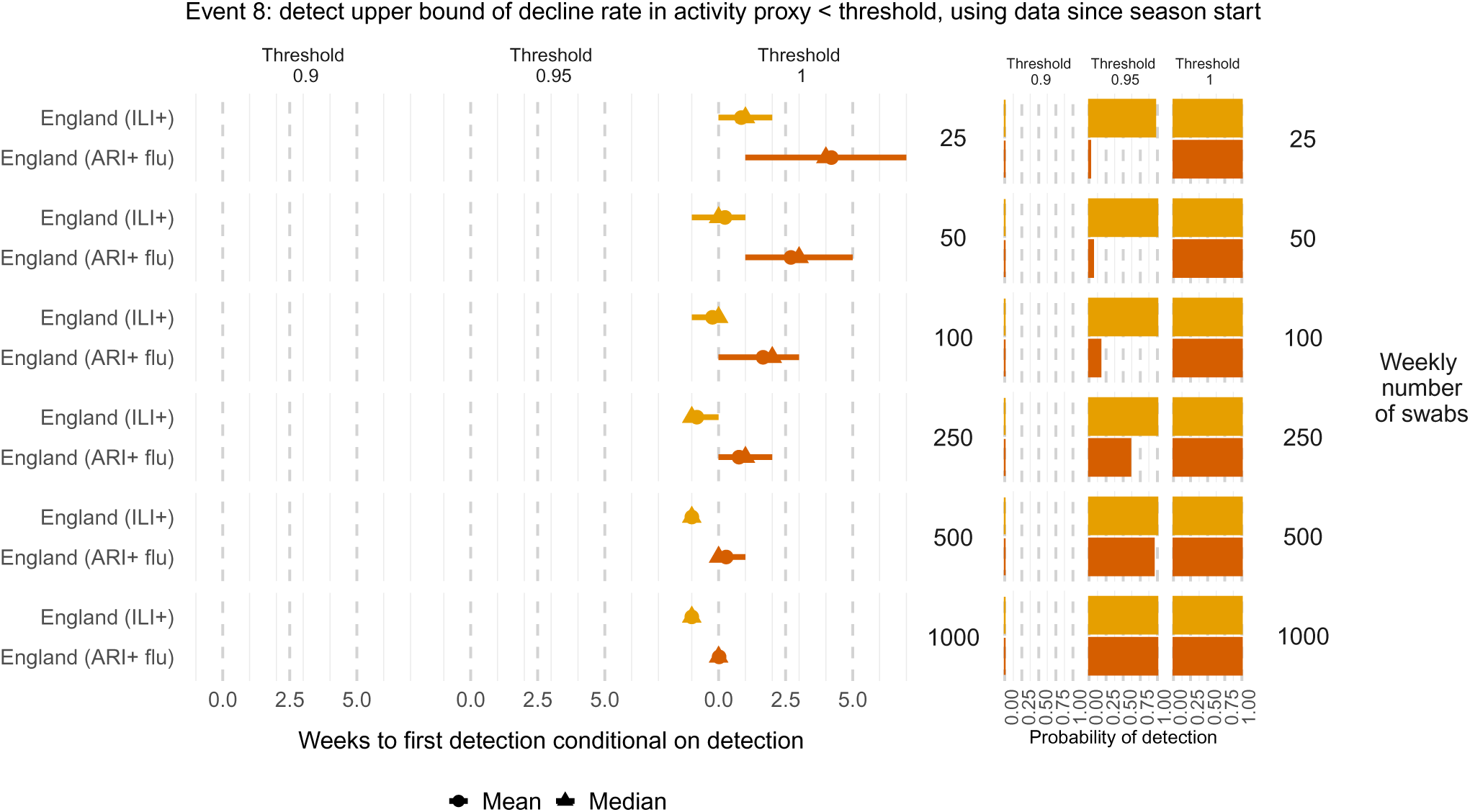
Times to (left) and probabilities of (right) detecting a decline rate in influenza activity significantly smaller than specified thresholds, based on all data since the start of the season, by weekly number of swabs tested and symptom profile for the consultation rate (ILI or ARI). Mean (circles), median (triangles) and 2.5 and 97.5 percentiles (line ranges) of times to detection are conditional on detection occurring. The probabilities of detection are obtained as the proportion of simulated datasets where the event occurs.

**Figure D.15:**
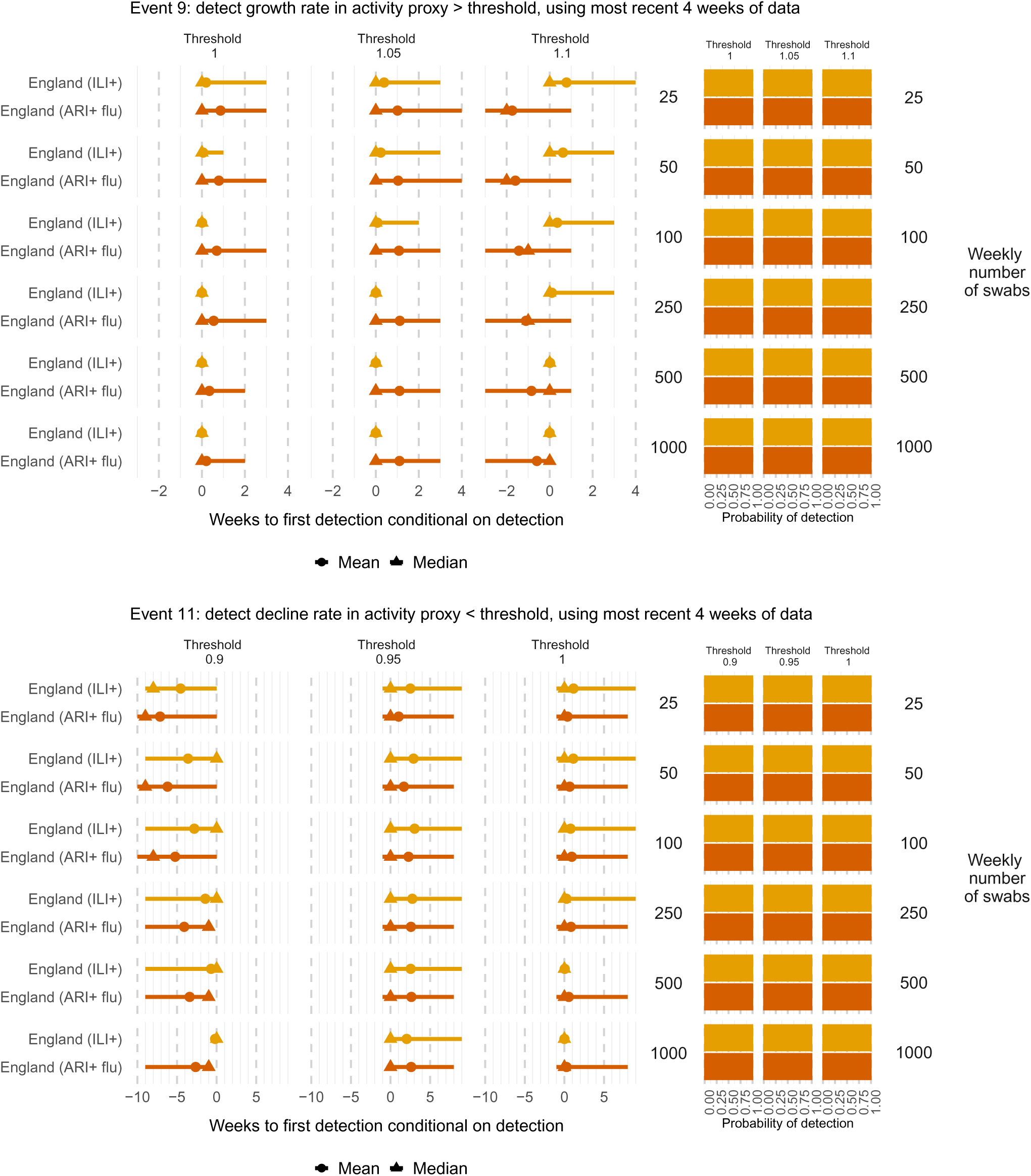
Times to (left) and probabilities of (right) detecting at a growth rate (top) or decline rate (bottom) in influenza activity greater or smaller respectively than specified thresholds, based on the last four weeks of data, by weekly number of swabs tested and symptom profile for the consultation rate (ILI or ARI). Mean (circles), median (triangles) and 2.5 and 97.5 percentiles (line ranges) of times to detection are conditional on detection occurring. The probabilities of detection are obtained as the proportion of simulated datasets where the event occurs.

**Figure D.16:**
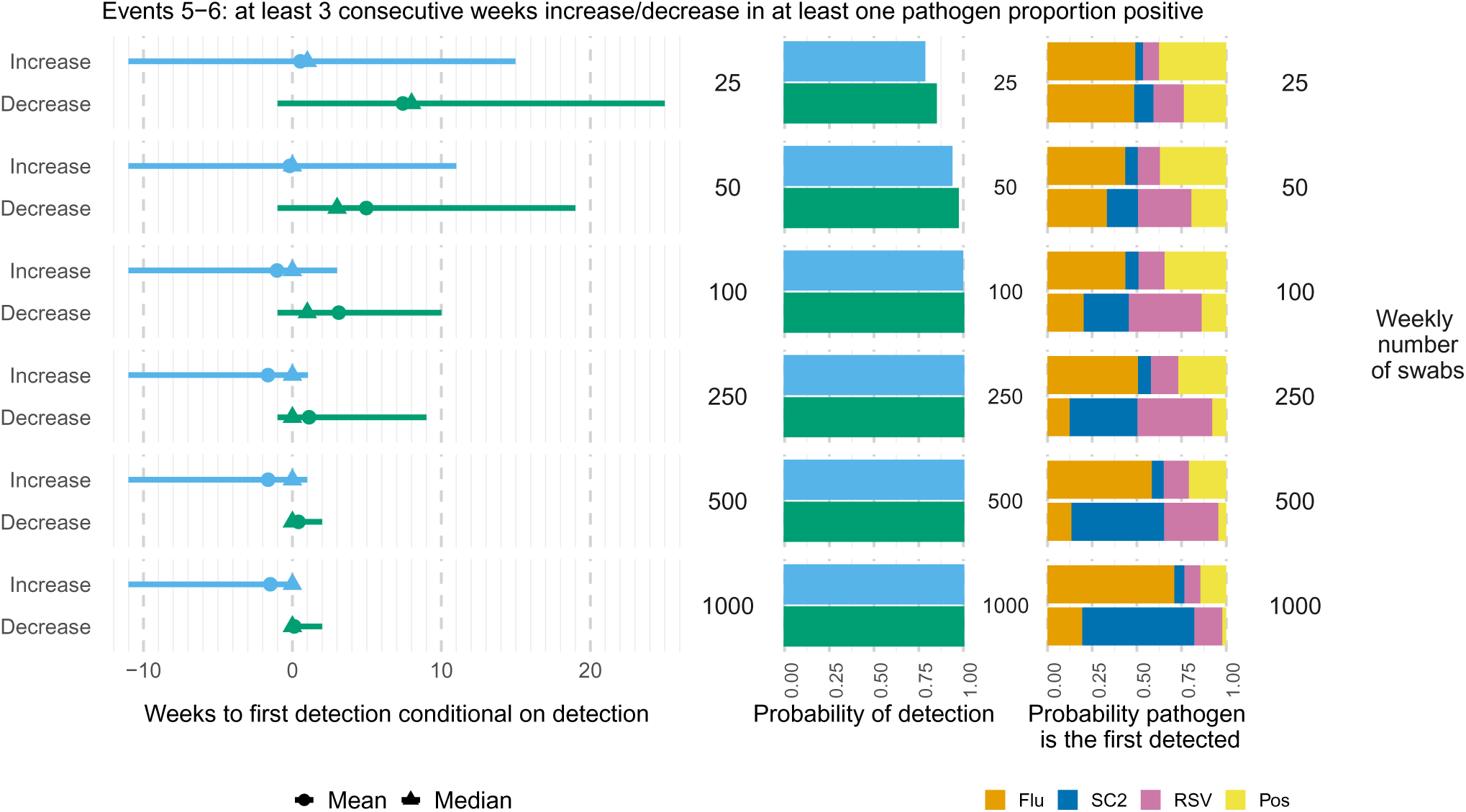
Times to (left) and probabilities of (middle) detecting at least 3 consecutive weeks increase or decrease in the proportion positive for a specific pathogen, conditional on not having tested positive for a previous pathogen, by weekly number of swabs tested. The right-hand panel shows the probability each pathogen is the first one detected. Mean (circles), median (triangles) and 2.5 and 97.5 percentiles (line ranges) of times to detection are conditional on detection occurring. The probabilities of detection are obtained as the proportion of simulated datasets where the event occurs.

**Figure D.17:**
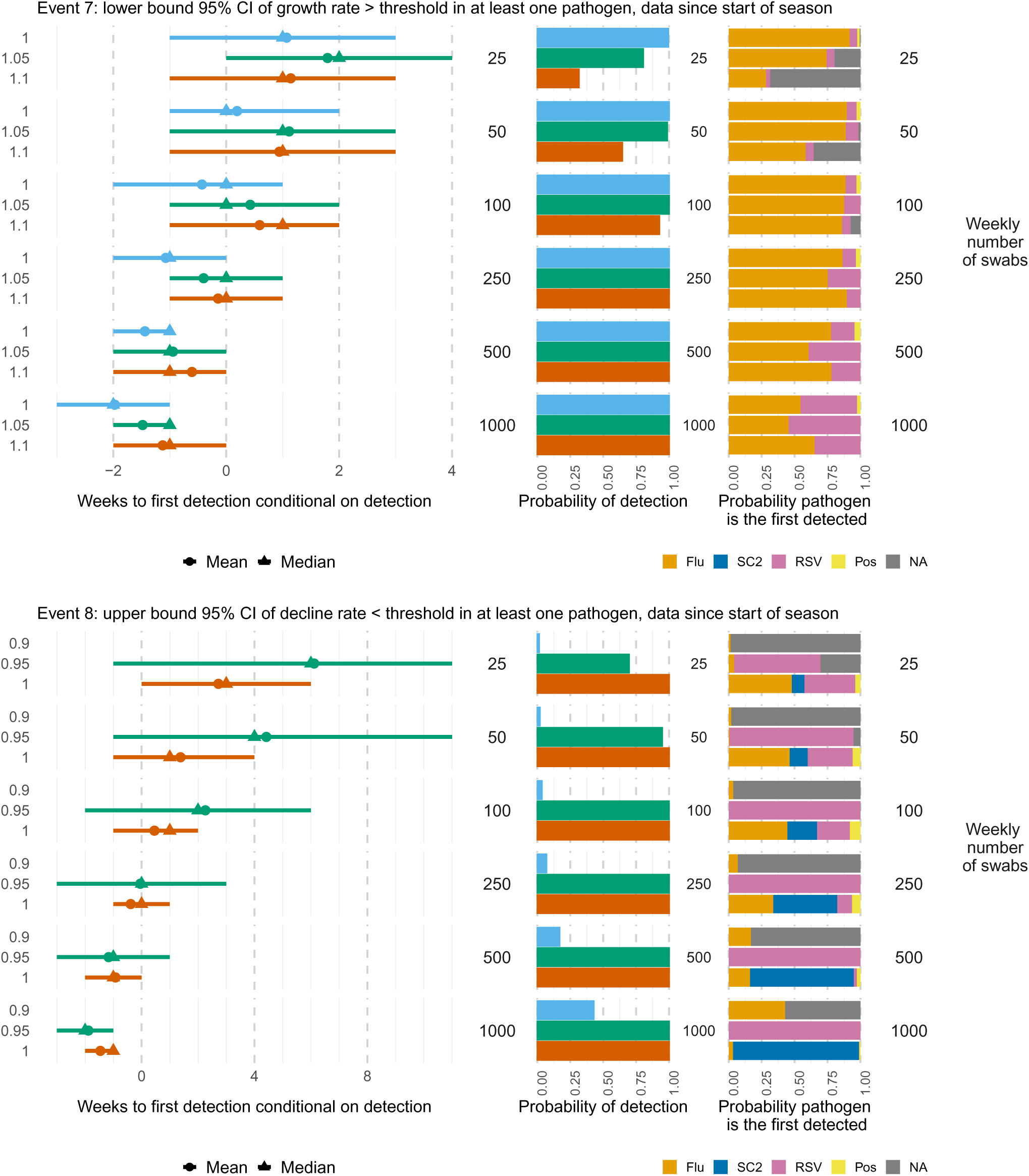
Times to (left) and probabilities of (middle) detecting at a growth rate (top) or decline rate (bottom) in at least one pathogen proportion positive in England significantly greater or smaller respectively than specified thresholds, based on all data since the start of the season, by weekly number of swabs tested. The right-hand panel shows the probability each pathogen is the first one detected. Mean (circles), median (triangles) and 2.5 and 97.5 percentiles (line ranges) of times to detection are conditional on detection occurring. The probabilities of detection are obtained as the proportion of simulated datasets where the event occurs. Note that when either the event does not occur in the ground truth data or in any of the simulated datasets, then a distribution of detection times is not available (e.g. for the highest / lowest thresholds).

**Figure D.18:**
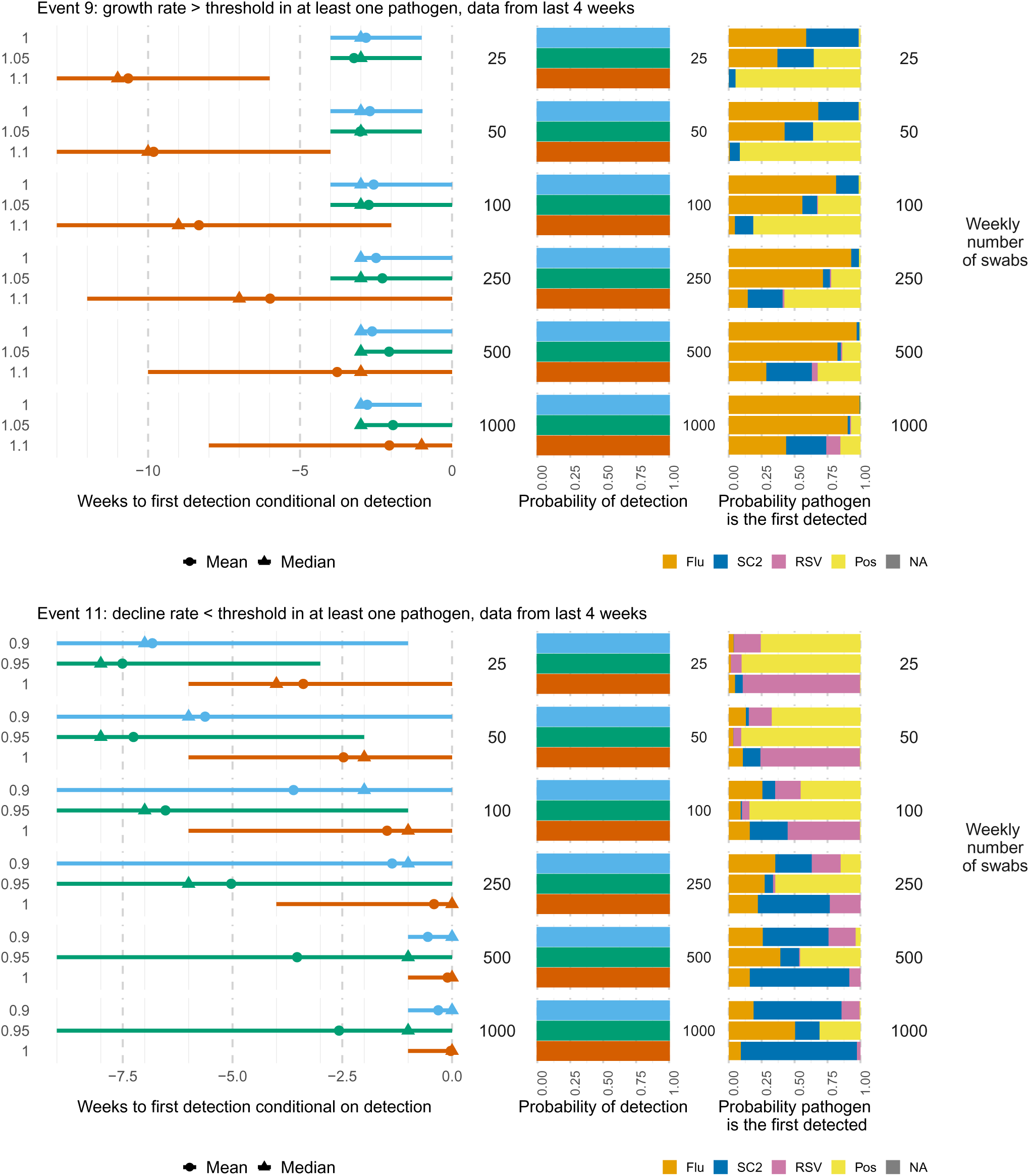
Times to (left) and probabilities of (middle) detecting at a growth (top) or decline (bottom) rate in proportion positive smaller/larger than specified thresholds in at least one pathogen, based on the last four weeks of data, by weekly number of swabs tested. The right-hand panel shows the probability each pathogen is the first one detected. Mean (circles), median (triangles) and 2.5 and 97.5 percentiles (line ranges) of times to detection are conditional on detection occurring. The probabilities of detection are obtained as the proportion of simulated datasets where the event occurs.

**Figure D.19:**
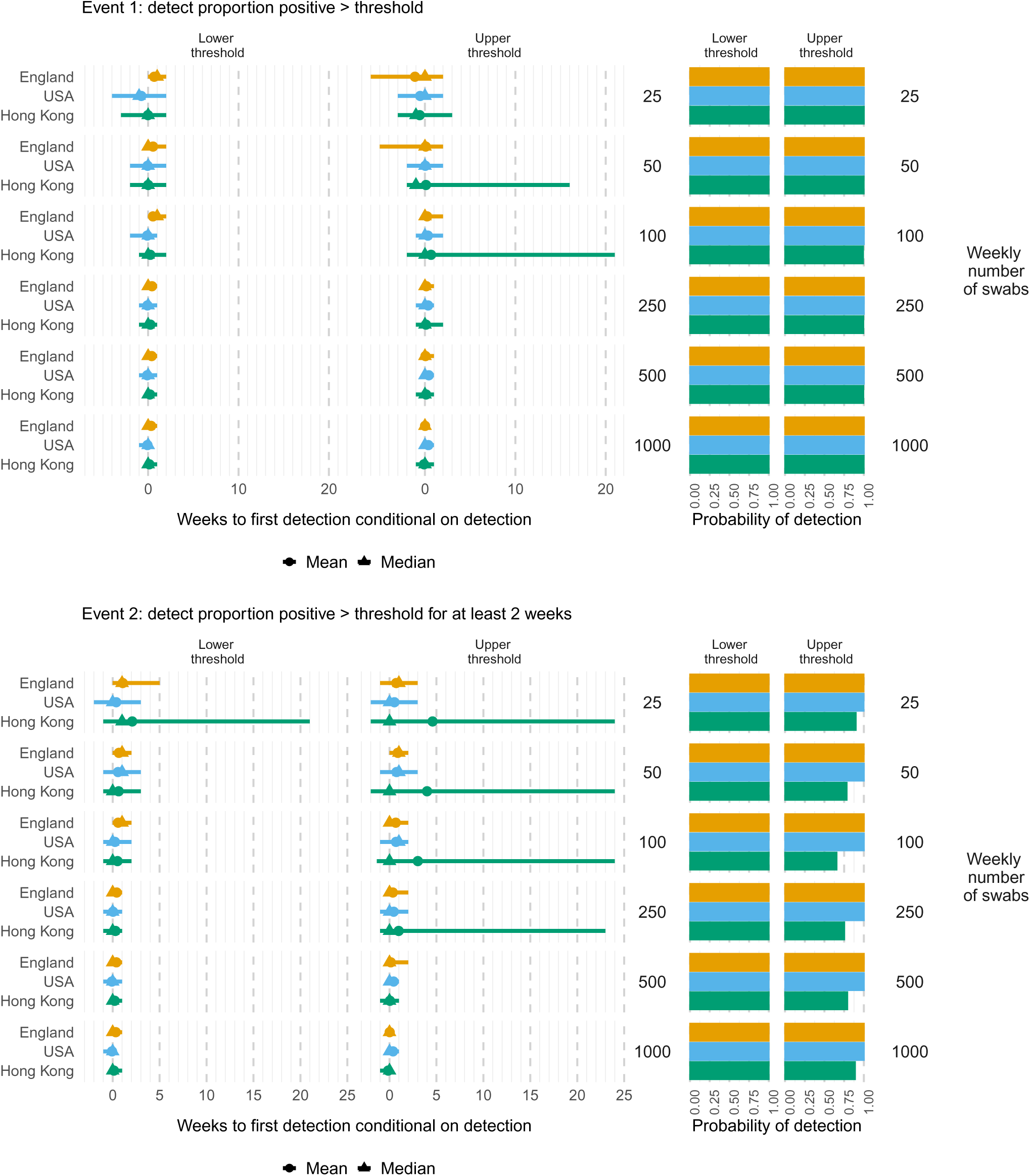
Times to (left) and probabilities of (right) detecting proportions of ILI cases testing positive for influenza greater than a country/region-specific lower or higher threshold (Table B.1), for 1 week (top) or at least 2 consecutive weeks (bottom), by weekly number of swabs tested and country/region. Mean (circles), median (triangles) and 2.5 and 97.5 percentiles (line ranges) of times to detection are conditional on detection occurring. The probabilities of detection are obtained as the proportion of simulated datasets where the event occurs.

**Figure D.20:**
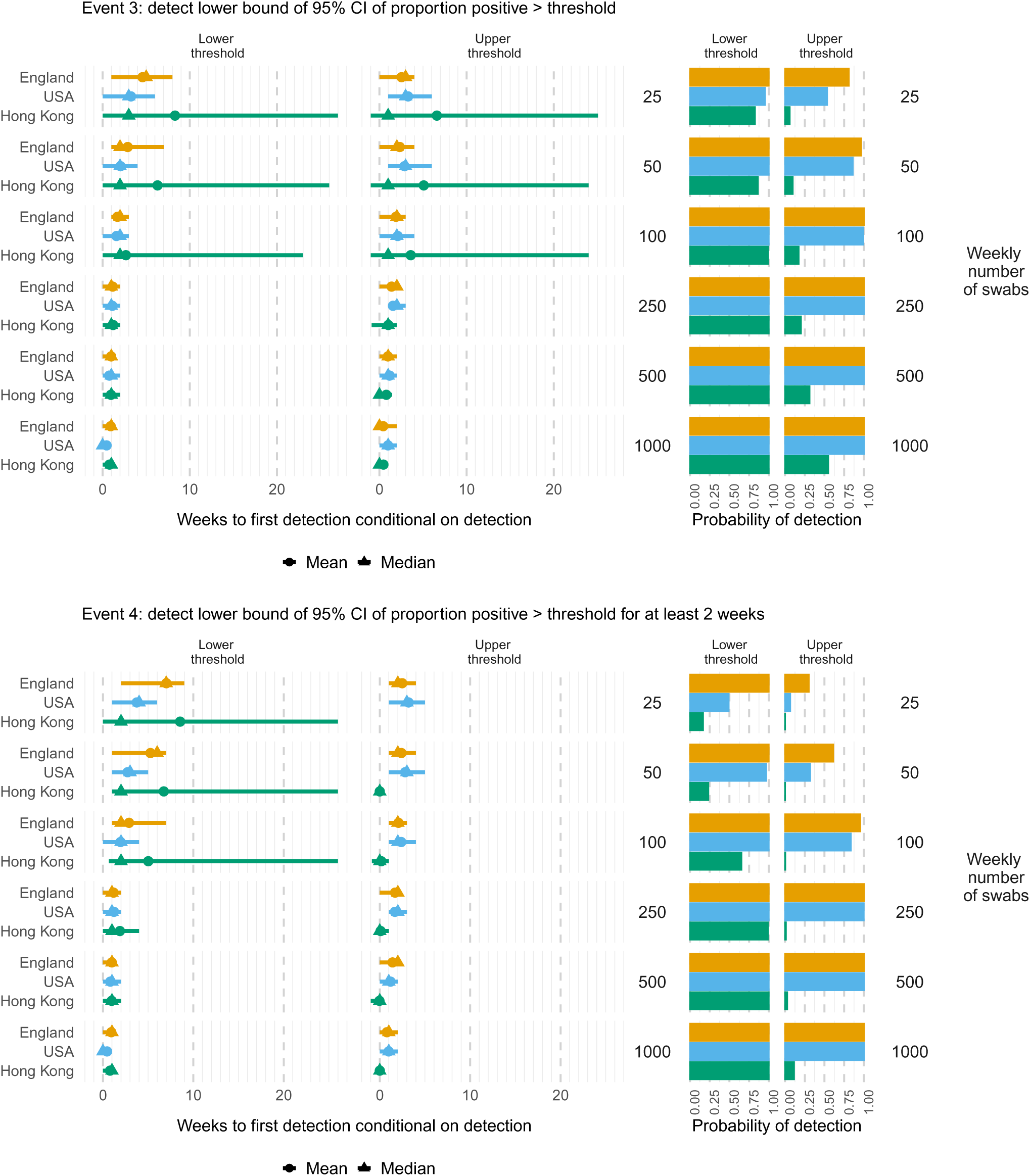
Times to (left) and probabilities of (right) detecting proportions of ILI cases testing positive for influenza significantly greater than a country/region-specific lower or higher threshold (Table B.1), for 1 week (top) or at least 2 consecutive weeks (bottom), by weekly number of swabs tested and country/region. Mean (circles), median (triangles) and 2.5 and 97.5 percentiles (line ranges) of times to detection are conditional on detection occurring. The probabilities of detection are obtained as the proportion of simulated datasets where the event occurs.

**Figure D.21:**
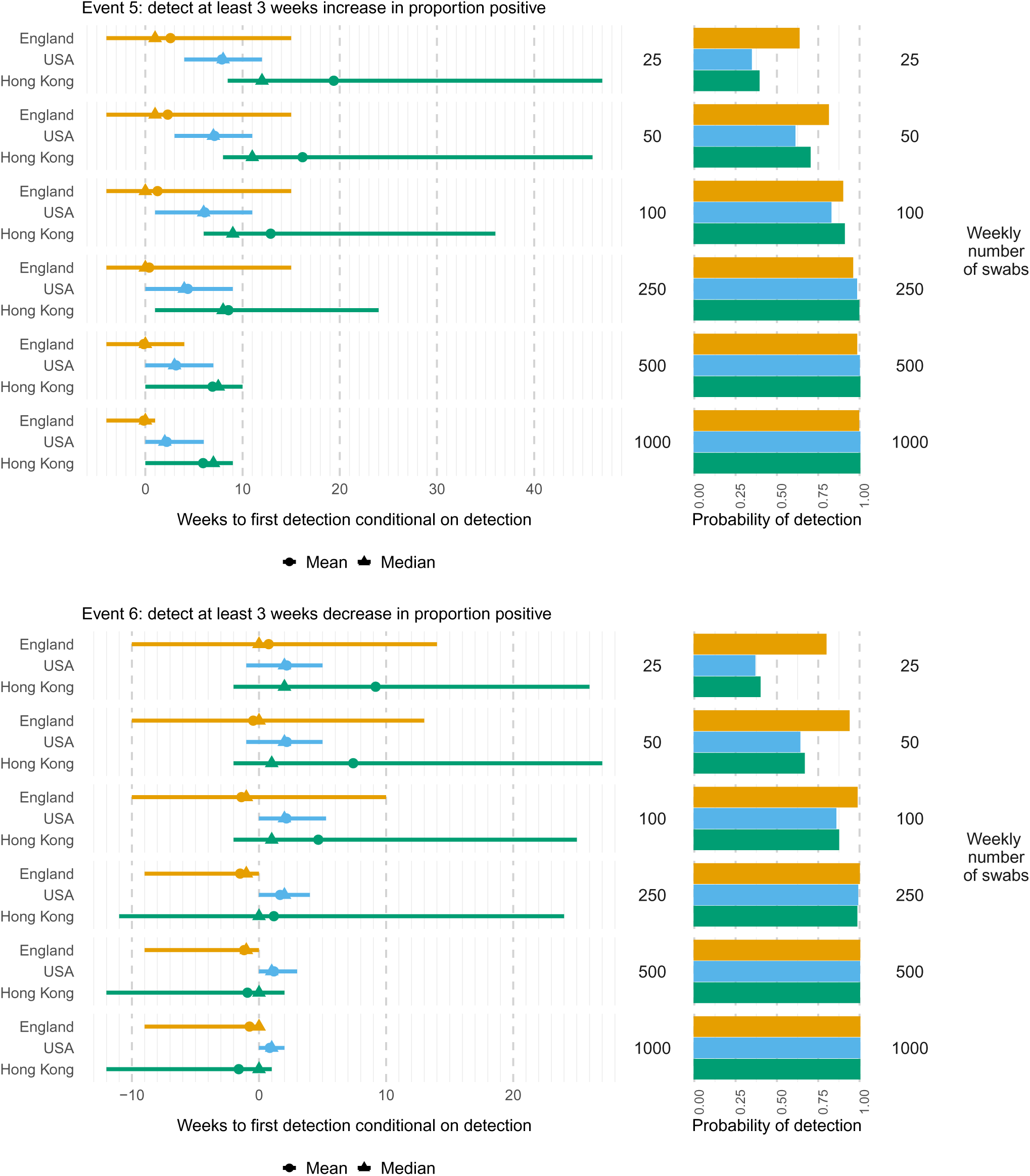
Times to (left) and probabilities of (right) detecting at least three consecutive weeks increase (top: positive trend) or decrease (bottom: negative trend) in the proportions of ILI cases testing positive for influenza, by weekly number of swabs tested and country/region. Mean (circles), median (triangles) and 2.5 and 97.5 percentiles (line ranges) of times to detection are conditional on detection occurring. The probabilities of detection are obtained as the proportion of simulated datasets where the event occurs.

**Figure D.22:**
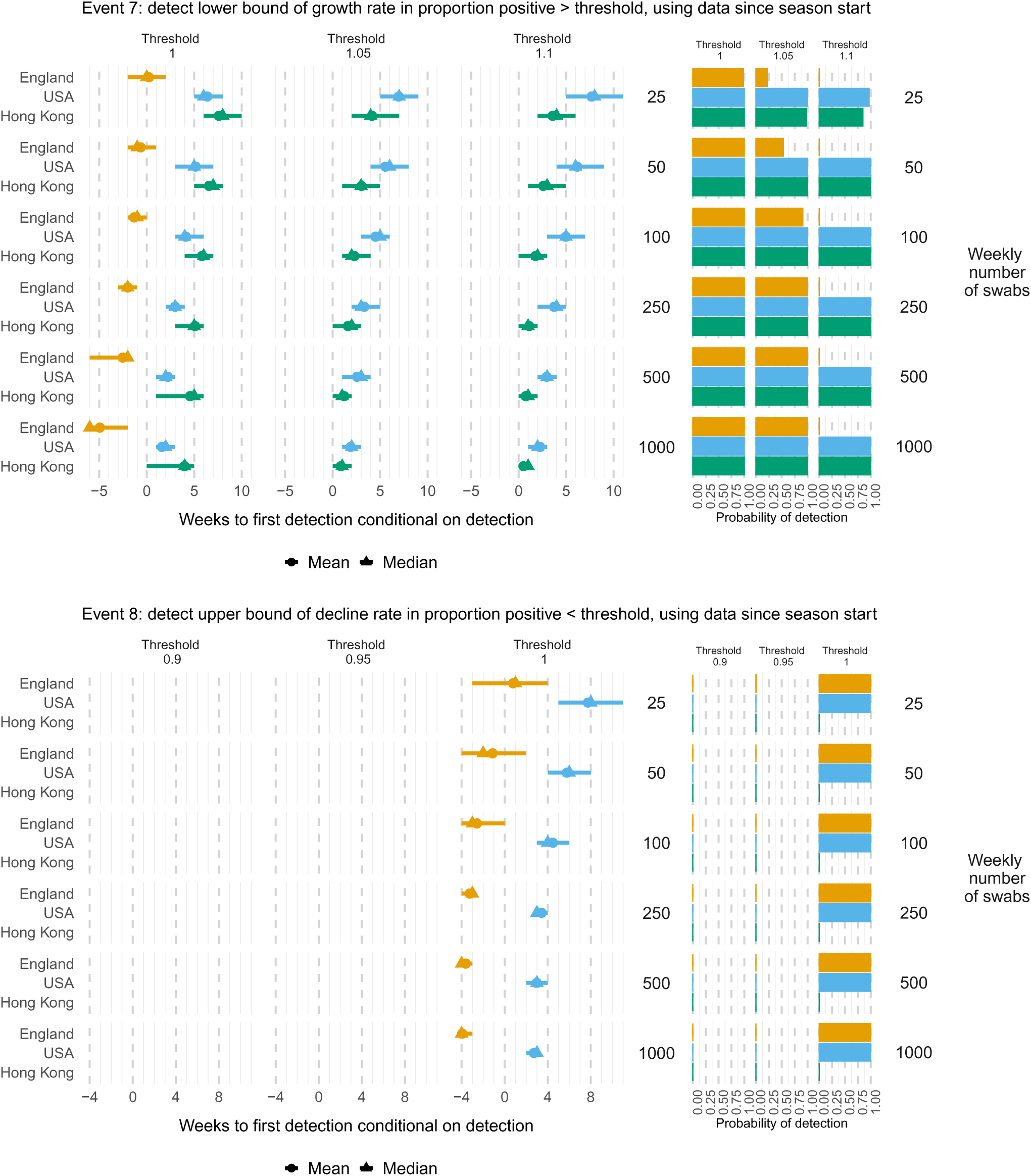
Times to (left) and probabilities of (right) detecting a growth rate significantly greater than a threshold (top) or decline rate significantly lower than a threshold (bottom) in the cumulative data to date on the proportions of ILI cases testing positive for influenza, by weekly number of swabs tested and country/region. Mean (circles), median (triangles) and 2.5 and 97.5 percentiles (line ranges) of times to detection are conditional on detection occurring. The probabilities of detection are obtained as the proportion of simulated datasets where the event occurs.

**Figure D.23:**
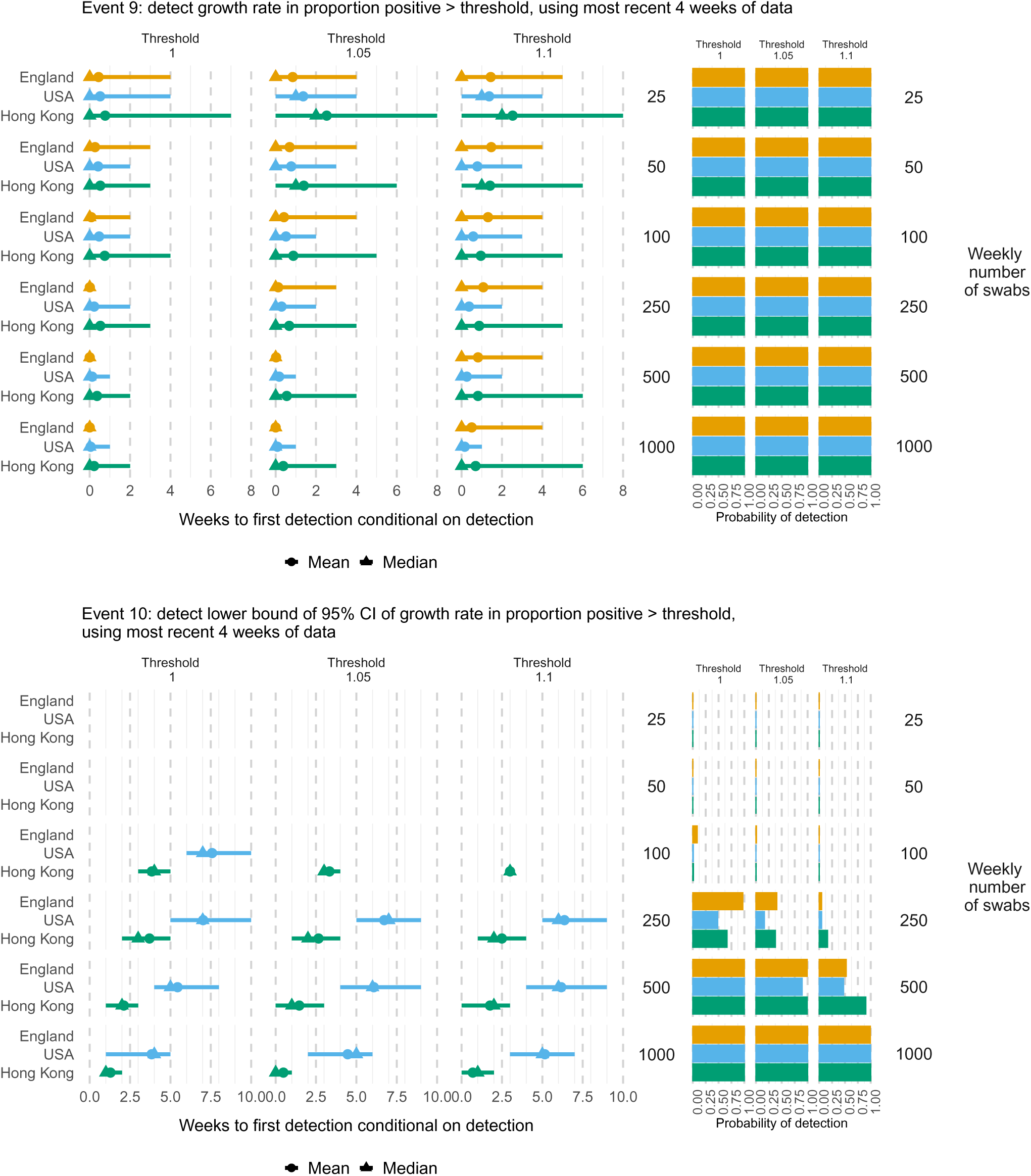
Times to (left) and probabilities of (right) detecting a growth rate greater than a threshold (top) with significance (bottom) in the most recent 4 weeks of data on the proportions of ILI cases testing positive for influenza, by weekly number of swabs tested and country/region. Mean (circles), median (triangles) and 2.5 and 97.5 percentiles (line ranges) of times to detection are conditional on detection occurring. The probabilities of detection are obtained as the proportion of simulated datasets where the event occurs.

**Figure D.24:**
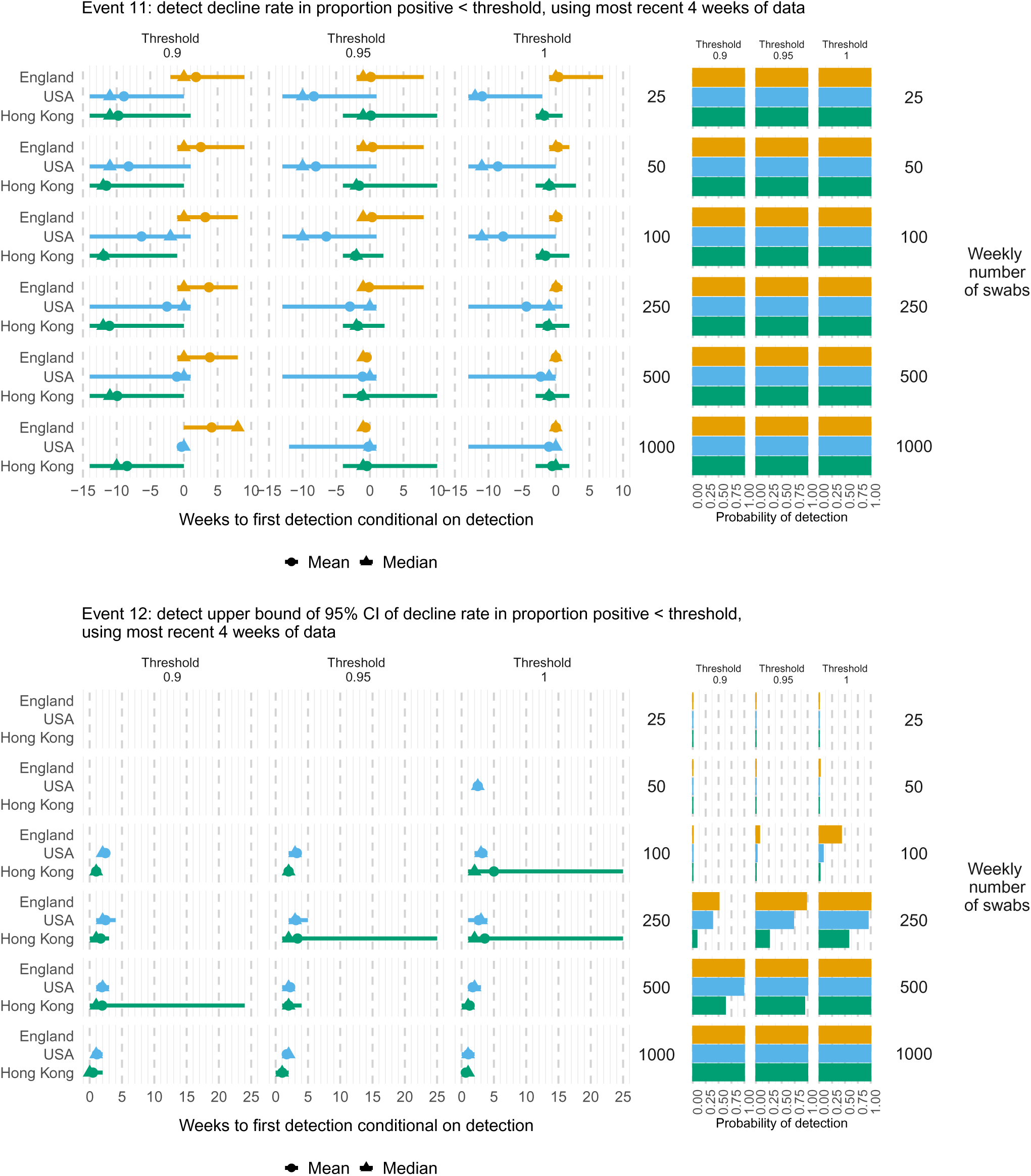
Times to (left) and probabilities of (right) detecting a decline rate less than a threshold (top) with significance (bottom) in the most recent 4 weeks of data on the proportions of ILI cases testing positive for influenza, by weekly number of swabs tested and country/region. Mean (circles), median (triangles) and 2.5 and 97.5 percentiles (line ranges) of times to detection are conditional on detection occurring. The probabilities of detection are obtained as the proportion of simulated datasets where the event occurs.

**Figure D.25:**
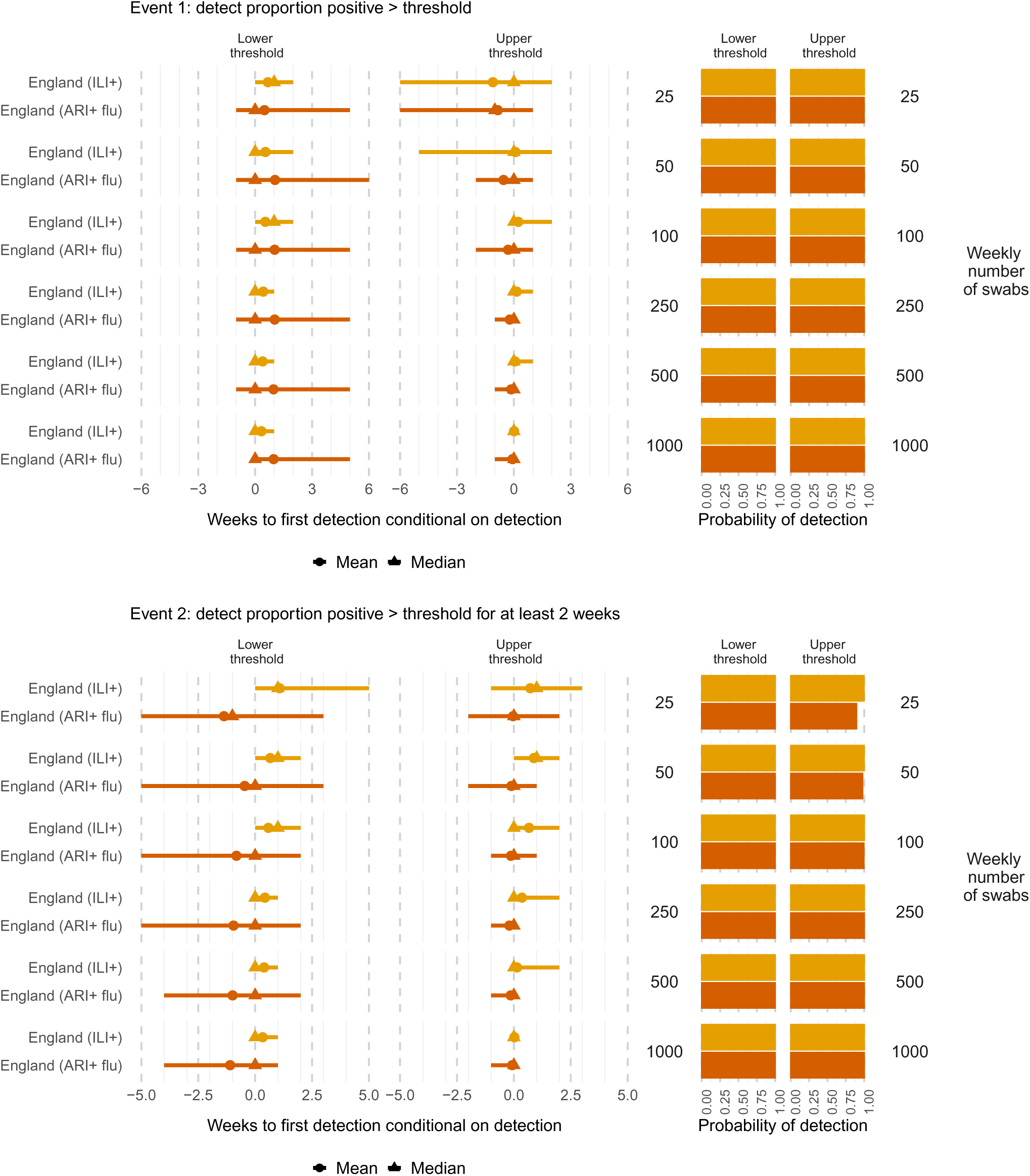
Times to (left) and probabilities of (right) detecting the proportion positive for influenza greater than a threshold (top), for at least two weeks (bottom), by weekly number of swabs tested and denominator (ILI or all swabs). Mean (circles), median (triangles) and 2.5 and 97.5 percentiles (line ranges) of times to detection are conditional on detection occurring. The probabilities of detection are obtained as the proportion of simulated datasets where the event occurs.

**Figure D.26:**
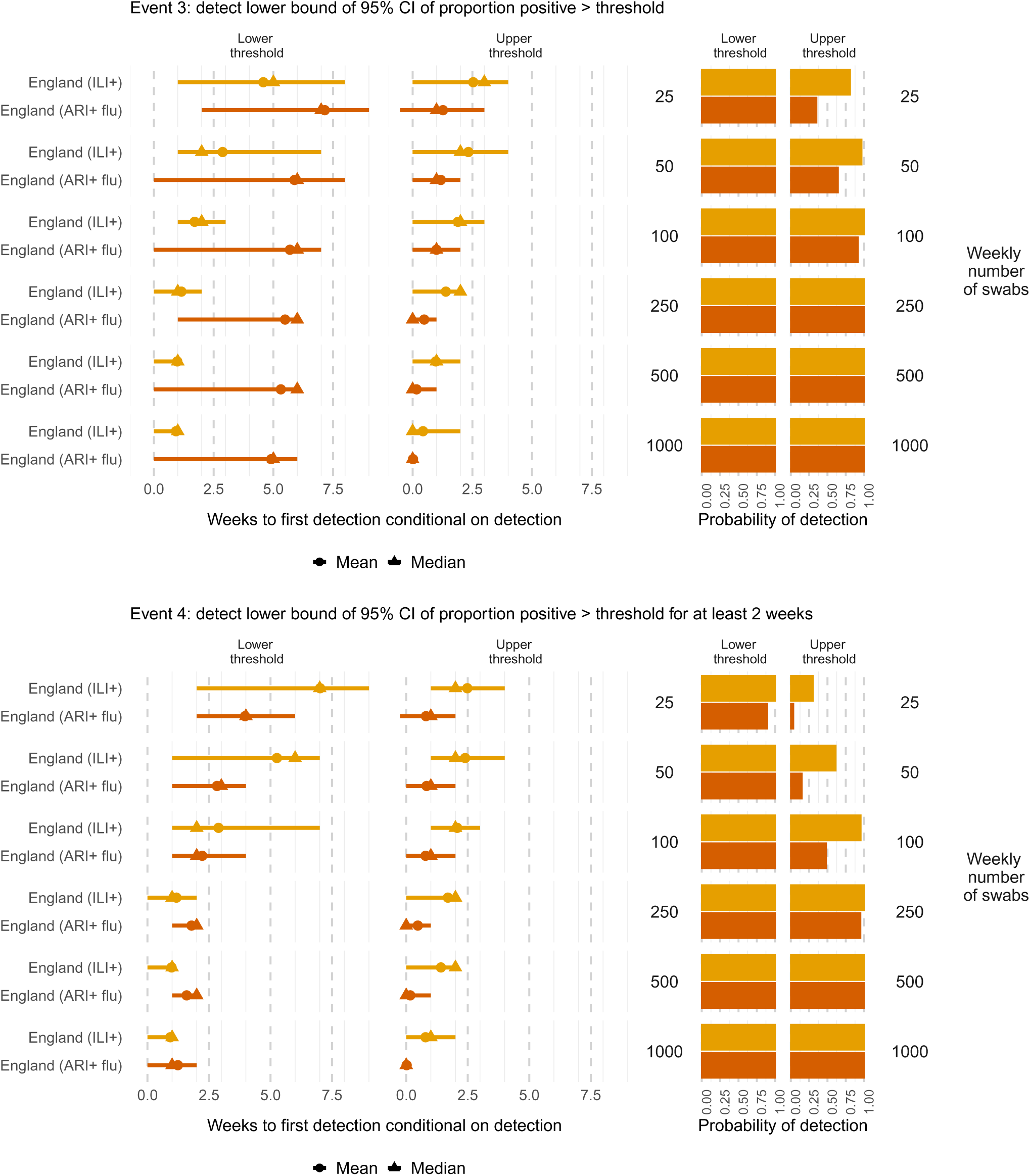
Times to (left) and probabilities of (right) detecting proportions testing positive for influenza significantly greater than a threshold (top), for at least two weeks (bottom), by weekly number of swabs tested and denominator (ILI or all swabs). Mean (circles), median (triangles) and 2.5 and 97.5 percentiles (line ranges) of times to detection are conditional on detection occurring. The probabilities of detection are obtained as the proportion of simulated datasets where the event occurs.

**Figure D.27:**
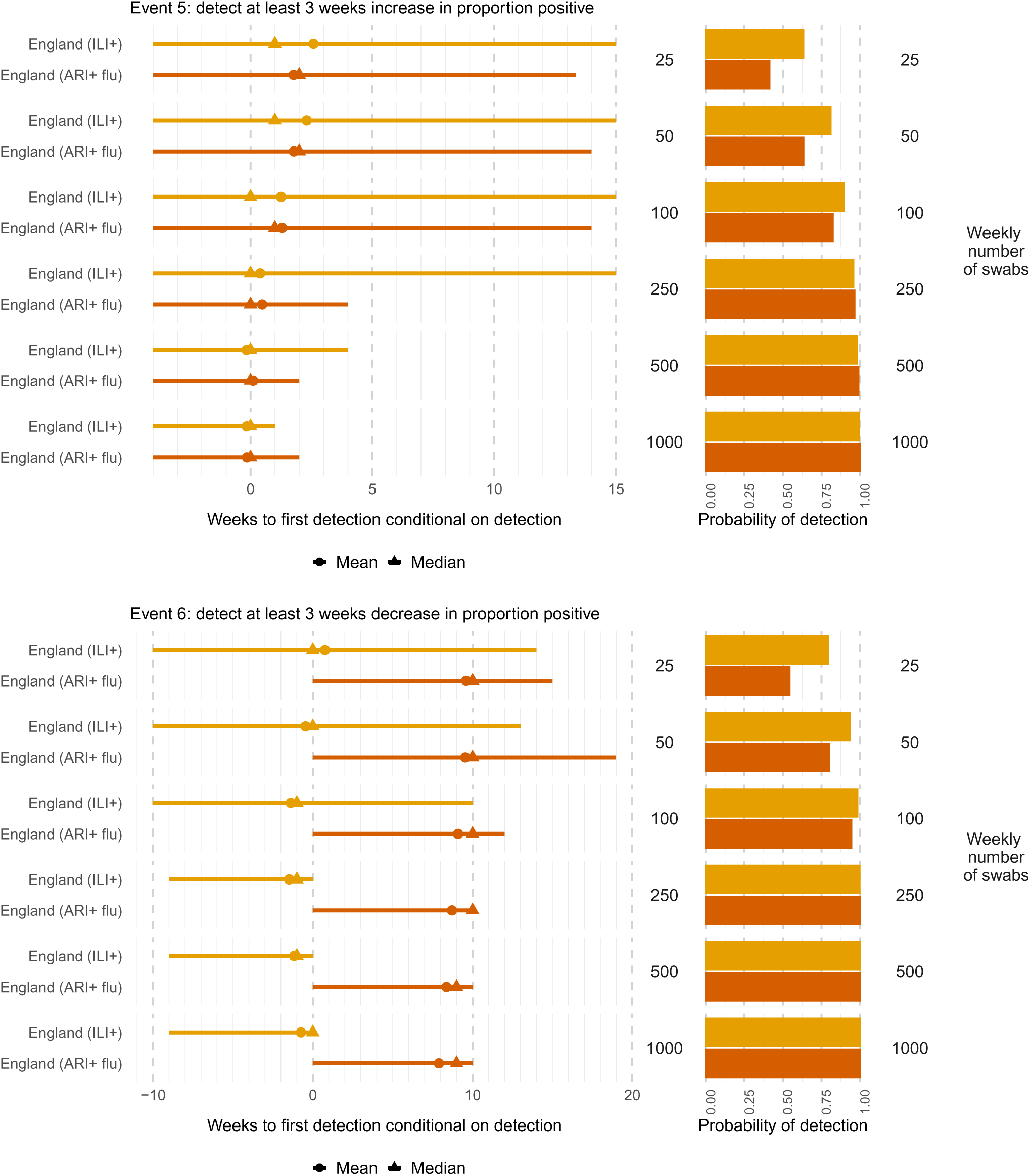
Times to (left) and probabilities of (right) detecting at least three weeks consecutive increase (top) or decrease (bottom) in the proportion testing positive for influenza, by weekly number of swabs tested and denominator (ILI or all swabs). Mean (circles), median (triangles) and 2.5 and 97.5 percentiles (line ranges) of times to detection are conditional on detection occurring. The probabilities of detection are obtained as the proportion of simulated datasets where the event occurs.

**Figure D.28:**
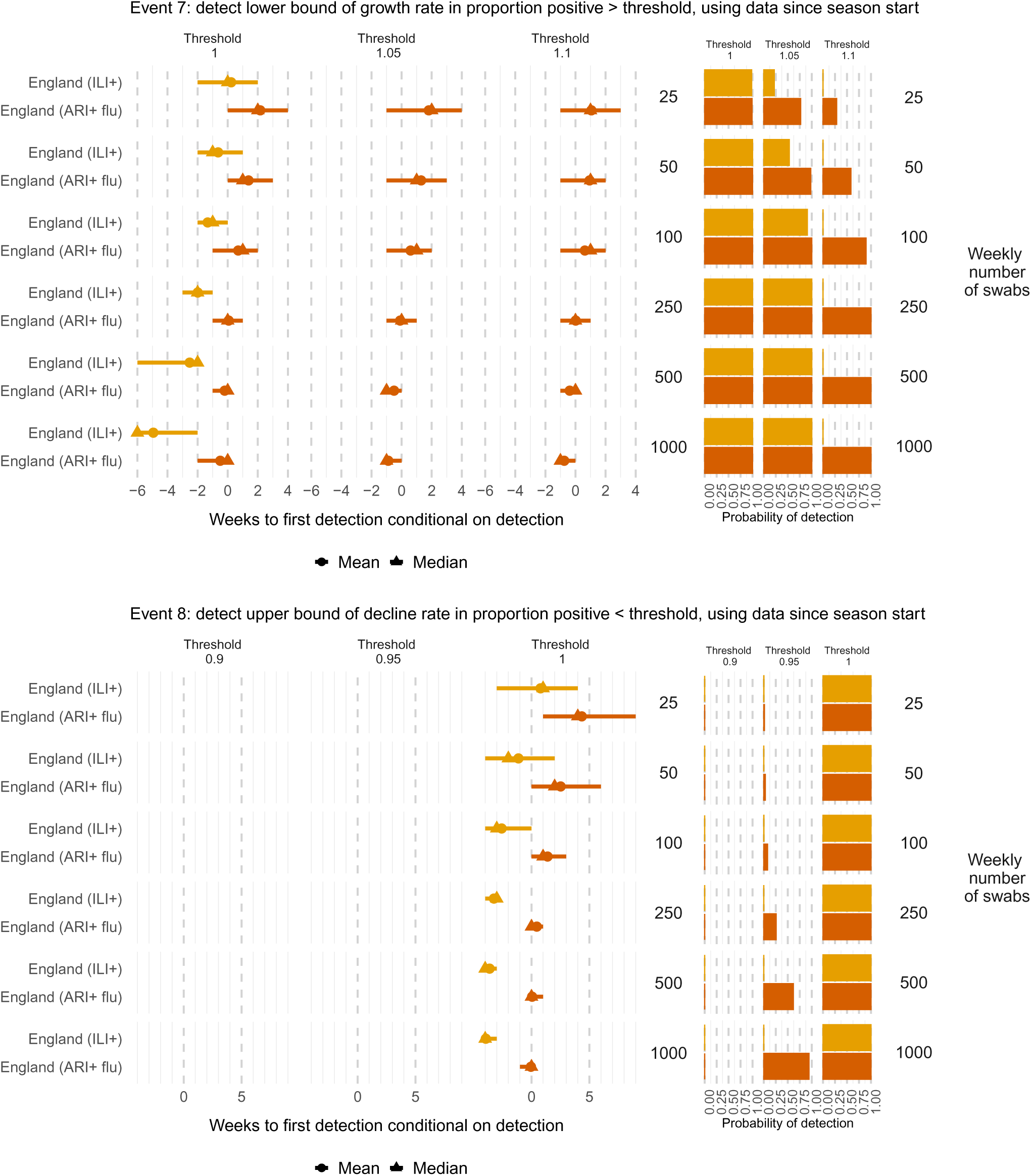
Times to (left) and probabilities of (right) detecting a growth rate in the proportion testing positive for influenza significantly greater (top) or less (bottom) than a threshold, by weekly number of swabs tested and denominator (ILI or all swabs). Mean (circles), median (triangles) and 2.5 and 97.5 percentiles (line ranges) of times to detection are conditional on detection occurring. The probabilities of detection are obtained as the proportion of simulated datasets where the event occurs.

**Figure D.29:**
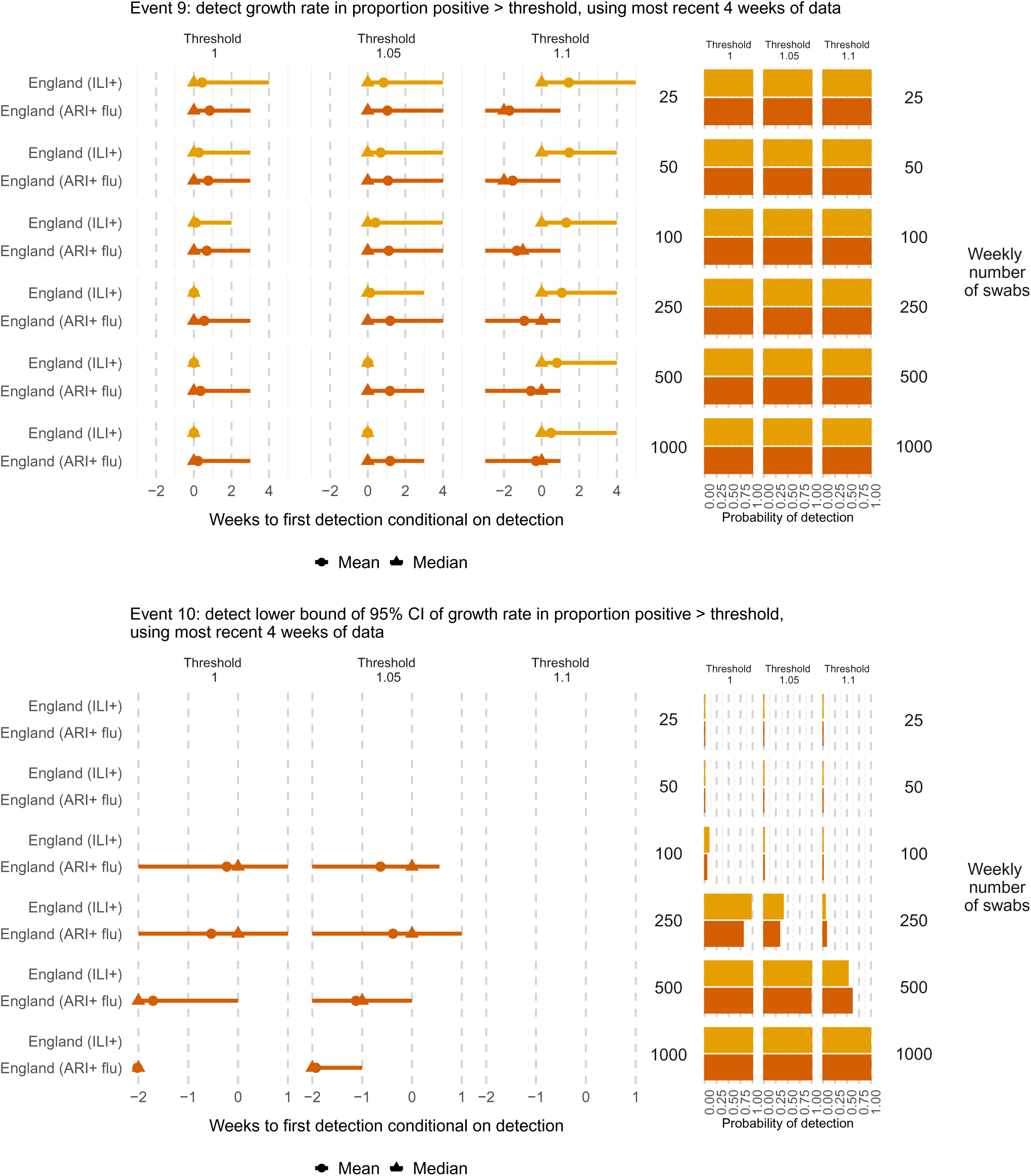
Times to (left) and probabilities of (right) detecting a growth rate greater (top) or significantly greater (bottom) than a threshold, based on the most recent 4 weeks of data on the proportion testing positive for influenza, by weekly number of swabs tested and denominator (ILI or all swabs). Mean (circles), median (triangles) and 2.5 and 97.5 percentiles (line ranges) of times to detection are conditional on detection occurring. The probabilities of detection are obtained as the proportion of simulated datasets where the event occurs.

**Figure D.30:**
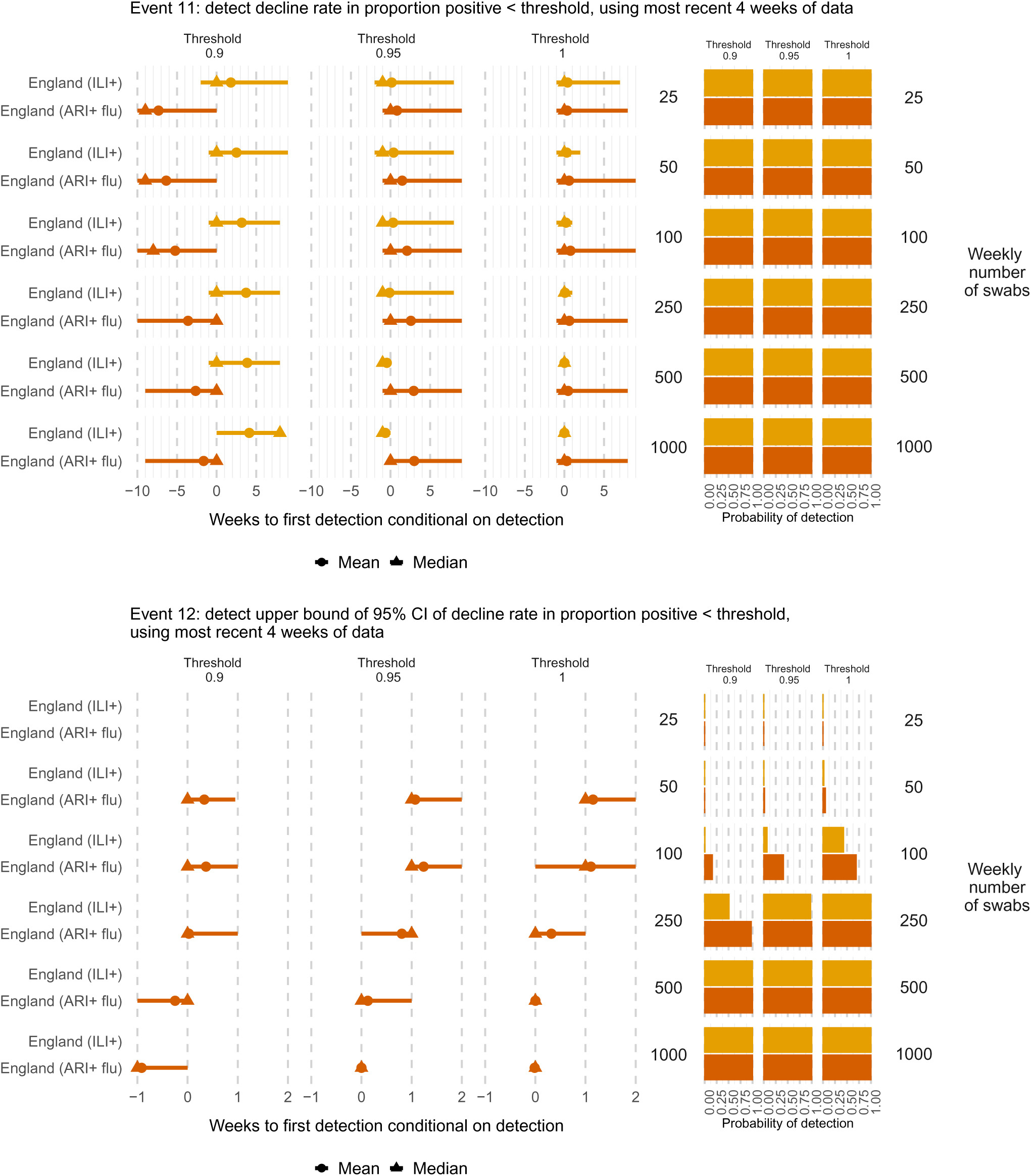
Times to (left) and probabilities of (right) detecting a decline rate smaller (top) or significantly smaller (bottom) than a threshold, based on the most recent 4 weeks of data on the proportion testing positive for influenza, by weekly number of swabs tested and denominator (ILI or all swabs). Mean (circles), median (triangles) and 2.5 and 97.5 percentiles (line ranges) of times to detection are conditional on detection occurring. The probabilities of detection are obtained as the proportion of simulated datasets where the event occurs.

## Notes

### Competing Interest Statement

The Immunisations and Vaccine Preventable Diseases division at UKHSA has undertaken post-marketing surveillance and regulatory analyses requested by influenza vaccine manufacturers for which cost-recovery charges have been made.

### Author Declarations

This study used simulated data. The data were simulated to mimic aggregate data that are publicly available, as detailed in Section 2.1 of the manuscript. The pseudo-anonymised individual-level surveillance data underlying the publicly available aggregate data were collected by NHS England and the UK Health Security Agency with permissions granted under Regulation 3 of The Health Service (Control of Patient Information) Regulations 2002, and without explicit patient permission under Section 251 of the NHS Act 2006.

